# Buffering and non-monotonic behavior of gene dosage response curves for human complex traits

**DOI:** 10.1101/2024.11.11.24317065

**Authors:** Nikhil Milind, Courtney J. Smith, Huisheng Zhu, Tamara Gjorgjieva, Jeffrey P. Spence, Jonathan K. Pritchard

**Affiliations:** Department of Genetics, Stanford University, CA, USA; Department of Biology, Stanford University, CA, USA; Institute for Human Genetics, University of California, San Francisco, CA, USA; Department of Epidemiology & Biostatistics, University of California, San Francisco, CA, USA

## Abstract

The genome-wide burdens of deletions, loss-of-function mutations, and duplications correlate with many traits. Curiously, for most of these traits, variants that decrease expression have the same genome-wide average direction of effect as variants that increase expression. This seemingly contradicts the intuition that for individual genes reducing expression should have the opposite effect on a phenotype as increasing expression. To understand this paradox, we use the gene dosage response curve (GDRC), which relates changes in gene expression to expected changes in phenotype. We show that, for many traits, GDRCs are systematically biased in one trait direction relative to the other, and we develop a simple theoretical model that explains this bias in trait direction. Our results have broad implications for complex traits, drug discovery, and statistical genetics.

## Introduction

Genome-wide association studies (GWASs) have identified thousands of variants associated with complex traits [1, 2], most of which are in non-coding regions of the genome [3]. This suggests that a substantial proportion of phenotypic variance is likely explained by variation in gene expression [3–8].

One source of variation in gene expression is gain or loss of functional copies of a gene [9]. Such gene dosage changes can have phenotypic consequences, with aneuploidies being an extreme example [10–13]. Copy number variants (CNVs), which typically perturb the dosage of several genes, are another source of expression variation [14]. CNVs segregate in humans [15–18] and have measurable effects on various traits and disorders [19–27], presumably by varying the expression of one or more of the genes with atypical copy numbers [9].

There has been substantial work associating the total genome-wide CNV burden to traits [19–22, 24, 25, 27–30]. An individual’s genome-wide burden is estimated by counting the total number of CNVs carried by an individual, regardless of which genes are affected. Associating this burden with a trait can be thought of as roughly estimating how much deleting or duplicating a random gene affects the trait. The large number of traits significantly associated with genome-wide deletion burden suggests that the effect of reducing the expression of a randomly chosen gene is often biased in a particular trait direction. Interestingly, genome-wide duplication burden and genome-wide deletion burden often have the same direction of effect (Appendix A.2) [20–22]. That is, increasing expression often has the same effect as decreasing expression on average. This seemingly contradicts the intuition that, for a given gene, increasing expression should have the opposite effect of decreasing expression.

To make sense of these genome-wide CNV burden test results from the perspective of what is happening at individual genes, we estimated the gene-level effects of loss-of-function (LoF) variants, deletions, and duplications using whole-exome sequencing (WES) and whole-genome sequencing (WGS) data from the UK Biobank (UKB) for 94 continuous traits [31, 32].

Our analysis revealed that, across traits, the average effects of gene deletions and duplications are often non-zero and usually affect the trait in the same direction. The data also confirm the intuition that, for genes with large effects on traits, deletions generally have opposite effects to duplications. To explain these counterintuitive observations, we use the concept of gene dosage response curves (GDRCs), study their properties, and develop a simple model of their evolution.

## Results

### Gene-level burden tests for loss-of-function variants and duplications

To better understand how variants with different effects on dosage impact traits, we performed burden tests using LoF variants, deletions, and duplications in the UKB. Our primary analyses focus on LoF variant burden tests because they are better powered than tests based on deletions, but deletion-based tests showed broadly similar patterns of effect (Appendix I.5). We took care to keep the analysis of the different variant types as similar as possible so that the burden effect sizes could be interpreted jointly. We used the gene burden test implemented in REGENIE [33], and included covariates to correct for sex, age, batch effects, and other sources of confounding (Methods). We also performed numerous quality control checks (Appendices H.1 and H.2). We selected 410 continuous traits from various blood biochemistry, blood count, blood metabolite, and anthropometric measurements available in the UKB (Data Availability). We filtered these to a subset of 94 continuous traits with sufficient data and burden signal, and used these for most of our analyses (Methods). The summary statistics from these burden tests are provided as a resource to the community (Data Availability).

### Connecting gene-level curves with genome-wide burden effects

Estimating the effect of genes on complex traits is central to understanding the mechanisms underlying trait biology. Changes in gene expression, which we refer to as ‘gene dosage’, are often implicitly assumed to have a linear effect on a trait (Figure 1A). For example, transcriptome-wide association studies (TWASs) use a linear model for the relationship between predicted expression and trait value [5, 6]. Under such a linear model, LoF and duplication burden estimates are expected to have opposite effects for any given gene (Figure 1A) because LoF variants and duplications likely affect traits by modulating gene dosage in *cis* (Appendix I.5), which we observe empirically with protein levels (Appendix A.5). The genome-wide burden effects for LoF variants and duplications, which can be thought of as averaging across these linear models for all genes, would be expected to have opposite signs. Thus, a linear model is incompatible with the genome-wide burden effects observed for various complex traits (Appendix A.2), and suggests that these relationships must be, at a minimum, non-linear.

**Figure 1.**
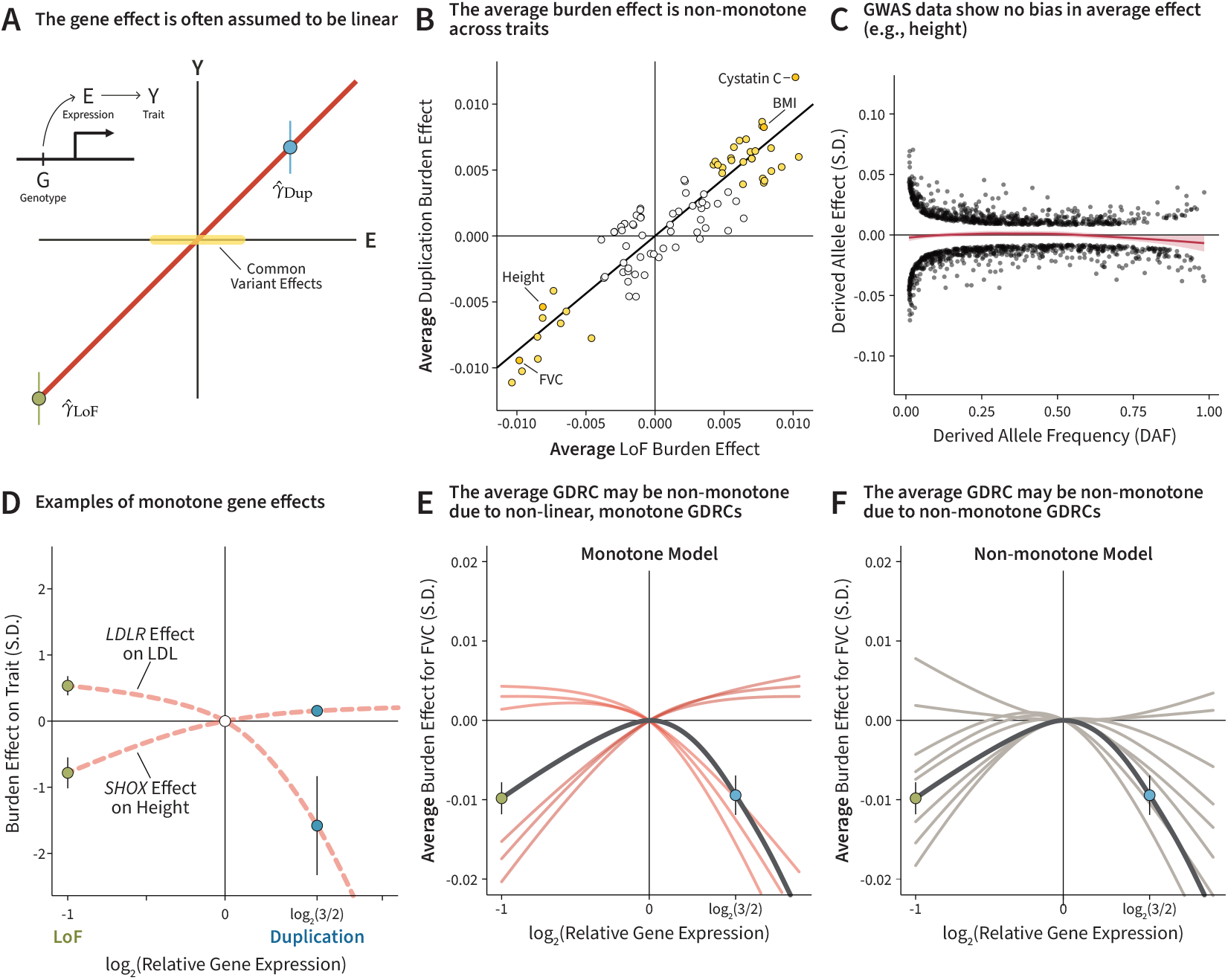
Explaining the paradoxical observations in burden data. **A**. The hypothetical relationship between gene dosage (E) and expected trait value (Y) is often assumed to be linear. LoF variants and duplications estimate the gene effect at different dosage values (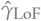 and 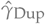). **B**. The average burden effect from LoF variants and duplications across traits. Yellow points represent significant effects from both variant classes (*p <* 0.05), and the regression line was estimated using total least squares (*β* = 0.87, *z* = 20.42). **C**. Conditionally independent GWAS top hits for height. The LOESS curve shows no bias across the frequency spectrum. For variants with a derived allele frequency less than 0.5, the proportion of trait-increasing alleles is 51% (95% CI: [0.48, 0.55]). For further discussion, refer to Appendix I.2. **D**. Burden estimates and 95% confidence intervals for two illustrative gene-trait pairs, with hypothetical GDRCs as dashed lines. *LDLR* encodes LDL receptor, which is a known negative regulator of LDL levels in blood, and acts by sequestering LDL out of the bloodstream [34, 35]. *SHOX* haploinsufficiency is associated with short stature, while increased dosage is associated with tall stature [36]. **E**. The first model explains the non-monotone average GDRC of FVC (with 95% confidence intervals) using monotone gene curves. **F**. The second model explains the non-monotone average GDRC of FVC (with 95% confidence intervals) using non-monotone gene curves.

However, the interpretation of each genome-wide burden effect estimator at the gene level is complicated by the strategy used to define an individual’s burden value. Previous studies have calculated various measures of genome-wide burden — counting the number of variants, the total length of CNVs, and the number of genes affected by CNVs. We can interpret these approximately as the effect of perturbing a randomly-chosen gene, assuming that the effect of CNVs occurs via perturbing individual genes, but this depends on other factors such as the local mutation rate of CNVs and the additivity of dosage effects across genes.

To estimate the actual effect of perturbing a randomly-chosen gene, we estimated the effect of each individual gene on a trait by using burden tests, and then averaged those estimates across all genes. We found that the estimated *average burden effect* for LoF variants was almost always in the same direction as that of duplications (Figure 1B), consistent with the genome-wide burden effects reported previously. Curiously, however, when we look at top hits from GWASs, we find no evidence for such an effect (Figures 1C and I.1), consistent with previous analyses [37].

Taken together, these observations provide contradictory information about how dosage is associated with traits. To explain these findings, we need to consider the gene-level heterogeneity present in dosage-response relationships. We envision the relationship between gene expression and trait as a gene dosage response curve (GDRC): a continuous curve describing how changes in gene expression affect the average trait value. The core idea is that if a gene is involved in the biology of a trait, then different baseline expression values for the gene will result in different trait values on average. A given point on the GDRC is the hypothetical mean trait value of individuals whose gene expression is set to a particular level. In other words, the GDRC characterizes the relationship between expression and trait value for all possible dosage perturbations. The GDRC extends prior work exploring the quantitative relationship between gene dosage and trait value [38–41]. Here, we propose that the GDRC acts as a conceptual framework to unify estimates of gene effects from LoF variants, duplications, and expression quantitative trait loci (eQTL), allowing us to understand the source of non-linearity in the data.

Averaging over genetic backgrounds, a particular variant will have a specific effect on expression and thus correspond to a specific point on the GDRC. We define the origin to be the mean trait and expression value in the population (Appendix A.4). A burden estimate of the LoF effect, 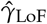, is an estimate of a point on the left tail of the curve, while a burden estimate of the duplication effect, 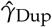, is an estimate of a point on the right tail of the curve (Figure 1A). The effect estimated by TWAS, 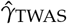, is the slope of a linear approximation to the GDRC in the region of expression levels spanned by common eQTL. While we visualize LoF variants and duplications as having specific dosage effects on expression (50% and 150% respectively), most of our analyses require only that they generally decrease or increase expression respectively (Appendix A.5).

Intuition suggests that GDRCs should be monotone, meaning that reducing and increasing expression should have opposite phenotypic effects (Figure 1D) [24, 25, 27, 42, 43]. The average burden effects that we computed (Figure 1B) can be interpreted as points on the average GDRC (aGDRC) obtained by averaging the GDRC of each gene for a given trait. If average LoF and duplication effects are in the same direction, as we observe for many traits, it would imply a non-monotone aGDRC. It therefore seems surprising that aggregating presumably monotone GDRCs would result in a non-monotone aGDRC.

We suggest two models that could contribute to non-monotone aGDRCs. In the *monotone model*, monotone gene-level GDRCs that are systematically buffered against one trait direction could average to a non-monotone aGDRC (Figure 1E). In the *non-monotone model*, some GDRCs are non-monotone, and among the non-monotone GDRCs it is more common to be non-monotone in a particular direction, driving the aGDRC to be non-monotone (Figure 1F). These models are not mutually exclusive and may both contribute to the non-monotone aGDRC observed for a trait.

### Top hits are consistent with monotone GDRCs, but also reveal non-monotone GDRCs

To understand the relative contribution of these models to aGDRCs, we started by looking at the results from the burden tests. Using a Bayesian analysis of the p-value distribution [44], we conservatively estimate that 5% of the gene-trait pairs have a true association (Appendix A.6). This suggests that even at current sample sizes, burden tests of LoF variants and duplications are relatively underpowered, and that it will be challenging to explain the non-monotone aGDRCs using only ascertained GDRCs.

We confirmed this intuition by performing an exome-wide analysis. We performed a stringent multiple testing correction on our burden tests, accounting for the number of genes, number of traits, and both types of tests conducted (Bonferroni p-value threshold of *p <* 2.87 *×* 10^−8^, equivalent to |*z*| *>* 5.55). At this threshold, we detected 443 LoF hits and 677 duplication hits. However, these hits were mostly non-overlapping. Only two hits overlapped, both of which indicated monotone GDRCs (Figure 2).

**Figure 2.**
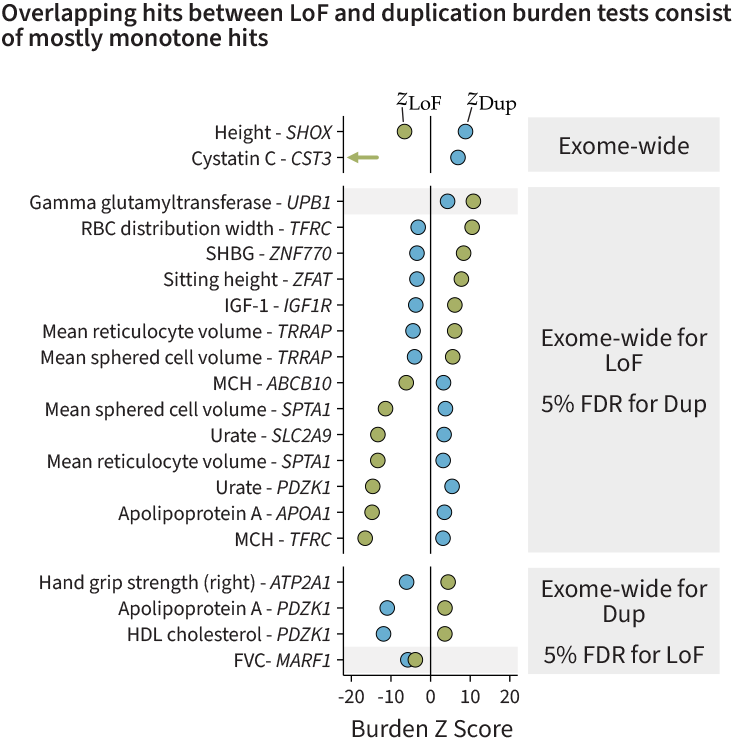
Top hits from burden tests. The Z scores from regression-based burden tests using LoF and duplication variants across various continuous traits. Only two genes had exome-wide significant LoF and duplication burden hits. To reduce the multiple-testing burden, we performed exome-wide ascertainment on LoF burden tests, and ascertained the duplications for the subset at a 5% false discovery rate (FDR). We performed the reciprocal test using the duplications. The green arrow represents a Z score smaller than -20. Non-monotone relationships are highlighted in grey.

To reduce the multiple testing burden, we used the 443 LoF hits as a discovery set and ascertained genes with significant duplication burden tests at a 5% false discovery rate (FDR). We also performed the reciprocal analysis using the duplication burden tests as a discovery set. Matching intuition, most top genes have monotone effects on traits (Figure 2), although we do observe two non-monotone GDRCs. At a 5% FDR, we expect 0.5 of the 20 discoveries to be a non-monotone false discovery, providing some evidence for their existence. However, it is difficult to know their relative proportions, let alone how much they contribute to the non-monotone aGDRCs, so we next developed quantitative methods to study monotonicity.

### Burden signal for traits is largely monotone

Due to the sparsity of the burden signal, we focused on quantitative measures that average signal across genes to explain the non-monotone aGDRCs (Figures 1E and 1F). First, we wanted to develop a notion of whether GDRCs generally tend to be monotone or not, beyond just considering the handful of significant hits.

We started with the observation that monotone GDRCs imply that LoF variants and duplications should have effects in opposite directions (Figure 3A), which we visualized by plotting the LoF burden effect against the duplication burden effect. If a duplication and an LoF variant have a large effect in the same direction, the GDRC must be non-monotone and the product of their effects would be positive. If the GDRC is monotone, then LoF variants and duplications will have opposite effects, and their product will be negative. We define *ϕ* to be a normalized version of the negative product of the effect sizes (Methods, Appendix B.2). *ϕ* is positive if genes with large effects on a trait tend to have monotone effects on average. Note that *ϕ* does not directly measure the proportion of monotone GDRCs, and its interpretation requires some care (Appendix F).

**Figure 3.**
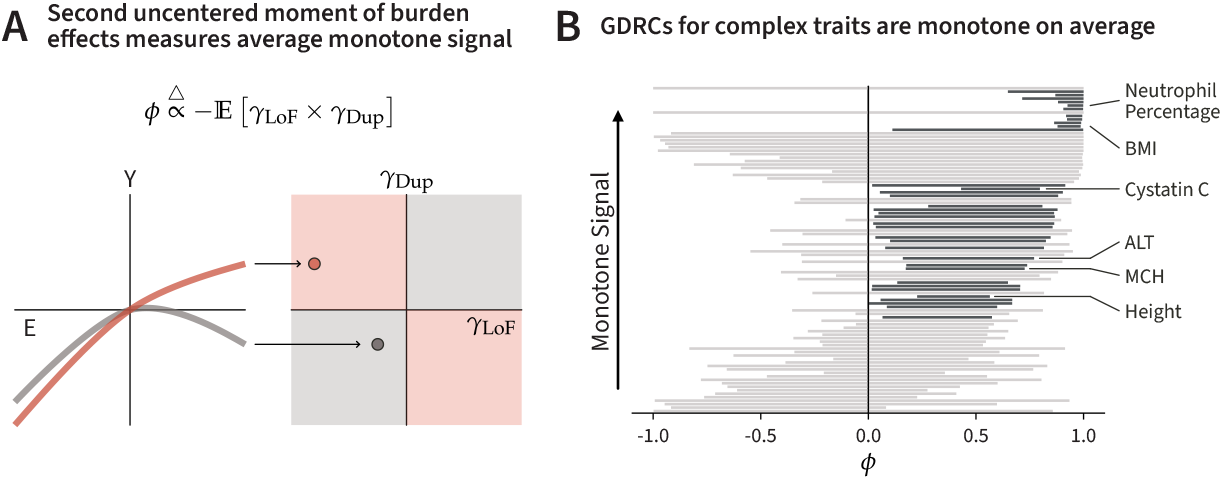
Monotonicity inference for traits. **A**. Monotone curves (red) and non-monotone curves (gray) fall in different quadrants on a plot of the duplication burden effect versus the LoF burden effect. A natural estimate to compare points in these quadrants is the second uncentered moment of the burden effect sizes. **B**. The bars represent 95% confidence intervals for *ϕ* for the traits in our analysis. The black bars are significantly non-zero (*p <* 0.05). The bars are ordered by 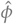.

We used a maximum likelihood approach to estimate *ϕ*. We performed extensive simulations from the individual-level genotype data to validate that our estimator, 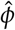, is relatively unbiased and well-calibrated (Appendix G.1.5). Across traits, we found that the estimated values of *ϕ* were generally positive (Figure 3B), suggesting that the burden signal is explained predominantly by monotone GDRCs. The same trend towards monotone signal was observed using a method-of-moments estimator for *ϕ* that makes fewer distributional assumptions (Appendix I.4). This matches our biological expectations of monotone gene-level effects, and is potentially consistent with the monotone model.

### A model for how non-monotone GDRCs arise

Our estimates of *ϕ* suggested that monotone GDRCs play a significant role in complex traits, but even among the small number significant hits for individual gene-trait pairs, we detected two non-monotone associations (Figure 2). While averaging across all genes indicates that the predominant signal is generally monotone (Figure 3), this does not preclude the possibility that non-monotone GDRCs, in aggregate, contribute to the non-monotone aGDRCs.

Indeed, prior evidence suggests that non-monotone GDRCs play some role in complex traits. One line of evidence comes from the analysis of genomic disorders caused by reciprocal CNVs, which are loci where both deletions and duplications are observed. Many reciprocal CNVs are known to induce the same phenotype [45, 46], suggesting that the effect of increasing or decreasing the dosage of the disorder-associated gene has the same direction of effect on the trait. Another line of evidence comes from careful analysis of CNVs in the UKB, where multiple loci show evidence of non-monotone associations with various complex traits [47, 48]. This suggests that, although challenging to detect at an exome-wide level, non-monotone GDRCs may still contribute to the non-monotone aGDRCs.

Before investigating genome-wide signal for non-monotone GDRCs, we wanted to develop intuition for how they might arise. Consider a gene that affects a trait via a single biological pathway (Figure 4A). Intuition about biological mechanisms suggests that each step along this pathway should be monotone, but possibly non-linear. For instance, protein levels are often buffered against large changes in gene expression [9], which results in a non-linear, but monotone response of protein levels to gene expression. Similarly, perturbations of the expression levels of transcription factors can result in non-linear but mostly monotone changes in the expression of their targets [40, 41]. As the dosage effect percolates through a pathway towards the focal trait, these curves compose with one another to form the relationship between gene dosage and trait. If each curve along the pathway is monotone, then the overall relationship between gene expression and trait is mathematically guaranteed to also be monotone (Figure 4A, Appendix A.7).

**Figure 4.**
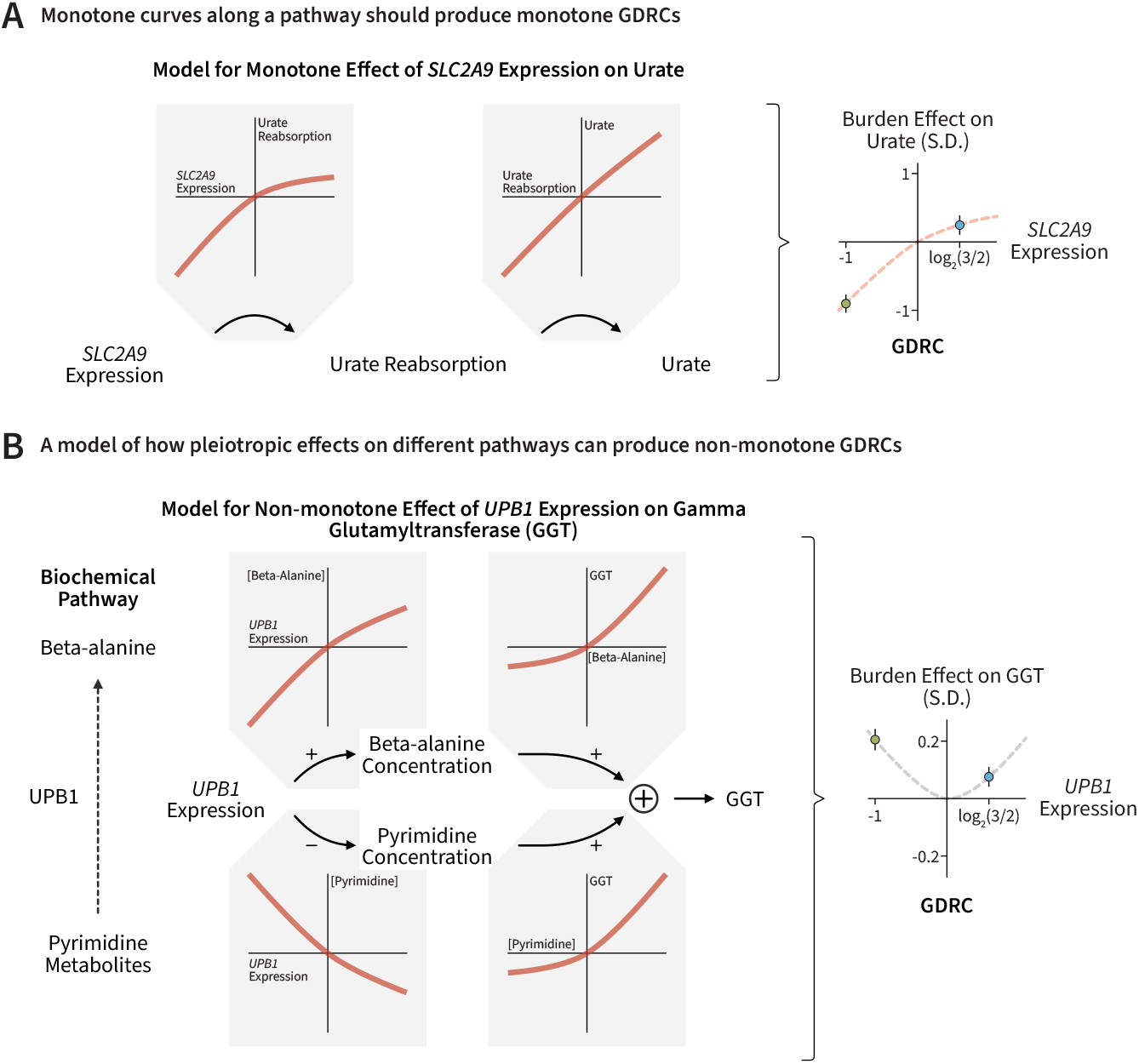
Model for non-monotone GDRCs. **A**. A simple model on the left, composed of monotone curves at each step, can explain the monotone GDRC observed for *SLC2A9* and urate levels. *SLC2A9* encodes a kidney transporter responsible for reabsorbing urate back into the blood. Thus, increased *SLC2A9* expression may contribute to increased urate reabsorption, which in turn can increase the levels of urate in blood. **B**. In our model, the effect of *UPB1* propagates through two separate paths to affect gamma glutamyltransferase (GGT) levels, which may explain the non-monotone GDRC. The first path, similar to our monotone example, consists of two increasing steps (denoted by + symbols above the arrows). The second path consists of one increasing and one decreasing step (denoted by the − symbol below the arrow). The outcome of the paths is summed in our conceptual model, resulting in a non-monotonicity that might explain the observed data.

One example of a monotone association in our data is that of *SLC2A9* expression and urate levels (Figure 2). *SLC2A9* encodes a transporter expressed in the kidneys that is responsible for reabsorbing urate into the blood by transporting urate from the kidney into the bloodstream [49, 50]. A simple model of the observed monotone GDRC is that *SLC2A9* expression has a monotone effect on urate reabsorption, which in turn has a monotone effect on urate levels in blood (Figure 4A) [51]. The composition of these two monotone steps can explain the monotone GDRC we observe in the data.

However, non-monotone GDRCs can arise if a gene’s effects flow through two or more pathways (Figure 4B). We propose a simple model for the non-monotone relationship between *UPB1* expression and gamma glutamyltransferase (GGT) levels in blood (Figure 2). Both LoF variants and duplications in *UPB1* increase GGT levels on average (Figure 4B). GGT is an enzyme found in the membrane of epithelial cells in various tissues, particularly in the hepatobiliary system, and acts as a readout of oxidative stress [52–54]. *UPB1* encodes an enzyme involved in the breakdown of pyrimidines in the liver to downstream byproducts such as beta-alanine. Both excess pyrimidine metabolites and excess beta-alanine can result in toxicity and increased oxidative stress [55–57]. Thus, we propose that increased *UPB1* expression has two monotone effects: it decreases the concentration of pyrimidines, and increases the concentration of beta-alanine, which both can contribute to increased GGT levels by increasing oxidative stress. The total effect of these two pathways could explain the non-monotone GDRC observed between *UPB1* and GGT levels (Figure 4B). Generalizing these biological principles, non-monotone GDRCs can arise whenever the effect of a gene propagates through multiple pathways to affect the trait (Figure 4B).

### Genes are buffered against one trait direction

One interpretation of the GDRCs in the monotone and non-monotone models (Figures 1E and 1F) is that traits are more likely to be perturbed in one direction than the other regardless of the direction of expression perturbation. Therefore, this is a form of buffering that occurs at the trait level, which we refer to as *trait buffering*. We endeavored to quantify how much monotone and non-monotone GDRCs contributed to trait buffering and the non-monotone aGDRCs.

When plotting duplication burden effects against LoF burden effects, genes experiencing trait buffering preferentially map to a region that is away from a diagonal line (Figure 5A). The diagonal line represents perfectly linear GDRCs (Figure D.1). Points above the diagonal line correspond to GDRCs that are buffered against negative trait values, and points below the diagonal line correspond to GDRCs buffered against positive trait values.

**Figure 5.**
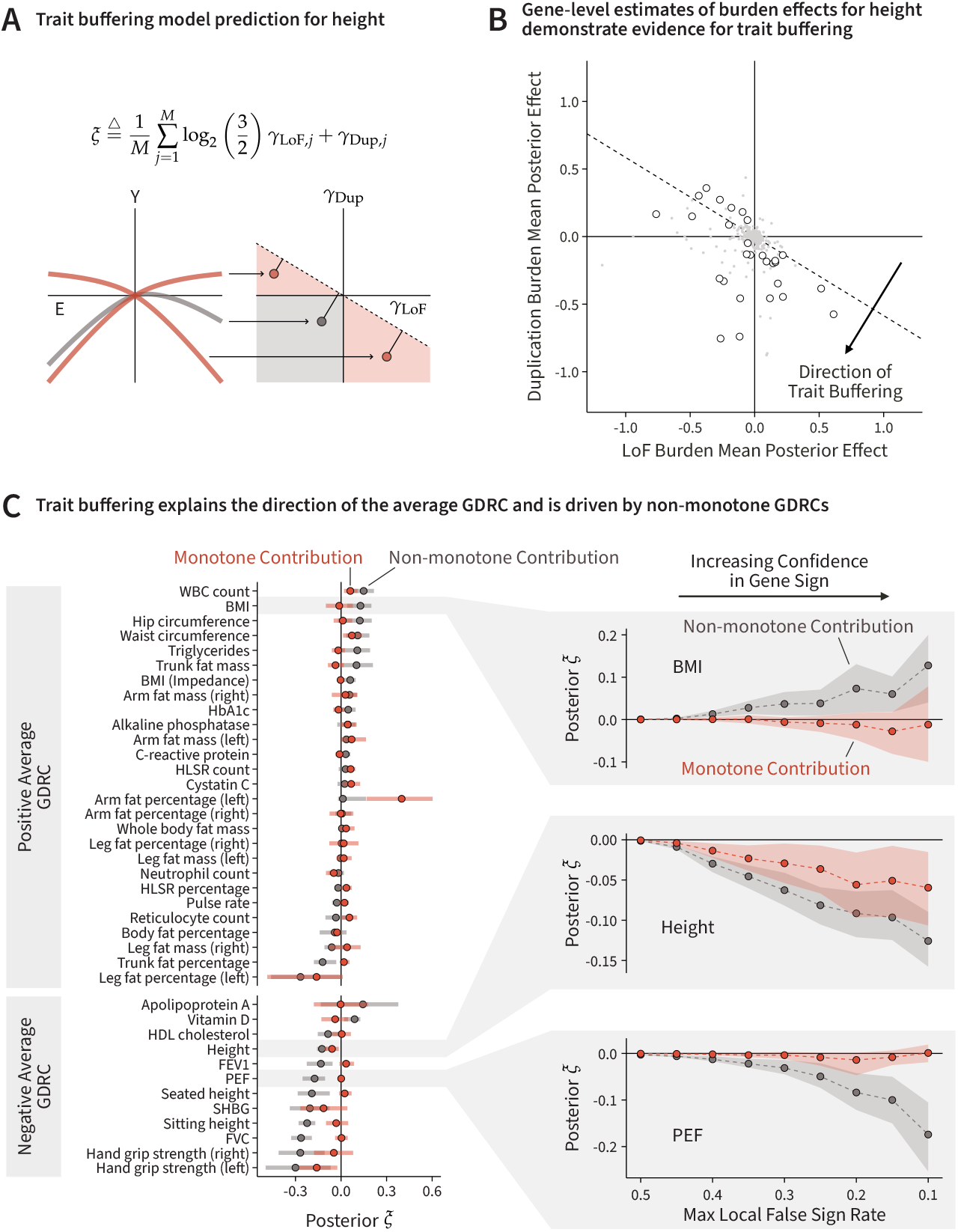
Trait buffering as a model for GDRC architecture. **A**. After combining both models of GDRCs contributing to the non-monotone aGDRC, the GDRCs fall below a diagonal line for traits with negative average burden effects such as height. The average LoF and duplication burden effects will be in the direction of trait buffering. *ξ* measures the average signed deviation from the diagonal line, represented by the lines orthogonal to the diagonal line. **B**. Estimates of the latent burden effects show, visually, that height is consistent with trait buffering. Each point represents a gene. The large circles represent genes where the local false sign rate for both LoF and duplication burden effects is less than 10%. **C**. (Left) The posterior estimates of trait buffering, *ξ*, decomposed into contributions from non-monotone and monotone GDRCs. These estimates are derived from genes with a local false sign rate of less than 10% for the LoF and duplication burden effects. (Right) The estimates of the components remains non-zero with increasing confidence in the signs of the gene-level effects.

To develop some preliminary intuition about the trait buffering model, we estimated *γ*_LoF_ and *γ*_Dup_ for each gene. Since burden effect estimates are noisy for individual genes, we performed Bayesian inference using a flexible multivariate adaptive shrinkage (MASH) prior [58], which models the joint distribution of *γ*_LoF_ and *γ*_Dup_ as a mixture of bivariate normal distributions that is learned from the data. We used posterior samples of *γ*_LoF_ and *γ*_Dup_ for each gene to better understand how genes might contribute to trait buffering. As an example, the posterior means of *γ*_LoF_ and *γ*_Dup_ for height are displayed in Figure 5B, and further examples are provided in Appendix I.7. Both monotone and non-monotone GDRCs are predominantly below the diagonal, concordant with the direction of the aGDRCs for height.

To test this more formally, we developed a statistic, *ξ*, to measure trait buffering. *ξ* aggregates the signed strength of deviation from the diagonal line across all GDRCs (Appendix D). We calculate the statistic using posterior samples of *γ*_LoF_ and *γ*_Dup_.

The sign of *ξ* indicates which trait direction is being buffered against. For example, based on their aGDRCs, we would predict that height should have a negative value of *ξ* and cystatin C should have a positive value of *ξ*. If GDRCs are not preferentially buffered against either direction of a trait, our statistic should be close to zero. Indeed, this measure of trait buffering showed strong concordance with the direction of the non-monotone aGDRCs (Figure I.8).

To understand the extent to which our proposed models (Figures 1E and 1F) contributed to trait buffering, we decomposed *ξ* into the contribution from non-monotone and monotone GDRCs (Methods, Appendix D), and estimated these components from genes where we were relatively confident in the sign of effect using a local false sign rate filter. These measures revealed that both monotone and non-monotone curves contribute to trait buffering (Figure 5C). These patterns persist as we increase the confidence in the sign of the gene’s effect (Figure 5C). We performed extensive simulations to demonstrate how these component estimates behave under various extreme scenarios (Appendix G.2). Juxtaposed with these simulations, our results for certain traits (*e*.*g*., FVC, PEF, FEV1, BMI, height) show a clear signature of non-monotone genes having a substantial contribution to trait buffering. Ultimately, genes with both monotone and non-monotone GDRCs (Figures 1E and 1F) likely contribute to observed non-monotone aGDRCs.

### A dysregulation phenotype may be upstream of many complex traits

To recapitulate, we found evidence for two models contributing to the observed average burden effects. These models imply that GDRCs for complex traits are organized in a manner that results in trait buffering.

Two observations initially seem to imply that the complex traits with non-monotone aGDRCs are under directional selection. First, the GDRCs for these traits are buffered against a particular trait direction. Second, non-monotone aGDRCs suggest that any variant should on average have a non-zero effect in the same direction regardless of whether the variant increases or decreases expression.

To see why these observations are signatures of directional selection, we consider how the average expression of a given gene will evolve over time in response to directional selection. Suppose a gene affects a trait where higher trait values are selected against (Figure 6A). Then, selection will tune the expression of the gene until the trait is minimized, resulting in a non-monotone GDRC where both deletions and duplications increase the trait. Thus, the non-monotone aGDRC and trait buffering in the direction of the average burden effects suggests that the GDRCs of these complex traits may be shaped by directional selection.

**Figure 6.**
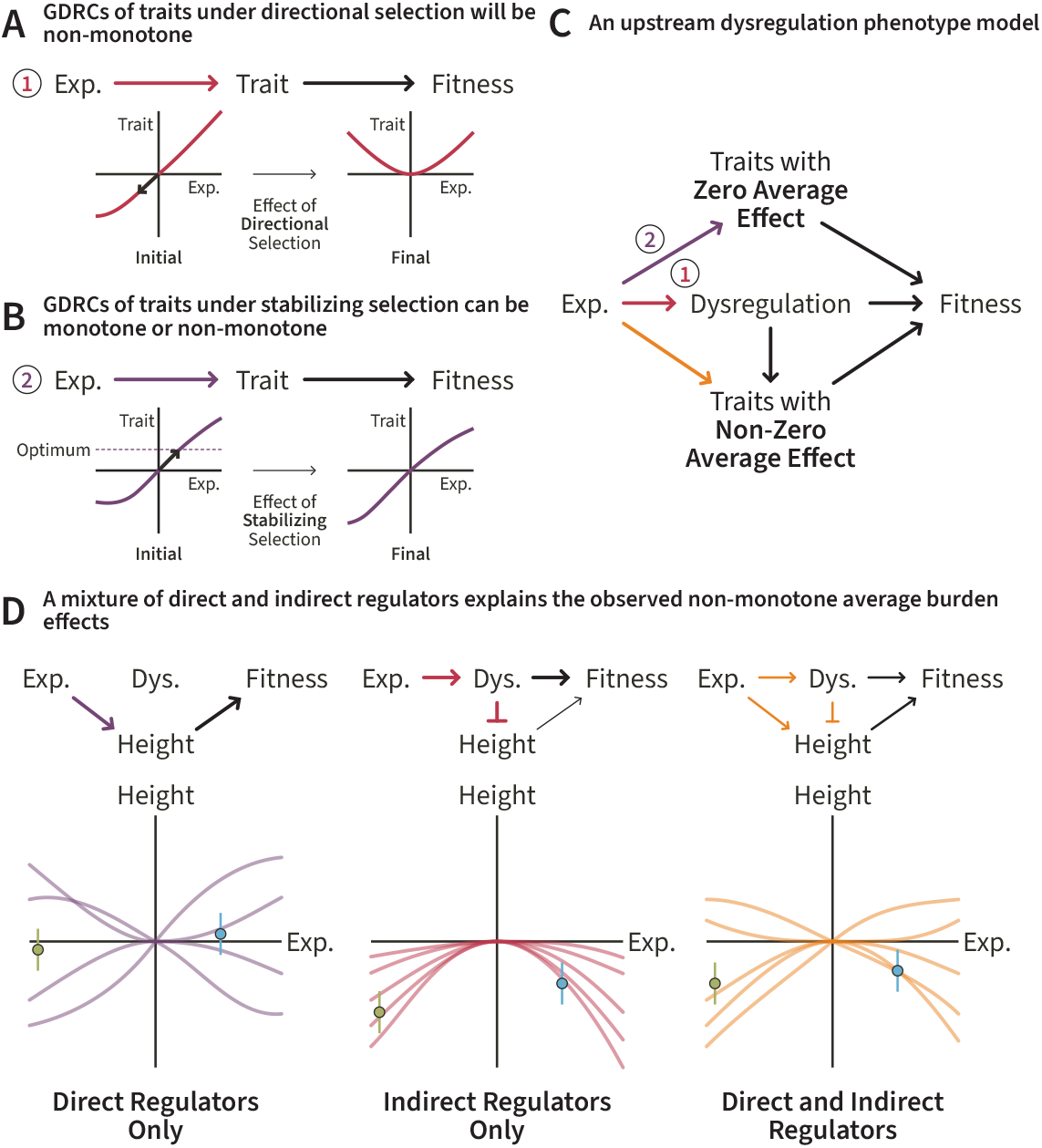
A theoretical model to explain trait buffering. **A**. GDRCs of traits under directional selection will become non-monotone over evolutionary time (Exp. = expression). **B**. GDRCs of traits under stabilizing selection will have no preference for GDRC shape (Exp. = expression). **C**. In our model, traits with non-monotone aGDRCs have an upstream dysregulation phenotype, which we hypothesize is under directional selection (Exp. = expression). **D**. Example GDRCs of genes that follow specific paths to affect the focal trait (for example, height) under our theoretical model (Exp. = expression, Dys. = dysregulation).

However, prior analyses using GWAS data for many of these traits suggest that they are under stabilizing selection, not directional selection [59–63]. Additionally, we do not see a non-zero average effect in GWAS data (Figure 1C, Appendix I.2), inconsistent with directional selection.

Stabilizing selection affects GDRCs differently than directional selection. Suppose a gene affects a trait under stabilizing selection (Figure 6B). Stabilizing selection acts to maintain an optimum trait value, and will tune the expression of the gene to reach the optimum trait value. Unlike in the case of directional selection, the local shape of the GDRC around the optimum trait value is not restricted to minima. In this case, we do not expect any particular GDRC shape to dominate, and we do not expect LoF variants or duplications to be biased in a particular direction.

To explain the incongruous non-zero average burden effects and zero average GWAS effect, we propose that traits with non-monotone aGDRCs are downstream of an unobserved trait under directional selection that we refer to as “dysregulation” (Figure 6C). If the focal trait is downstream of dysregulation, the GDRC of the gene for the focal trait may be shaped by both forms of selection depending on if the gene is a direct or indirect regulator of the focal trait (Figure 6D). For genes that act primarily on the trait via dysregulation, the GDRCs will be non-monotone and in the direction of increasing dysregulation. For genes that directly modulate the focal trait, we expect no particular preference for monotone or non-monotone GDRCs. Genes that act on the focal trait both directly and via dysregulation explain the monotone curves that experience trait buffering.

Overall, this model explains the non-monotone average effects that we observe in the data. Depending on the balance between direct regulation or indirect regulation via dysregulation, we can still have predominantly monotone GDRCs but have a consistent bias toward one direction of the focal trait.

Our model also explains why the non-zero average effect is apparent in LoF variants and duplications across various traits, but is not discernible in GWAS effect sizes. Consider two variants that have the exact same effect on the focal trait. One variant affects the trait directly, while the other affects the trait via dysregulation. Under our model, the second variant that acts via dysregulation can only affect the focal trait in one direction because it has a non-monotone GDRC, while the first variant has no such restriction. Both variants will experience the exact same fitness consequences from stabilizing selection on the trait. However, the variant acting via dysregulation will experience additional fitness consequences. Thus, holding all other factors equal, the variant acting via dysregulation will have a lower allele frequency and be more challenging to detect in a GWAS, thus making it less likely to contribute to a non-zero average effect.

We call this unobserved latent phenotype “dysregulation” because the downstream traits with large non-monotone average burden effects are readouts of general dysfunction. The traits with significant non-monotone average effects (Table I.1) include measures of organ dysfunction such as alkaline phosphatase, cystatin C, and C-reactive protein, which all have positive average LoF and duplication effects. Put another way, both deletion or duplication of a random gene tend to increase disease risk and measures of organ dysfunction. Similarly, measures of lung capacity and muscle strength have negative average LoF and duplication effects. Overall, the average effect is often in the direction of dysregulation across the continuous traits we studied.

Recent analyses of predicted biological age in the UKB also support the presence of a dysregulation phenotype [64, 65]. Age is associated with general dysregulation and disease [66]. Strong predictors of biological age in published predictive models include FEV1, cystatin C, HbA1c, alkaline phosphatase, and hand grip strength [64, 65], suggesting that these continuous traits with large non-monotone average effects may act as readouts of dysregulation.

## Discussion

In this study, we interpreted dosage-perturbing rare variants through the lens of the GDRC. Our analysis suggests that both monotone and non-monotone GDRCs contribute to the concordant average burden effects of LoF and duplication variants. This results in GDRCs that are buffered against one trait direction. We hypothesized that this may be explained by the effect of genes on an upstream dysregulation phenotype, resulting in directional selection shaping GDRCs.

### Implications of the observed characteristics of gene dosage response curves

In a “linear world” where dosage has a linear relationship with a trait, ascertaining any point on the GDRC along the dosage spectrum would carry all the information about the gene’s effect on the trait. However, biology is inherently non-linear, and our analysis shows that these assumptions break down for variants with large effects. This has significant implications for drug design. If the GDRC of the target gene is non-monotone, perturbation in either direction may only increase disease risk. This occasional lack of concordance between LoF and duplication burden effects motivates the use of assays that test both ends of the dosage spectrum [67]. The shape of the GDRC implied by the burden estimates can also be a useful prior for fine-mapping causal variants [68] and linking variants to genes [69–73].

The assumption of linearity in TWAS [5, 6, 74] and Mendelian randomization [75] approaches may cause estimated effect sizes to be attenuated towards zero for non-monotone genes. The ability of TWAS to recover trait-associated genes with non-linear GDRCs needs to be explored further. Yet, TWAS-type methods would be invaluable for understanding the behavior of the GDRC in the physiological range of expression, thus motivating the continued development of such methods.

### Towards the inference of gene-level curves across tissues and contexts

Moving from genetic associations towards mechanistic insights for complex traits remains challenging. The GDRC captures a large fraction of the biological insights that we want to extract from association studies. Although inference of the GDRC may be challenging, it is worth considering what aspects of the GDRC we can estimate using population-level and experimental approaches.

In principle, each tissue or cell type has its own specific GDRC for a gene-trait pair. In our approach, we focused on the global GDRC — which we could interpret as a weighted sum of tissue-specific GDRCs — by using variants that affect all tissues and contexts to understand the aggregate effect of dosage perturbation on trait. Modern approaches that estimate the effect of genes in specific tissues or cell types, such as TWASs, can be thought of as estimating or approximating aspects of tissue-specific GDRCs.

Even with methodological advances, it is unlikely that we will be able to infer all points of the GDRC of a gene with population-level data alone. The effects of selection make it challenging to detect variants in genes with large effects on traits [76, 77]. This suggests that the ability to effectively infer the GDRC will vary across the genome, and likely be most challenging for genes that are particularly important to the biology of traits.

These limitations of population-level data suggest that sequence-to-expression models [78–81] and experimental approaches [82–84] will play a critical role in fleshing out the GDRC for key genes. Missense variants, which in aggregate have a measurable effect on complex traits [31], are challenging to integrate into the GDRC framework. Accurate computational effect prediction [85] or experimental approaches such as deep mutational scanning [86] are critical for translating the effect of missense variants into an “effective dosage”, which would allow them to be placed on the GDRC. Similarly, for common variants, we are restricted to using dosage effect estimates from eQTL studies in tissues that have been assayed, which are often post-mortem and from adult donors [87]. However, we cannot feasibly assay all tissues and cell types in all contexts. Thus, predicting common variant effects from sequence and multi-omic context or using experimental approaches such as massively parallel reporter assays [83] will be necessary to estimate full GDRCs. New techniques are also being developed to modulate dosage directly to measure molecular or cellular phenotypes [41, 67, 88–91], which can provide *in vitro* measurements of GDRCs.

In summary, we have explored the nature of the relationship between gene dosage and trait for various complex traits. Our final model explains the initial observation of non-monotone average effects and provides intuition for how these relationships may be shaped by natural selection. The insights about GDRCs underlying complex traits will be informative for various downstream applications, including therapeutics and methods development.

## Methods

The goal of our data analysis approach was to estimate burden summary statistics for LoF variants, whole-gene deletions, whole-gene duplications, and partial deletions using a consistent set of decisions and tools. This consistency between the different burden tests allowed us to compare effect sizes across the variant classes. We additionally performed burden tests using synonymous variants as a negative control to estimate any residual confounding (Appendix H.1). We improved upon previous LoF burden approaches by using variable frequency filters based on gene constraint (Appendix H.1.3). We also improved upon prior duplication burden tests in the UKB [23–25, 27] by using copy number calls from WGS data rather than microarray data. CNV calls from WGS are more accurate, have higher resolution of breakpoints, and can better detect rare CNVs [92].

To make our approach amenable to researchers without access to individual level data, we modeled the summary statistics rather than the underlying genotype data. We use the regression with summary statistics (RSS) likelihood [93], which provides a likelihood model for association test summary statistics. To our knowledge, this is the first use of the RSS likelihood for burden regression. To demonstrate its applicability and correctness, we performed extensive simulations, which are cataloged in Appendix G.

### Inclusion and Ethics

This study relies on data provided by participants in the United Kingdom to the UKB. The UKB is a large biomedical database that provides non-exclusive access to individual-level data. Primary administration of the UKB is handled by groups within the United Kingdom, and a local ethics board provides oversight over the collection of data. The UKB has approval from the North West Multi-centre Research Ethics Committee as a Research Tissue Bank (RTB) approval. This approval means that researchers do not require separate ethical clearance and can operate under the RTB approval.

The main burden tests performed in this paper do not subset individuals based on measures of genetic similarity. However, to compare the effects of confounding and stratification between our study and previous studies, we subset individuals based on self-identification as “White British” or “non-British White” and genetic principal components (PCs) for a small set of burden tests presented in Appendix H.1.1. Results and summary statistics from all analyses presented here are provided as a resource to the scientific community (Data Availability).

### Statistics and Reproducibility

Genome-wide burden tests performed in this study follow quality control standards set by prior analyses, with additional quality control measures discussed in Appendix H. Results from parameter inference are paired with appropriate measures of uncertainty. Inclusion and exclusion criteria during analysis are discussed wherever appropriate. We provide code to replicate our analysis (Code Availability).

### Individuals

We followed a protocol similar to a prior analysis of rare LoF variants in the UKB [31]. We use all 460K individuals with WES data in the UKB data. This is slightly larger than the fraction used by Backman et al., who focused on 430K individuals with genetic similarity to the EUR “superpopulation” as defined by the 1000 Genomes Project. Although relatedness and population structure can introduce environmental confounding, our pilot analyses with synonymous variants (Appendix H.1.2) indicated minimal levels of uncorrected confounding.

### Phenotypes

We curated 410 continuous phenotypes in the UKB on which to perform burden tests. We filtered the phenotypes used for the main analysis in the paper as follows. We required phenotypes to have measurements in at least 40K individuals. We excluded any traits where more than 50% of the instances match the mode of the trait to remove quasi-categorical traits, which is more relaxed compared to Backman et al., who exclude traits with a cutoff of 20%. To perform inference, we chose traits where at least 7,500 genes had gene-level burden test estimates for both LoF variants and duplications. Finally, we required that either the mean squared effect size of the LoF burden tests or the duplications was nominally significant at the *α* = 0.05 level for each trait. This removes traits where there is not enough signal to perform inference. After filtering, 94 traits remained and were used throughout the main analysis of the paper.

For phenotypes that were collected more than once in returning visits, we use the first instance of the observation for each individual. Backman et al. took the mean across such instances, but we believe that this would reduce the noise for some individuals more than others in a not completely at random fashion. It would also make it challenging to interpret the relationship between mean phenotype and covariates that are observed at specific instances, such as age. Some of our traits had arrayed measurements, but they were all taken in the same visit so we decided to use the mean value across the arrayed items.

Separately, we acquired access to the full 500K baseline metabolomics data from the nuclear magnetic resonance (NMR) metabolomics data set generated by Nightingale Health PLC [94].

### Genotypes

We included 15 genotyping PCs, estimated previously in the UKB [95]. Backman et al. performed ancestry-specific analyses and included 10 genotyping PCs for each ancestry. Since we used all 460K WES samples, we included an additional 5 PCs to address potential stratification. Our analysis of synonymous variants (Appendix H.1.1) supported this choice.

Following the lead of Backman et al., we included 20 rare variant PCs, by which we mean PCs derived from rare WES variants. We subsampled 300K variants uniformly at random from the rare fraction, which we defined as variants with minor allele count (MAC) greater than 20 and minor allele frequency (MAF) less than 1%. We used an approximate principal components analysis (PCA) algorithm implemented in FlashPCA2 [96].

We restricted our burden analysis to LoF sites with MAF less than 1%. High-confidence LoF sites were annotated using Ensembl’s Variant Effect Predictor [97] with the LOFTEE plugin [98]. LOFTEE filters remove annotated LoF sites that might escape nonsense-mediated decay (NMD). Instead of using progressively stricter MAF cutoffs as in Backman et al., we use all LoF sites with MAF less than 1% but require that the misannotation probability be less than 10%. The misannotation probability [77] takes into account both the frequency of the site and the constraint experienced by the gene to determine a posterior probability of being misannotated as a LoF. These probabilities were previously reported for LoF-introducing single nucleotide polymorphisms (SNPs) for genes with estimated *s*_het_ values [77]. A small number of genes do not have *s*_het_ estimates, and not all variants in the WES data were SNPs. For genes with missing *s*_het_ values, *s*_het_ was imputed using the mean *s*_het_ value. Then, for all LoF variants missing a misannotation probability, misannotation probabilities were imputed using k-nearest neighbors (kNN) regression with 10 nearest neighbors as determined by allele frequency and *s*_het_. A 10% cutoff on this probability retains 96.46% of variants. We demonstrated that relative to standard frequency cutoffs, using a misannotation probability cutoff increases the signal in the burden tests without increasing the size of standard errors (Appendix H.1.3).

CNVs were previously called in the UKB using Illumina’s DRAGEN software [99, 100]. A deletion was defined as any loss that affected the entire gene body with an additional 1 Kbp flanking region. A duplication was defined as any gain that affected the entire gene body with an additional 1 Kbp flanking region. Any deletions that affected only part of the gene body were categorized as potential loss-of-function (pLoF) alleles. For deletions and pLoF variants, the alternative allele coded the number of copy loss events. For duplications, a copy number of two was coded as the reference homozygote and a copy number greater than two was coded as a heterozygote. We observed that some samples had a large amount of genome-wide copy gain or loss, which is likely a genotyping error (Appendix H.2). To account for this, we removed any sample with greater than 10 Mbp of deleted or duplicated genome sequence.

We used GENCODE version 39 to define gene intervals [101]. We excluded genes on the Y chromosome and the mitochondrial genome.

### Burden Tests

We used REGENIE to perform gene-level burden tests [33]. For the whole-genome regression, which is the first step in REGENIE, SNPs from the genotyping array were pruned with a 1000 variant sliding window with 100 variant shifts and an *R*^2^ threshold of 0.9. SNPs were then filtered to have MAF > 1%, genotype missingness < 10%, and Hardy-Weinberg equilibrium (HWE) test p-value > 10^−15^.

For LoF burden tests, we used the following covariates: age, sex, age-by-sex, age squared, 15 genotyping PCs, 20 rare variant PCs, and WES batch. For the duplication, deletion, or pLoF burden tests, we used the following covariates: age, sex, age-by-sex, age squared, 15 genotyping PCs, 20 rare variant PCs, and WGS batch. When analyzing NMR metabolites, we additionally included the spectrometer and processing batch as covariates.

After whole-genome regression, we used the second step of REGENIE to perform burden tests. Phenotypes were rank-inverse-normal transformed in both the first and second step. The same covariates were used in both steps.

### Monotonicity Model

We modeled the standardized trait of interest **Y** in *N* individuals as

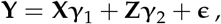

where **X** and **Z** are the burden genotypes from LoF variants and duplications respectively. The error ***ϵ*** was assumed to be drawn from a normal distribution. Polygenic signal, population stratification, and other sources of confounding were accounted for when performing the burden tests by including covariates. We built a hierarchical model by specifying a distribution on the latent effect sizes of the *j*th gene:

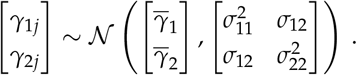

This allowed us to model the quantities of interest. The average effects of deletions and duplications are represented by 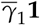 and 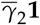 respectively. We defined the monotonicity, *ϕ ∈* [−1, 1], to be a quantity that is proportional to the negative uncentered second moment of the latent effect sizes,

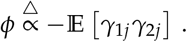

Thus, if genes with large effects tend to have opposite signs on average, *ϕ* will be closer to 1. Similarly, if genes with large effects tend to have the same sign on average, *ϕ* will be closer to −1. The exact definition of the parameter is provided in Appendix B.2.

The observed burden effect sizes for the *j*th gene, 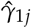 and 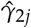, are noisy estimators of the underlying latent effect size. Furthermore, due to correlation between duplication burden genotypes, 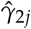 is not an unbiased estimator of *γ*_2*j*_ because the observed effect size is inflated by the effect of nearby genes. To account for these concerns, we used the RSS likelihood [93], which formalizes an approximation for regression-based summary statistics that is commonly used in statistical genetics [102, 103]. An extensive description of the model is provided in Appendix B.

We developed unbiased estimators of 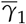 and 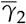. Because *ϕ* is a complex function of the parameters of the prior, it was challenging to derive unbiased estimators. Instead, we opted to use maximum likelihood estimation for *ϕ*, which guarantees consistency. Inference and simulations under this model are described in Appendices C and G.1 respectively.

### Trait Buffering Model

Since the monotonicity, *ϕ*, is a function of first and second moments of the latent effect sizes, we used a normal distribution as the prior. However, our buffering model proposed a more complex hypothesis about the distribution of the latent effect sizes, which required a more flexible prior over the latent effect sizes.

To address this, we modified our monotonicity model to use the MASH model prior [58], which is a flexible multivariate prior. MASH uses a mixture of *K* multivariate normal distributions to model the latent space:

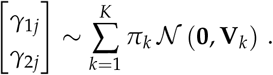

Since the true latent distribution is unknown, MASH uses a grid of fixed covariance matrices and learns the mixture weights ***π*** using empirical Bayes. In its original implementation, MASH assumes independence between observed samples, allowing for efficient optimization. Because the RSS likelihood necessarily induces dependencies between the observations via correlation between the burden genotypes, we implemented a stochastic approximation expectation maximization (SAEM) algorithm to optimize ***π*** [104, 105]. Alternative approaches to this model that use variational inference have been proposed [106–108].

As we defined it, the amount of trait buffering can be estimated by taking the dot product of the latent effect size with the normal vector of the hyperplane separating the two types of buffering, which we explain further in Appendix D.2. We define the measure of trait buffering as

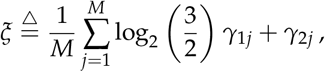

where *M* is the number of genes. We perform inference using the posterior distribution for *ξ* given the observed summary statistics. To determine how much is contributed to *ξ* by monotone or non-monotone genes, we defined

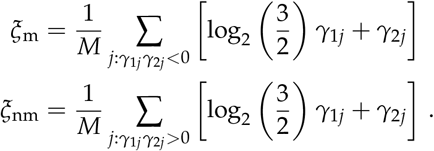

The intuition here is that *ξ*_m_ sums up contributions from monotone genes (where *γ*_1*j*_*γ*_2*j*_ *<* 0), and *ξ*_nm_ sums up contributions from non-monotone gene (where *γ*_1*j*_*γ*_2*j*_ *>* 0). Note that, by construction,

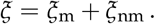

*ξ* is zero under the null model where GDRCs are equally likely to be buffered against increasing or decreasing the trait. The details of the model and inference can be found in Appendices D and E respectively.

### Data Analysis

Throughout, we use unbiased maximum likelihood estimates (MLEs) for the duplication burden estimates when they are displayed in the main text, which account for correlations with neighboring genes.

In Figure 1B, we used total least squares to fit a regression line with no intercept. Weights in the regression corresponded to the inverse variance of the estimates. To provide a better fit, we used multiple initial parameter values. We retrieved GWAS summary statistics from the UKB to generate trumpet plots (Figures 1C and I.1). The collection and analysis of these data is described in Appendix I.2. We fit a locally estimated scatterplot smoothing (LOESS) curve to visualize the conditional mean of the derived allele effect given the derived allele frequency.

For the analysis presented in Figure 5, we use the local false sign rate (LFSR) [109]. For a given burden test effect size estimate for a gene, a rate of less than 10% implies that more than 90% of the posterior density is on one side of zero, indicating confidence in the sign of the effect.

## Data Availability

All genetic and health data were acquired from the UK Biobank, a biomedical database containing information from half a million UK participants (https://www.ukbiobank.ac.uk/). Data for genetic association analysis of continuous traits were acquired under application 52374. Data for genetic association analysis of metabolite traits were acquired under application 30418. These data are available upon application to the UK Biobank. All summary statistics generated from genetic association analyses and other processed data files are deposited in Zenodo at https://doi.org/10.5281/zenodo.16800547.

## Code Availability

Scripts used to process and analyze data are deposited in Zenodo at https://doi.org/10.5281/zenodo.16799580. Scripts were executed either on the UK Biobank Research Analysis Platform (https://ukbiobank.dnanexus.com/) or on the Sherlock High-Performance Computing Cluster managed by the Stanford Research Computing Center (https://www.sherlock.stanford.edu/).

## Acknowledgments

We thank the members of the Pritchard lab for their feedback on this project and manuscript. In addition, we thank Yi Ding and Zeyun Lu (Gusev Lab) for reading over and providing feedback on the initial draft. We thank Luke O’Connor and an anonymous reviewer for their careful reading and feedback on our initial submission, which helped improve the paper during peer review. This research has been conducted using data from the UK Biobank (https://www.ukbiobank.ac.uk/), a major biomedical database. We thank all participants and researchers involved in the UK Biobank for their contribution, and Nightingale Health PLC for early access to the NMR metabolomics data. Computing for this project was performed on the Sherlock High-Performance Computing Cluster. We thank Stanford University and the Stanford Research Computing Center for providing computational resources and support that contributed to this research. N.M. was supported by a National Science Foundation Graduate Research Fellowship and Stanford’s Knight-Hennessy Scholars Program. C.J.S. was supported by Stanford’s Knight-Hennessy Scholars Program and the Stanford Center for Computational, Evolutionary and Human Genomics. H.Z. was supported by Stanford Biology Department’s graduate student assistantship. T.G. was supported by Stanford’s Knight-Hennessy Scholars Program. This work was supported by the National Institutes of Health (R01HG011432, R01HG008140, and R01HG014005 to J.K.P.).

## Author Contributions

**N.M**. Methodology, Software, Formal Analysis, Data Curation, Writing – Original Draft, Visualization. **C.J.S**. Methodology, Software, Writing – Original Draft, Visualization. **H.Z**. Methodology, Software, Writing – Original Draft, Visualization. **T.G**. Methodology, Software, Writing – Original Draft. **J.P.S**. Conceptualization, Methodology, Writing – Original Draft, Supervision. **J.K.P**. Conceptualization, Methodology, Writing – Original Draft, Supervision, Project Administration, Funding Acquisition.

## A Gene Dosage Response Curves

### A.1 Glossary of Terms

**Table A.1.** A table containing terms and associated symbols used in this paper.

| Symbol | Term | Description |
| --- | --- | --- |
| $\gamma_{1j}$ or $\gamma_{\text{LoF},j}$ | LoF Burden Effect | Effect of a loss-of-function variant in the $j$th gene on a trait. |
| $\gamma_{2j}$ or $\gamma_{\text{Dup},j}$ | Duplication Burden Effect | Effect of duplicating the $j$th gene on a trait. |
| $\bar{\gamma}_1$ | Average LoF Burden Effect | The mean of the LoF burden effects for all genes. |
| $\bar{\gamma}_2$ | Average Duplication Burden Effect | The mean of the duplication burden effects for all genes. |
| GDRC | Gene Dosage Response Curve | The continuous relationship between expression and average trait value. |
| aGDRC | Average Gene Dosage Response Curve | The curve for a specific trait resulting from taking the average of GDRCs across all genes. |
| $\phi$ | Monotonicity | A measure of the amount of monotone signal present for a trait. |
| $\zeta$ | Trait Buffering | A signed measure of how much GDRCs deviate from linearity in a particular trait direction. |
| $\zeta_m$ | Monotone Trait Buffering | The component of $\zeta$ that is explained by contributions from monotone GDRCs. |
| $\zeta_{nm}$ | Non-monotone Trait Buffering | The component of $\zeta$ that is explained by contributions from non-monotone GDRCs. |
| — | Genome-wide Burden Effect | A class of estimates previously reported in the literature. In each case, an individual is assigned a certain burden value based on their LoF variant or CNV burden, which is then tested for association with the trait of interest. |

### A.2 Genome-wide Burden

We retrieved genome-wide burden effect estimates from work by Auwerx et al., where burden was defined as the number of genes affected by deletions or duplications [1]. We subset to traits where both the genome-wide deletion and duplication burden effects had a p-value of less than 0.05. In all cases, the deletion and duplication burden effects are in the same direction.

**Figure A.1.**
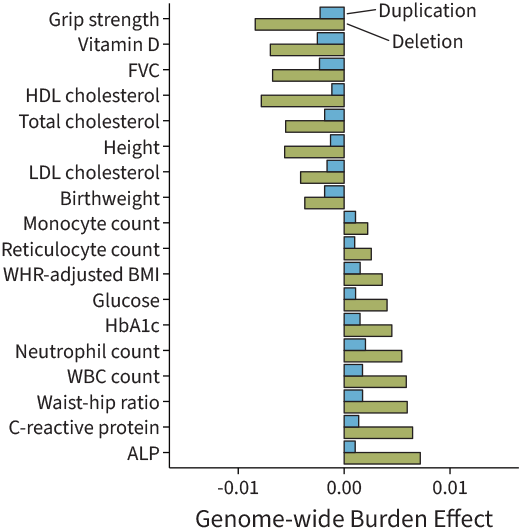
Genome-wide burden data, reprocessed from work by Auwerx et al. [1]. These effects have a p-value of less than 0.05. The deletion or duplication effect is the effect of increasing numbers of deleted/duplicated genes on the trait.

### A.3 Properties of Analyzed Variants in the UK Biobank

We analyzed burden genotypes based on four classes of variants. For each gene to be used in the analysis, we required the associated burden genotype to have a minor allele count (MAC) of five. For loss-of-function (LoF) variants, for example, this corresponds to the requirement that at least five of the 2*N* gene copies in the population contain a LoF allele.

LoF variants represent single nucleotide polymorphisms (SNPs) or indels that are predicted to produce transcripts that undergo nonsense-mediated decay (NMD). These variants are called using the whole-exome sequencing (WES) data and are annotated using computational variant-effect prediction tools. Sufficient LoF variants were present in 17,706 of the 19,432 genes with polymorphisms available in the WES data.

We also defined duplications, deletions, and potential loss-of-function (pLoF) variants using the copy number variants (CNVs) in the UK Biobank (UKB) whole-genome sequencing (WGS) data set. Duplications and deletions were required to encompass the entire gene along with a 1 Kbp flanking region. There were sufficient duplications for 38,566 genes, sufficient deletions for 18,854 genes, and sufficient pLoF variants for 13,810 genes. The larger number of genes available for the CNV data is due to the availability of calls across the genome from the WGS data, which we intersected with all the genes in the GENCODE release.

When intersected with the 17,706 genes from the LoF burden tests, there are 9,299 genes with sufficient duplications, 2,941 genes with sufficient deletions, and 7,645 genes with sufficient pLoF variants. These are upper bounds on the number of genes available to analyze for a trait, because we require at least five dosage-altering alleles to be present in phenotyped individuals.

### A.4 Scale of the Dosage and Trait

The scale and shape of a gene dosage response curve (GDRC) depends on how gene expression and the trait are measured. We assume that expression is measured on a logarithmic scale relative to the population mean. This allows us to define the expression effect of LoF and duplication variants on a logarithmic scale such that expression-decreasing variants have negative expression values and expression-increasing variants have positive expression values. A heterozygous LoF genotype thus has an expression effect of log 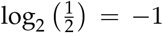, and a duplication resulting in a copy number of three has an expression effect of log_2_ 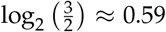. For traits, we chose to use an inverse rank normal transform. By centering the trait, we implicitly define trait-decreasing variants as having negative effects and trait-increasing variants as having positive effects. The consequence of centering both expression and trait is that the GDRC always passes through the origin.

### A.5 GDRCs for Protein Measurements

Our arguments generally require that LoF variants reduce expression, and duplications increase expression. To assess this, we ran LoF and duplication burden tests on the plasma protein measurements from the UK Biobank [2].

We performed burden tests for the plasma expression of 2,923 proteins in unrelated individuals using REGENIE, following the same procedure as our main burden analysis. We used the normalized protein expression (NPX) levels from the UKB [2]. We used the following covariates for both the first and second steps in REGENIE: sex, age, age-squared, age-by-sex, age-squared-by-sex, 20 genetic principal components (PCs), UKB center, whether the participant was preselected either by the UKB-PPP or as a part of the COVID-19 repeat-imaging study, genotyping batch, protein batch, WGS sequencing provider, WGS shipment, time from sampling to sample processing, and 40 protein PCs. The protein PCs were calculated from the NPX protein data using the irlba package in R.

We next used adaptive shrinkage [3] to model the burden effect estimates of variants on their cognate proteins. Briefly, adaptive shrinkage is an empirical Bayes approach that fits a flexible prior distribution for the latent effect sizes. Biological intuition suggests that LoF variants in a gene should reduce expression of the cognate protein, while duplications should increase expression of the cognate protein. We used the posterior mean estimates from this model for visualization. We found that in the vast majority of cases, LoF variants reduced and duplications increased the expression of the cognate protein in the blood proteome (Figure A.2).

**Figure A.2.**
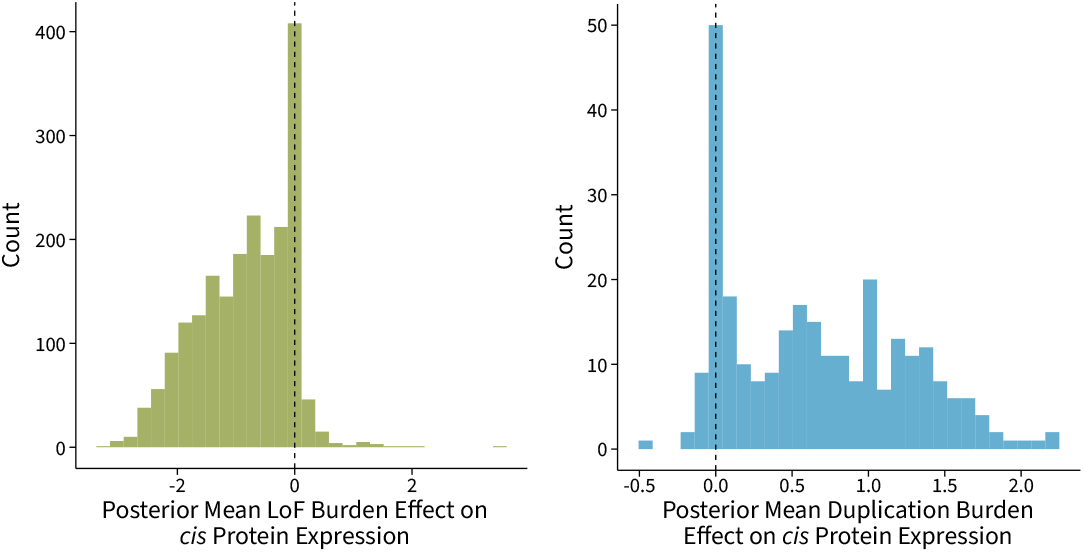
(Left) The distribution of the posterior mean LoF burden effect on the cognate protein’s expression in whole blood. (Right) The distribution of the posterior mean duplication burden effect on the cognate protein’s expression in whole blood.

We had both LoF and duplication burden effect estimates for a subset of the proteins. Comparing these estimates, we see that the relationship between gene expression and protein levels in plasma is predominantly monotone and increasing, as our biological intuition would suggest (Figure A.3).

**Figure A.3.**
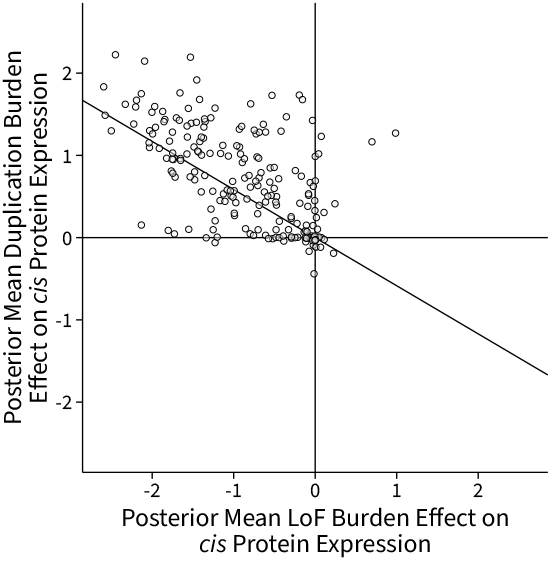
The duplication versus LoF burden effect estimates for proteins in the plasma. These are posterior mean effect sizes from the adaptive shrinkage model. These data suggest that the relationship between expression and protein levels in blood is monotone and increasing. Points on the diagonal line would correspond to linear GDRCs.

### A.6 Signal in Burden Tests

We used Storey’s method [4] to conservatively estimate the number of null tests in the LoF and duplication burden tests we performed across all traits. Briefly, Storey’s method assumes that the tests consist of a mixture of true positive and null tests, and attempts to estimate the proportion of true nulls 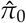. We estimated that the data are consistent with no less than 5% of gene-trait pairs being true associations (Figure A.4). This suggests that the burden test data are generally sparse or underpowered, making the identification of GDRCs for individual genes challenging at the genome-wide significance level.

**Figure A.4.**
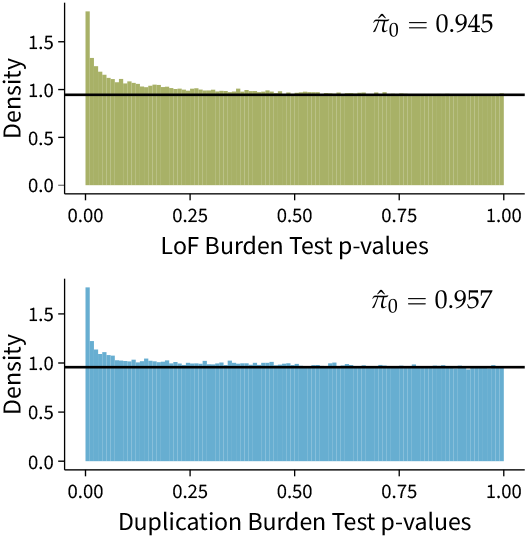
We conservatively estimated the number of true positive tests using Storey’s method [4]. The estimated proportion of null tests is reported as 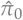. (Top) The distribution of p-values for the LoF burden tests across the traits selected for analysis in this paper. (Bottom) The distribution of p-values for the duplication burden tests across the traits selected for analysis in this paper. These p-values are derived from the maximum likelihood estimate for the burden test, which accounts for correlations between burden genotypes.

### A.7 Composition of Monotone Curves

Let *γ* : *E* ↦ *Y* be the GDRC for a gene. Suppose that the dosage effect percolates to the trait via a path, as diagrammed in Figure 4A. Consider a path consisting of two edges with associated functions *f* : ℝ →ℝ and *g* : ℝ → ℝ. The GDRC along this path will then be *Y* = *γ* (*E*) = *f* (*g* (*E*)) = (*f* ◦ *g*) (*E*). Here, *f* and *g* are the curves of individual arrows along the path.

Now, suppose that *g* is a monotone increasing function. That is, *x* ≤ *y* implies that *g* (*x*) ≤ *g* (*y*) for any *x, y ∈* ℝ. Similarly, suppose that *f* is a monotone increasing function. Then, *x* ≤ *y* implies that *g* (*x*) ≤ *g* (*y*), which implies that *f* (*g* (*x*)) ≤ *f* (*g* (*y*)). Thus, the composition (*f* ◦ *g*) is also monotone increasing. Similar arguments for monotone decreasing functions or pairs of increasing and decreasing functions demonstrate that the composition of monotone functions is also monotone.

We can develop more intuition for this property if we assume that the GDRC is a composition of differentiable functions,

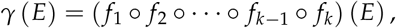

If the GDRC is monotone, then either *γ*^*′*^ (*E*) ≥ 0 or *γ*^*′*^ (*E*) ≤ 0 for all *E*. Using the chain rule, the derivative *γ*^*′*^ (*E*) is a product of component derivatives,

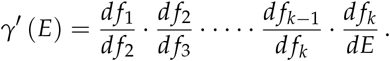

If each step in the path is monotone, then each component derivative is either nonnegative or non-positive over its entire domain. The product of such functions is either nonnegative everywhere or nonpositive everywhere. Therefore, if each step along the path is monotone, the resulting GDRC must be monotone.

## B Monotonicity Model

Here, we endeavor to jointly model burden summary statistics from different points on the dosage spectrum. Burden summary statistics are often reported as an effect size and a standard error from regression. We use hierarchical models to account for the sampling error and to pool information across genes.

Let **Y** *∈*ℝ^*N*^ be a standardized phenotype of interest, measured in *N* individuals. Let **X, Z** *∈*ℝ^*N×M*^ represent the burden genotype matrices for two variant classes respectively. By variant class, we mean a set of variants with a dosage effect in the same direction, such as deletions, duplications, or LoF variants. We assume that there are *M* genes that are polymorphic for both classes of variants. The genotypes are encoded as copies of the dosage-perturbing allele.

We use the following linear system to model the phenotype:

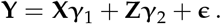

Here, ***γ***_1_, ***γ***_2_ *∈*ℝ^*M*^ are the unobserved, per-allele effect sizes of perturbations of each gene on the phenotype. The residual error is represented by ***ϵ*** *∈*ℝ^*N*^ and is assumed to be drawn from an isotropic normal distribution,

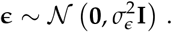

### B.1 Distribution of Effect Sizes

We model the latent effect sizes as coming from an uncentered normal distribution. Effect sizes between genes are assumed to be independent, but effect sizes for the same gene covary between the variant classes. That is to say, the effect of a deletion or LoF variant is assumed to carry some information about the effect of a duplication of the same gene. This covariance structure is represented with diagonal matrices **Σ**_11_, **Σ**_12_, and **Σ**_22_.

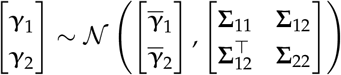

Suppose we collect the block matrices above into ***γ***, 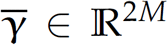 and **Σ** *∈*ℝ^2*M×*2*M*^. Then, we can succinctly state that

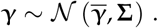

Let ***γ***_*·j*_ *∈*ℝ^2^ represent the latent effect sizes for the *j*th gene. For instance, the first coordinate may represent the LoF effect, and the second coordinate may represent the duplication effect. Then, the marginal distribution is written as

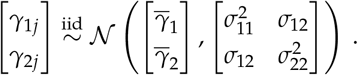

### B.2 Parameter of Interest

We consider a gene to have a “monotone” effect on a trait if the dosage-reducing alleles have an opposite direction of effect compared to dosage-increasing alleles. To this end, we define the monotonicity, *ϕ*, as a value proportional to the negative uncentered second moment of the effect sizes,

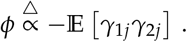

Thus, a positive *ϕ* represents monotone behavior. To compare monotonicity across traits, we restrict its codomain to [−1, 1]. We do this using the Cauchy-Schwarz inequality, which guarantees that

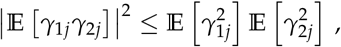

so that

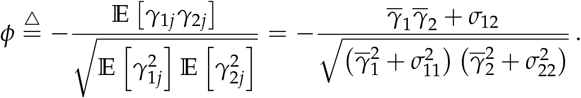

If the effect sizes have mean zero (that is,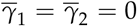), this is equivalent to the negative correlation coefficient,

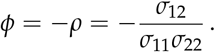

### B.3 Distribution of Genotypes

We consider burden genotypes in this analysis, which represent an aggregate of the genotypes across multiple sites. Specifically, we collapse different alleles that have the same effect on dosage into a single allele for each gene. Burden genotypes are encoded using the same scheme as biallelic variants.

We begin by assuming that there is no correlation between the different variant classes. This is reasonable to assume because different variants arise through different mutational processes, and burden genotypes represent aggregates of multiple rare variants, each of which generally have negligible linkage disequilibrium (LD) [5]. However, this assumption does not apply across genes within a variant class. Within duplications, for instance, large duplications consisting of multiple genes will induce correlations between the burden genotypes. This burden genotype correlation is similar to LD between biallelic genotypes at unique sites.

We model the different variant classes at a gene separately. We assume that the *M* burden genotypes are in Hardy-Weinberg equilibrium (HWE), which implies a binomial likelihood. Let *p*_*j*_ represent the allele frequency of the *j*th gene’s dosage-perturbing allele for the first variant class and let *q*_*j*_ represent the allele frequency of the *j*th gene’s dosage-perturbing allele for the second variant class. Without loss of generality, we assume that the genotypes are centered but not scaled:

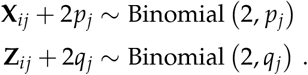

Our assumptions imply that there is no correlation between variant classes,

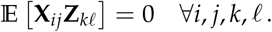

However, we do model the correlation between the same variant class. The correlation matrices **R**_1_ and **R**_2_ are used to represent this correlation. Let *P*_*j*_ = 2*p*_*j*_ (1 − *p*_*j*_) and *Q*_*j*_ = 2*q*_*j*_ (1 − *q*_*j*_) represent the heterozygosity of the burden genotypes for the *j*th gene. Then,

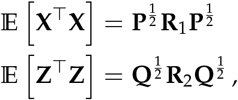

where

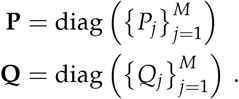

Because LoF variants affect individual genes, and we restrict ourselves to analyzing rare variants, we assume that the LoF burden genotypes are independent and that there is no correlation between them. That is, **R**_1_ = **I**.

### B.4 Burden Regression

Burden test results are reported as summary statistics from regression between the burden genotype and the phenotype. Regression is performed on centered and scaled phenotypes. The marginal summary statistics are calculated based on the following model for each gene:

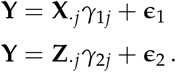

The effect size is approximately the following. Note that 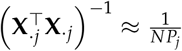 and 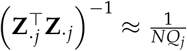,so

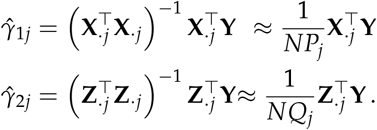

The standard error is approximately the following. Here, we assume that any individual gene explains a small fraction of the total variance. That is, 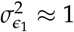 and 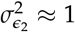. Thus,

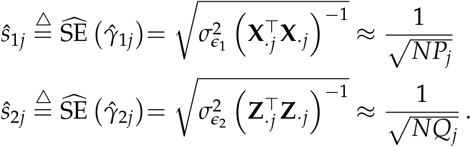

Therefore, the Z scores are

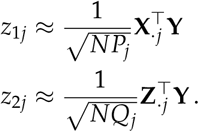

## C Monotonicity Inference

### C.1 Maximum Likelihood Estimation

For inference, we will use the summary statistics directly rather than the underlying genotype data. That is, we are given the estimated effect sizes (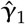 and 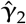) and their standard errors (**ŝ**_1_ and **ŝ**_2_). To simplify notation, we represent the prior as

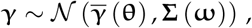

where **θ** and **ω** represent values used to parameterize the priors.

Next, the likelihood of the estimated effect sizes is defined using the approximate regression with summary statistics (RSS) likelihood [6]. Under this likelihood, the estimated effect sizes are patterned by the burden genotype correlation in the cohort. Furthermore, the standard error of the sampling distribution is used as an estimate of the dispersion around the mean. Zhu et al. showed that this likelihood asymptotically approaches the sampling distribution of the estimated effect sizes [6]. Let 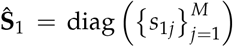 and 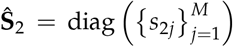, and let 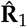 and 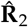 represent the in-sample correlation matrices of the two variant classes respectively. Then, in notation reflecting the RSS likelihood, we define

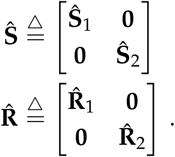

Then, the RSS likelihood for the observed effect sizes is

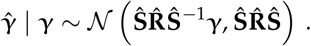

Some genes are perfectly correlated with each other. That is, the sample correlation matrix 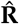 is not strictly positive definite. To improve numerical stability and to account for perfect correlation, we project the data onto the linear subspace of dimension *L < M* spanned by the correlation matrix (that is, a projection orthogonal to the null space). Consider the following eigendecomposition, with a matrix with orthogonal columns of eigenvectors **U** *∈* ℝ ^*M×L*^ and a diagonal matrix of positive eigenvalues **Λ** *∈*ℝ^*L×L*^,

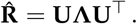

In practice, these are derived by dropping small eigenvalues from the numerical eigendecomposition of 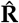. Rather than modeling the observations 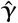, we model the projected data,

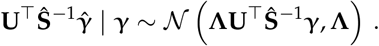

Note that under this model, the marginal likelihood of the estimates is

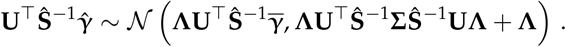

To simplify notation, let **A** = **ΛUŜ** ^−1^ and let **K** = **AΣA**+ **Λ**. Then,

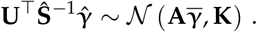

The pseudoinverse **A**^+^ is a useful quantity in downstream derivations. We can construct a pseudoinverse using components of the eigendecomposition. The pseudoinverse is

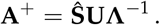

This pseudoinverse is a right pseudoinverse, as can be seen by

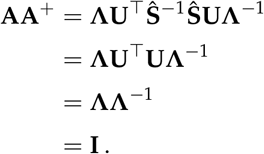

The other properties of the pseudoinverse are readily confirmed using matrix algebra.

#### C.1.1 Inference

We maximize the marginal likelihood of the observed effect sizes with respect to the parameters of the model. The likelihood is

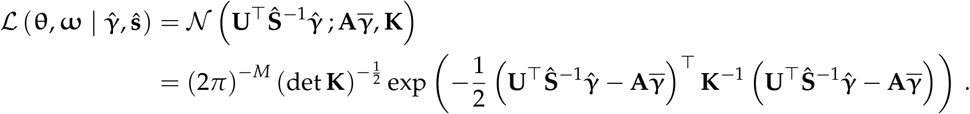

The log likelihood is

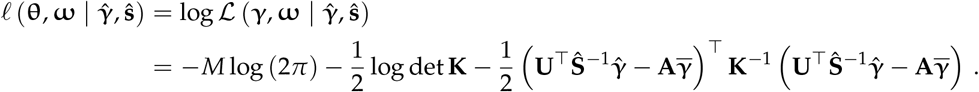

We are interested in obtaining the maximum likelihood estimates 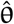 and 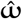 such that

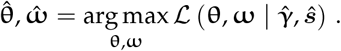

For maximum likelihood estimation of ***θ*** and ***γ***, we use natural gradient ascent [7]. This is an optimization approach that uses both first- and second-order information about the log likelihood via the gradient and Fisher information matrix. The gradient of the log likelihood function with respect to 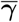 is

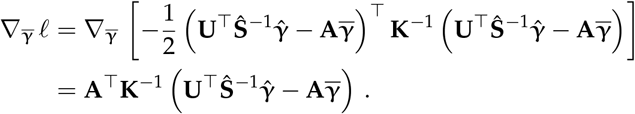

Using the matrix chain rule, the derivative of the log likelihood with respect to one of the mean parameters is

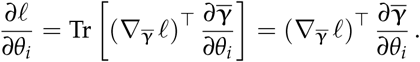

Therefore, it follows that the gradient of the log likelihood with respect to the mean parameters is

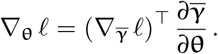

The matrix derivative of the log likelihood with respect to **K** is

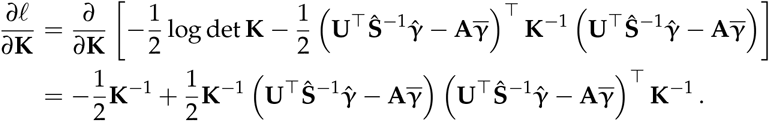

The derivative of **K** with respect to a given covariance parameter is

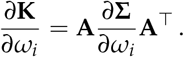

Using the matrix chain rule, the derivative with respect to a given covariance parameter is

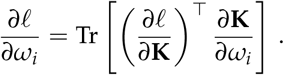

The Fisher information for multivariate normal distributions has a special form [8] such that the mean and covariance parameters do not share any information,

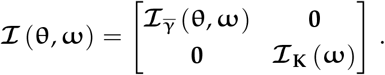

The Fisher information of the mean parameter is

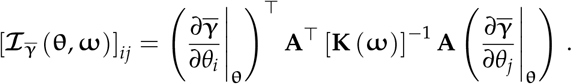

The Fisher information of the covariance parameters is

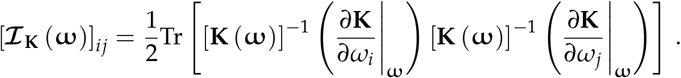

Natural gradient ascent involves Newton-Raphson updates with the Fisher information matrix. A dampening parameter 0 *< α*_*t*_ ≤ 1 is chosen using a backtracking line search [9] at each iteration to improve stability and serve as a stopping condition,

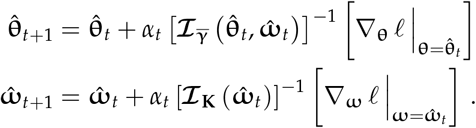

In practice, we estimate the gradients and Fisher information matrix for each chromosome separately and sum them up because effect estimates are assumed to be independent across chromosomes. The derivatives 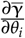 and 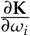 are computed using automatic differentiation [10, 11].

#### C.1.2 Uncertainty Estimation

We use the delta method to estimate the uncertainty in 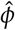. Since *ϕ ∈* [−1, 1], the delta method struggles to estimate the uncertainty near the boundaries. Instead, we estimate the standard error for

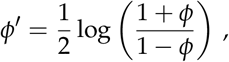

which is the Z transformation used to estimate confidence intervals for correlation coefficients. The inverse map is

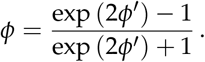

Let 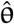 and 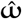 be the maximum likelihood estimates (MLEs) of ***θ*** and ***ω*** respectively. By the convergence properties of the MLE,

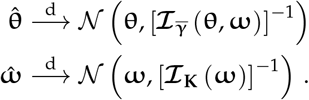

By the delta method, the estimator for *ϕ*^*′*^ converges to

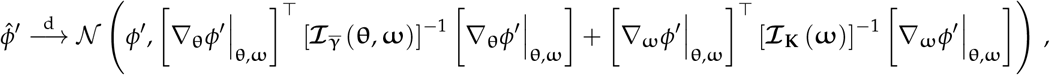

which can be used to derive approximate confidence intervals. The confidence intervals for 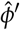 were inferred using this method, and the inverse map was used to determine the boundaries for the confidence intervals for 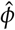.

### C.2 Fixed-Effects Model

In addition to consistent estimators provided by maximum likelihood estimation, it is useful to have method-of-moments (MoM) estimators of the parameters in our model, which avoid some of the assumptions on the distribution of ***γ*** that we make in the maximum likelihood approach. In some cases, these estimators have the additional advantage of being unbiased. In the fixed-effects model, we assume that ***γ*** is fixed, implying that the observations have the following likelihood:

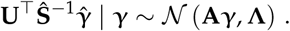

For unbiased estimation, the projection matrix **A**^+^ **A** is used frequently throughout our derivations. To see that it is a projection matrix, note that

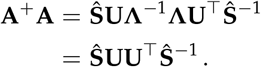

Then, it follows that

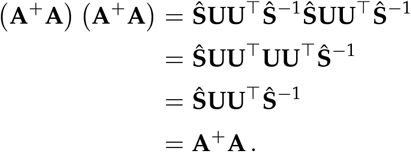

This makes **A**^+^ **A** an idempotent matrix and implies that it is a projection matrix.

#### C.2.1 Mean Effect

Suppose that the mean effect size is 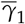 for the first variant class and 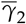 for the second variant class. Then,

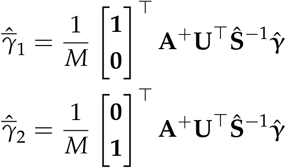

are unbiased estimators for these mean effect sizes under certain conditions discussed below. Since the estimator is the sum of normal random variables, we can easily construct confidence intervals from the variance. A straightforward computation shows that the variances of the estimators are

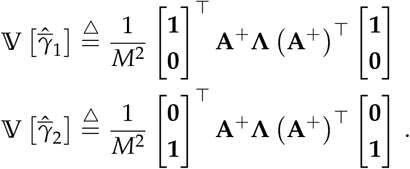

These estimators are guaranteed to be unbiased if **Ŝ** ^−2^ [ **1 0** ] ^⊤^and **Ŝ** ^−2^ [ **0 1** ] ^⊤^are orthogonal to the null space of **U**^⊤^.

To see that 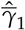 and 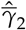 are unbiased under these conditions, we will focus on the former without loss of generality. In expectation,

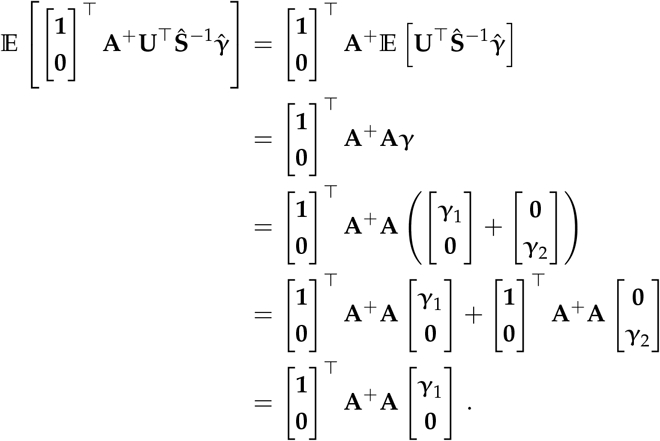

The last equality holds because **A**^+^ **A** is block diagonal. Note here that **A**^+^ **A** is a projection matrix and has the form **A**^+^ **A** = **ŜUU**^T^**Ŝ** ^−1^. Therefore, for our estimator to be unbiased, we require that

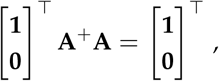

which is true if **Ŝ** ^−1^ [ **1 0** ] ^⊤^ is orthogonal to the null space of **U**^⊤^. A similar derivation demonstrates that for 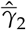 to be unbiased, **Ŝ** ^−1^ [ **0 1** ] ^⊤^must be orthogonal to the null space of **U**^⊤^. We tested this numerically with the eigendecomposition involving our specific burden genotype correlation matrix.

#### C.2.2 Mean Squared Effect

The mean squared effect is a useful measure of the amount of burden signal present for each trait. The following estimators,

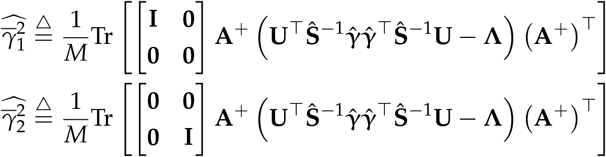

are unbiased estimators for the squared effect size if **Ŝ** ^−1^ [ ***γ***_1_ **0** ]and **Ŝ** ^−1^ [ **0*γ***_2_]^T^ are orthogonal to the null space of **U**^T^ respectively. Because the latent effect sizes are unobserved, we assume this within our model. If the assumption does not hold, our estimates will be biased and act as underestimates of the true squared effect size.

The variance of these estimators is challenging to compute. Instead, we invoke the Central Limit Theorem, noting that each element in the sum is an independent and unbiased estimate of the quantity of interest. We use an empirical estimate of the standard error to derive approximate confidence intervals.

Showing that 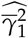 and 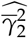 are unbiased under our assumptions follows a similar approach to 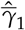 and 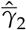. We will demonstrate that the first estimator is unbiased without loss of generality. To derive the expectation of the estimator, it is useful to note that

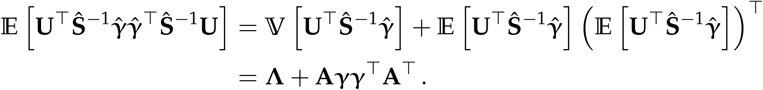

Using this derivation,

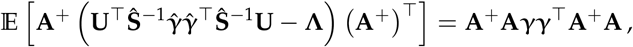

which follows because **A**^+^ **A** is symmetric. Since **A**^+^ **A** is also block diagonal,

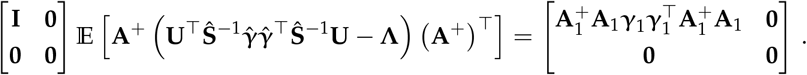

Note that 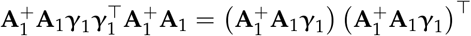 and therefore

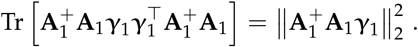

Taken together,

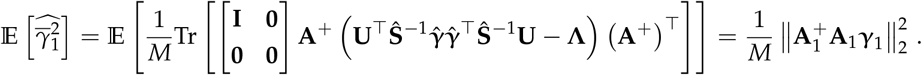

This estimator is thus unbiased when

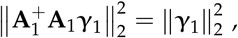

which is to say that the projection matrix 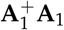 maps ***γ***_1_ to itself. This is the same as saying that **Ŝ** ^−1^ [ ***γ***_1_ **0** ] ^⊤^must be orthogonal to the null space of **U**^⊤^. Unlike the estimate of the average burden effect, we cannot test this empirically because the latent effect sizes are unobserved. Since 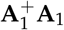 is a projection matrix, we have that

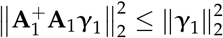

for any arbitrary ***γ***_1_. Therefore, if our assumption about the latent effect sizes is incorrect, we will underestimate the true squared effect size.

In practice, we model 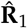 as the identity matrix for LoF burden tests, which makes 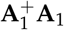 full rank. As such, we expect no bias in the estimate of 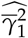. However, 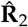 is empirically rank-deficient for duplications, which could potentially make 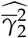 an underestimate of the true squared effect of duplications.

### C.3 Random-Effects Model

A random-effects model is required to develop estimators for the covariance components. We use an approach inspired by linkage disequilibrium score regression (LDSC) [12], and similar to that outlined in [6]. We assume that ***γ*** is random, with finite first and second moments. We also assume that the latent effect sizes are independent given **Ŝ** and 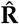. We make the following minimal assumptions about first and second moments of the joint distribution of the latent effect sizes:

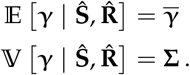

The Z scores from the burden regression can be defined as

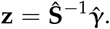

By the properties of the normal distribution, the Z scores are distributed as

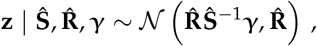

and Z scores for the two variant classes are independent,

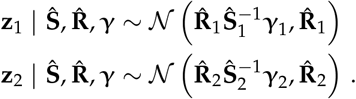

Without loss of generality for the second variant class, we focus on the *j*th gene of the first variant class. Let 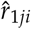 be the *ji*th element of 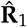. The marginal distribution of the Z score for a given gene is then

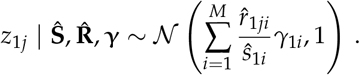

The marginal distribution of the Z scores between the two variant classes is

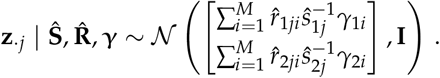

We define the scaled correlation score for the *j*th gene as

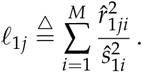

This is akin to the LD score used in LDSC, but is scaled by the variance of the gene burden test estimate. It is also useful to define

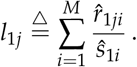

We define the cross-variant correlation score as

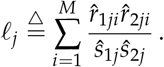

#### C.3.1 Mean Effect

Under the random-effects model, there is additional variance that needs to be accounted for in the estimator for the mean effect. The fixed-effects estimators from Appendix C.2.1,

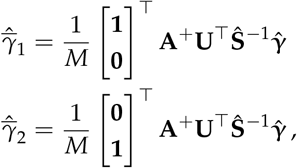

are unbiased estimators for 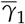 and 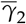 under the random-effects model as well. As before, we require that **Ŝ** ^−2^ [ **1 0** ] ^⊤^and **Ŝ** ^−2^ [ **0 1** ] ^⊤^are orthogonal to the null space of **U**^⊤^, which we have tested empirically for our traits.

To show that these estimators are unbiased, we will focus on 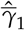 without loss of generality. In expectation,

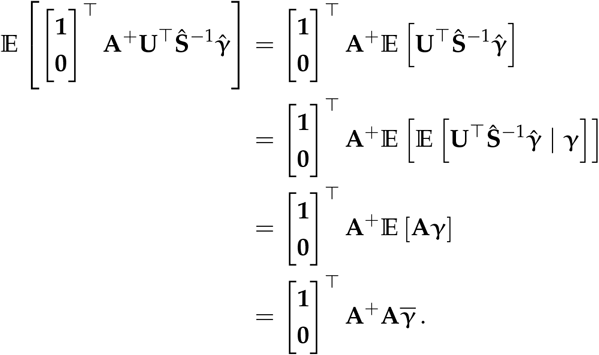

From here, the rest of the derivation is the same as the fixed-effects model estimators in Appendix C.2.1.

The variance of this estimator is greater than the variance of the estimator from the fixed-effects model. First, we derive the variance of the observed effect sizes,

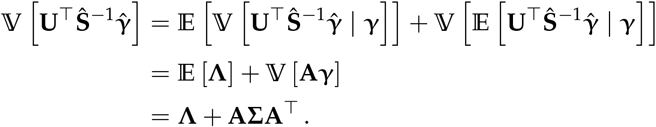

Using this result, the variance of 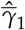 without loss of generality is

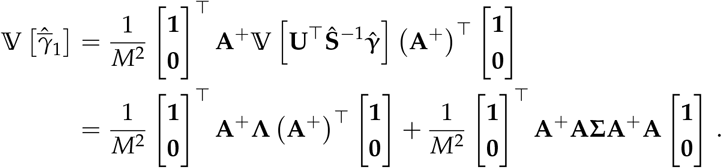

Recall that we assumed that [ **1 0** ] ^⊤^is mapped to itself under the projection matrix **A**^+^**A**. Furthermore, **Σ** is a block matrix, and the block **Σ**_11_ is a diagonal matrix where each diagonal entry is 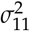 (Appendix B.1). Therefore,

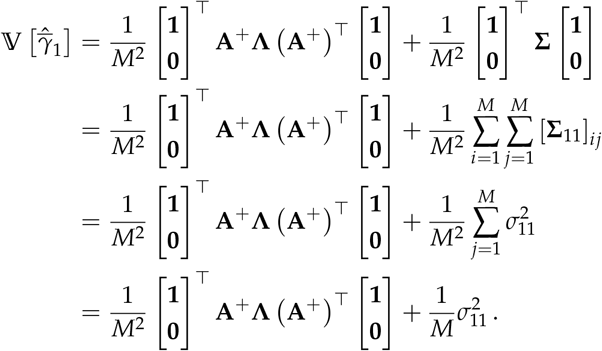

A similar derivation for 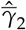 shows that

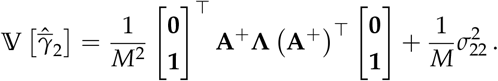

#### C.3.2 Variance of Latent Effects

Similar to LDSC, we now proceed by evaluating the expected squared Z score to develop estimators for 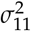 and 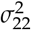. We integrate over the latent effect size, ***γ***, using the law of total expectation and the law of total variance. In the following, the conditioning on 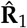, **Ŝ** _1_, *ℓ*_1*j*_, and *l*_1*j*_ is implicit to simplify notation. The expected squared Z score is

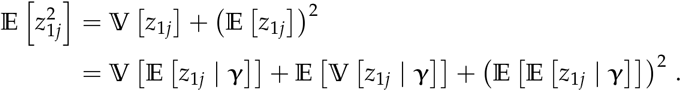

The first term introduces the correlation score into the expectation,

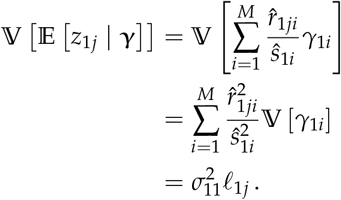

The second term introduces a constant,

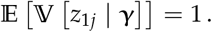

The last term is a bias introduced by the uncentered prior,

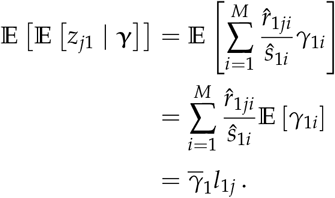

Taken together, the expected squared Z score is

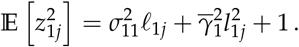

We can use the square of the estimator for the mean effect 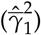 to approximate 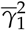, and account for the bias introduced by squaring the estimator. Specifically, recall that

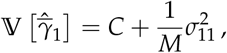

where *C* is a constant value (Appendix C.3.1). Then,

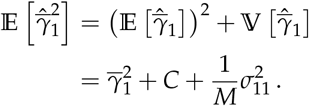

Given the relationship of the expected squared Z score with 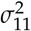, we propose the following unbiased estimator. A similar derivation follows for 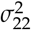. Our estimators are

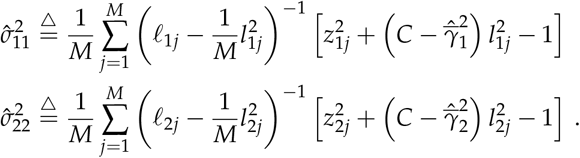

#### C.3.3 Covariance of Latent Effects

We can use the expected product of Z scores to build an estimator for the covariance *σ*_12_. We integrate over the latent effect size ***γ*** using the law of total expectation and the law of total covariance. In the following, the conditioning on 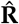, **Ŝ**, *ℓ*_*j*_, *l*_1*j*_, and *l*_2*j*_ is implicit to simplify notation. The expected product of Z scores is

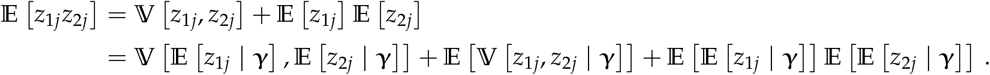

The first term introduces the cross-variant correlation score into the expectation,

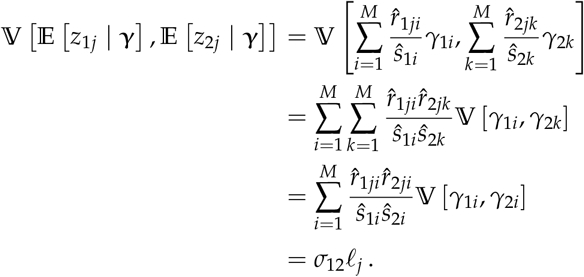

The second term does not contribute to the conditional expectation because the covariance between observed effect sizes is zero given the latent effect size:

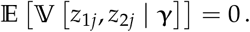

The last term is bias introduced by the uncentered prior,

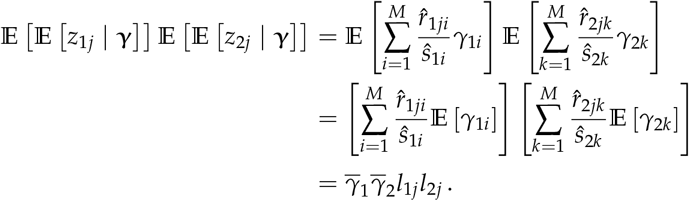

Taken together, the expected product of Z scores is

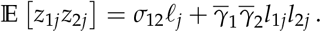

This suggests the use of the following unbiased estimator for *σ*_12_,

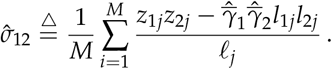

#### C.3.4 Monotonicity

These estimators of individual parameters in the model allow us to define the MoM estimator for monotonicity as

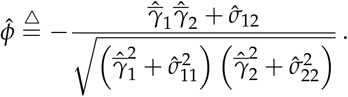

MoM estimators are not unbiased, but they are consistent. Since the estimator represents a function of various estimators, we used the bootstrap with 1000 iterations to estimate the standard error of the estimate. If the estimate was undefined due to a negative square root, we set 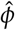 to zero.

## D Trait Buffering Model

In the trait buffering model, we are interested in evaluating if GDRCs are systematically buffered against one trait direction. This could occur if non-monotone GDRCs preferentially point in one direction over the other. This can also occur if monotone GDRCs achieve larger values for the trait in one direction compared to the other.

### D.1 Distribution of Effect Sizes

We use the multivariate adaptive shrinkage (MASH) model [13] as a flexible prior for our latent effect sizes. The MASH prior consists of a mixture of multivariate normal distributions. Suppose 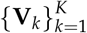 represents a fixed set of fixed covariance matrices in ℝ ^2*×*2^ for *K* mixture components. We use bivariate covariance matrices over a grid of variances and correlation values. Let ***π*** *∈* Δ_*K*−1_ represent the mixture weights. Then, the gene-level prior is

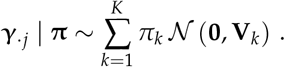

The prior over the entire set of *M* genes is

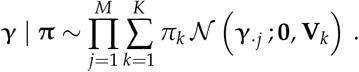

The likelihood model remains the same as before,

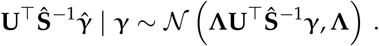

The joint likelihood is

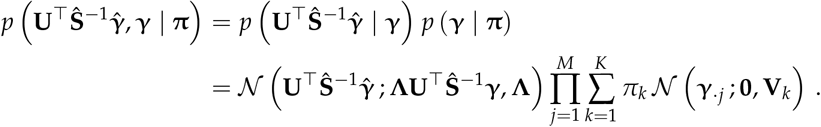

### D.2 Parameter of Interest

We define the estimand of interest as

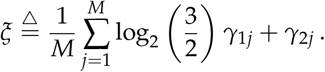

The estimand represents the dot product of ***γ***_*·j*_ with the normal vector of the diagonal separating the two types of buffering within our model (Figure D.1). The diagonal line satisfies

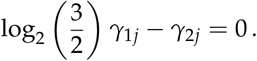

Thus, GDRCs on this diagonal line are linear GDRCs (Figure D.1).

**Figure D.1.**
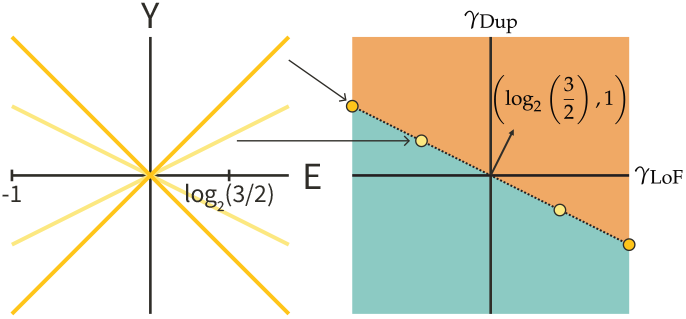
The boundary between buffering curves is represented by the yellow gene dosage response curves on the left. These map to a diagonal across the burden effect plot. To estimate signal in one direction of the diagonal versus the other, we use the normal vector in black shown on the right.

Each gene in the summation for *ξ* is either monotone or non-monotone. Genes are monotone if *γ*_1*j*_*γ*_2*j*_ *<* 0 (because the signs are opposite), while genes are non-monotone if *γ*_1*j*_*γ*_2*j*_ *>* 0 (because the signs are the same). We can rewrite the sum as

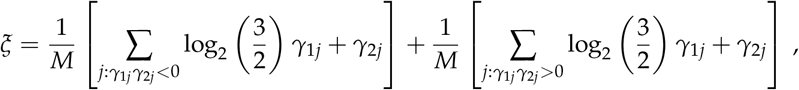

where the first term represents all the monotone genes, and the second term represents all the non-monotone genes. We define these two terms as

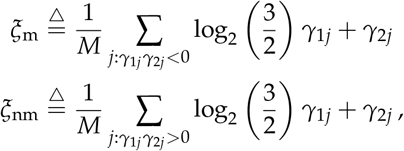

so that

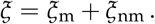

## E Trait Buffering Inference

In our model, the mixture weights, ***π***, are unknown. Following the method in MASH [3, 13], we use an empirical Bayes approach that combines both frequentist and Bayesian techniques. The first step, the frequentist step of empirical Bayes, involves maximizing the marginal likelihood to estimate 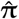, which we do using stochastic approximation expectation maximization (SAEM) [14, 15]. The second step, the Bayesian step of empirical Bayes, uses 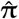 as a plug-in estimate of ***π*** for downstream posterior sampling.

The ideal approach to obtain the MLE of ***π*** is to directly maximize the marginal likelihood

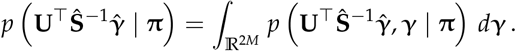

However, this integral is intractable. Instead, we can use expectation maximization (EM) to maximize this marginal likelihood [16]. Suppose ***π***^(*t*)^ represents the estimate at the *t*th iteration of the EM algorithm. The expectation step involves evaluating

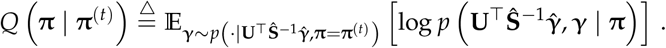

Since the analytical form of *Q*(***π*** | ***π***^(*t*)^) is also intractable under our model, we use SAEM to approximate the expectation. We sample 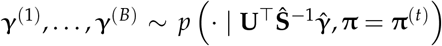 using Hamiltonian Monte Carlo [17, 18]. Then, we approximate the expectation with

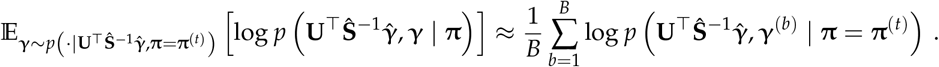

To speed up computation, we use stochastic gradient ascent, with each chromosome representing one mini-batch [19]. In the mini-batch approach, each expectation is a convex sum of the previous expectation and the current expectation. Let (*c*_*t*_)_*t*≥1_ be a positive, decreasing sequence such that 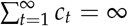and 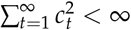. We use

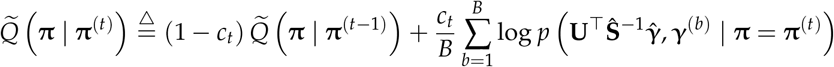

in the expectation step of EM. To define a valid recursion, we set 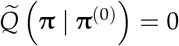. Maximization, the second step in EM, is performed using a fixed number of iterations of stochastic gradient ascent with a fixed step size to obtain

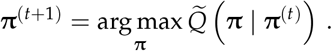

Gradients were estimated using automatic differentiation [10, 11]. For each batch, we used 50 iterations of gradient descent, using AdamW to optimize 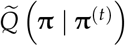 with an exponential decay learning rate schedule [20]. We run SAEM for at most five epochs, and estimate *Q* (***π*** | ***π***^(*t*)^) across all chromosomes at the end of each epoch. We stop early if the epoch estimate of *Q* (***π*** | ***π***^(*t*)^) decreases, and set 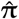 to the parameter value that maximized the epoch estimate of *Q* (***π*** | ***π***^(*t*)^).

We use Hamiltonian Monte Carlo [17, 18] to sample from the posterior distribution of ***γ***. Following the empirical Bayes approach, 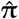 is used as a plug-in estimate. That is, we draw samples 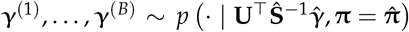. We use the posterior samples to estimate the cumulative density function of the posterior distribution of *ξ*. Specifically, if

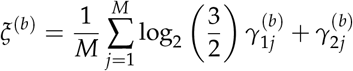

represents the value of *ξ* for the *b*th sample, then the empirical cumulative density function for the posterior distribution is

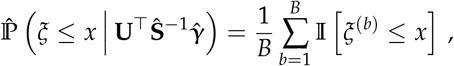

where 𝕀 [*·*] is the indicator function. Under this definition, the posterior mean is estimated as

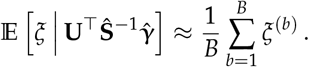

Furthermore, we estimated the 95% credible interval by using the 2.5th and the 97.5th percentile of the posterior density. The same procedure is used for *ξ*_m_ and *ξ*_nm_.

## F Interpretation of Monotonicity and Trait Buffering

We use *ϕ* as a measure of overall monotone signal for a trait. In Figure F.1, we provide some examples to aid in the interpretation of *ϕ*. If LoF and duplication variants have opposite effects on a trait for all genes, *ϕ* will be positive (Figure F.1A). In contrast, if LoF variants and duplications in all genes have the same direction of effect on a trait, *ϕ* will be negative (Figure F.1B). Thus, the sign of *ϕ* generally allows us to interpret the average behavior of the direction of effects of burden tests for a trait. The value of *ϕ* does not need to be 1 for all genes to be monotone, and it does not need to be -1 for all genes to be non-monotone.

**Figure F.1.**
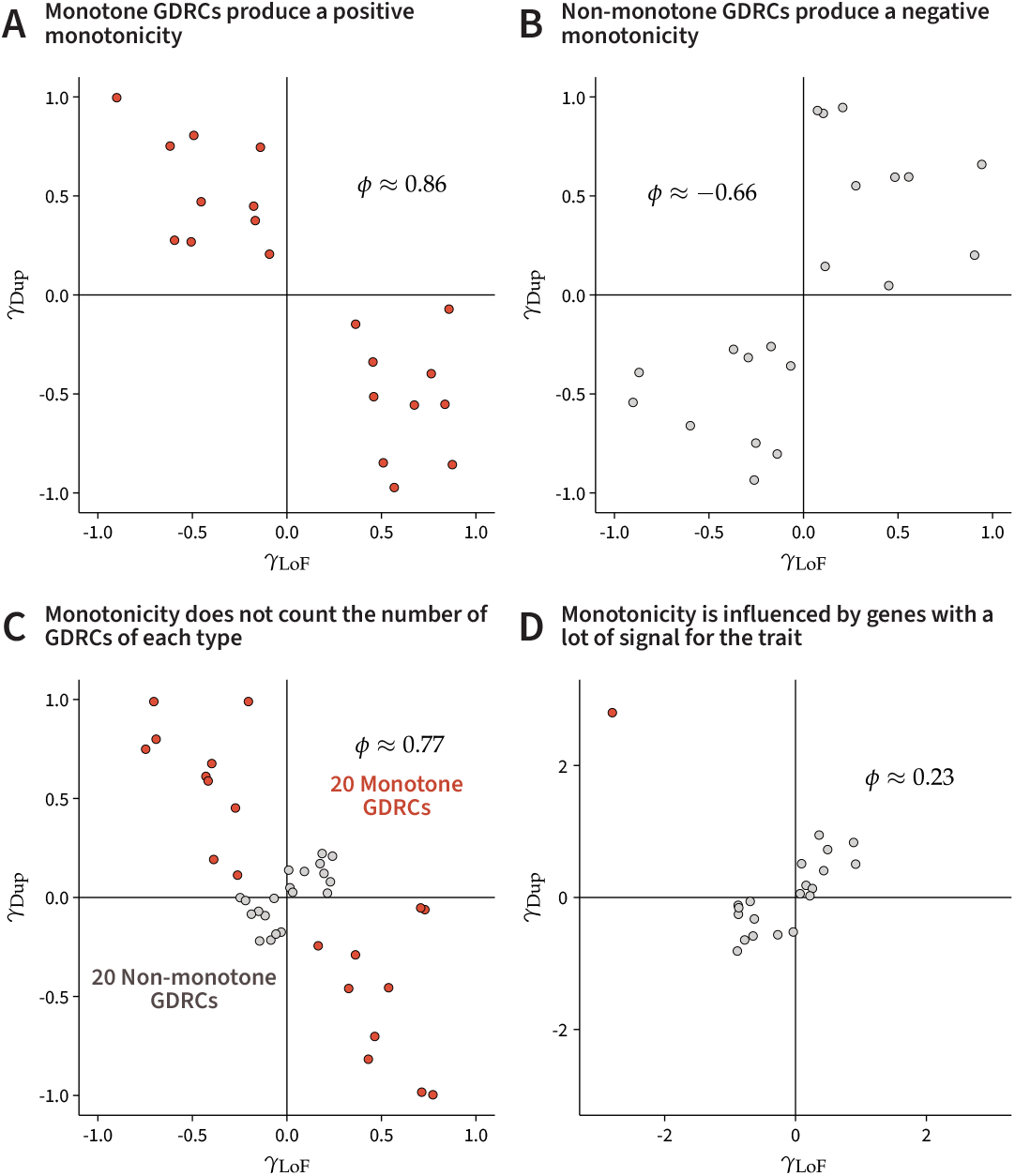
**A**. If loss-of-function variants and duplications have opposite effects on a trait for all genes, *ϕ* has some positive value. **B**. If loss-of-function variants and duplications have effects in the same direction on a trait for all genes, *ϕ* has some negative value. **C**. *ϕ* does not measure the number of monotone or non-monotone GDRCs. In this example, all quadrants have the same number of genes, but *ϕ* is positive because the monotone genes have larger effects on average. **D**. *ϕ* can be dominated by genes with large effects. In this example, all but one gene are non-monotone, but *ϕ* is still positive.

*ϕ* does not count the number of GDRCs present for a trait. As an example, consider Figure F.1C, where each quadrant contains the same number of genes, summing to 20 monotone GDRCs and 20 non-monotone GDRCs for the hypothetical trait. On average, the monotone GDRCs have larger effects on the trait, and thus *ϕ* is positive. This means that even one gene with a large effect can influence *ϕ* (Figure F.1D).

We use *ξ* to better understanding trait buffering. The goal is to use *ξ* to explain why the average GDRC is non-montone. In Figure F.2A, we display the burden effect sizes of the genes for a hypothetical trait experiencing negative trait buffering. Negative trait buffering means that GDRCs are buffered against increasing the trait value. For this trait, a majority of the contribution to this signal comes from non-monotone GDRCs (*ξ*_nm_ *≈* −0.23 and *ξ*_m_ *≈* −0.12). In our main figure (Figure 5C), we report the posterior means for the components *ξ*_m_ and *ξ*_nm_, which add up to exactly equal our estimate for *ξ* based on the law of total expectation.

**Figure F.2.**
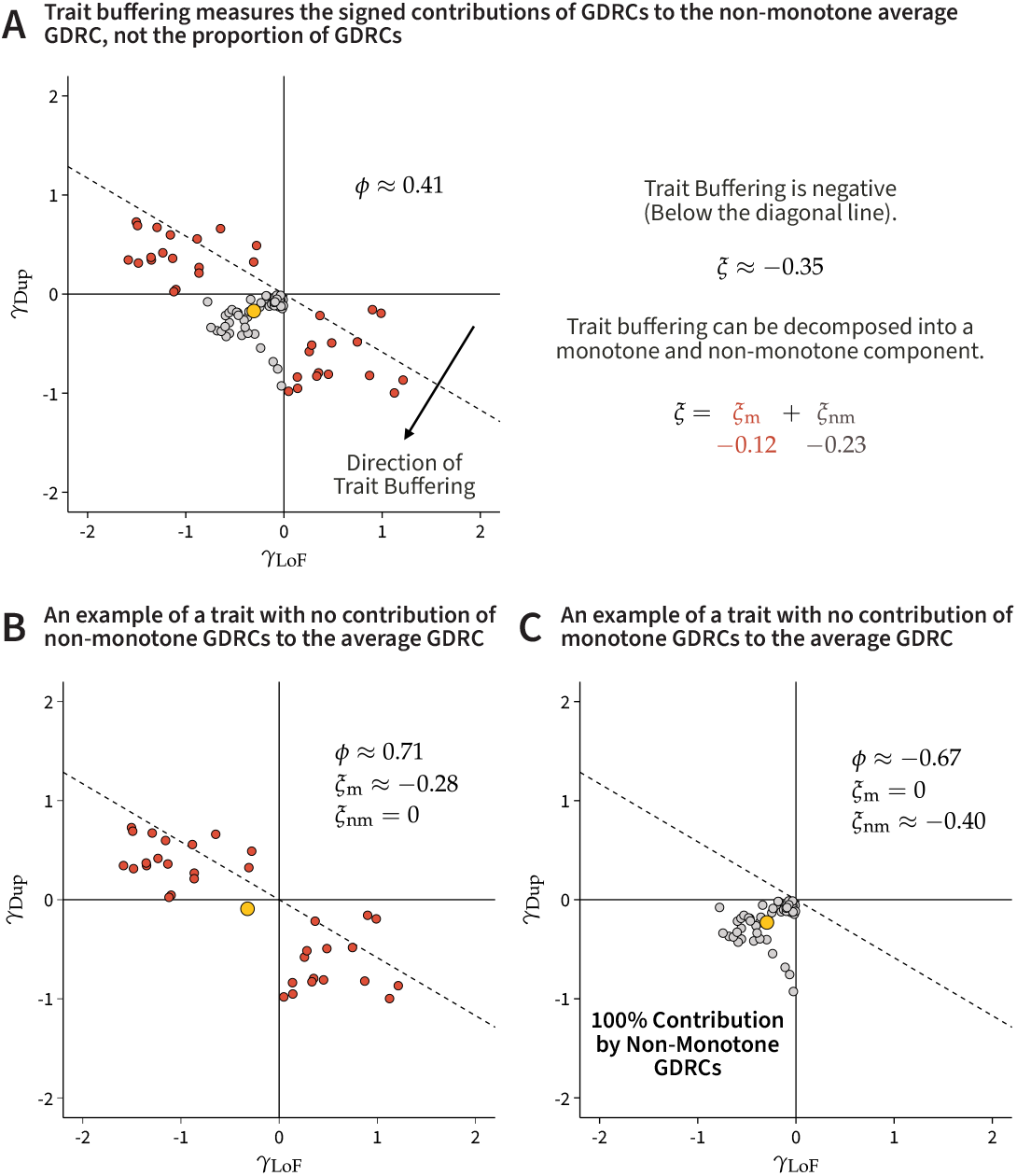
**A**. Trait buffering measures the signed deviation of GDRCs from the diagonal line, which explains the non-zero average GDRC (larger yellow point). **B**. Trait buffering can occur with only monotone GDRCs present (the average GDRC is still non-monotone). **C**. Trait buffering can occur with only non-monotone GDRCs present.

To understand the meaning of the contributions, we provide two examples where all of the contributions to trait buffering come from either monotone GDRCs (Figure F.2B) or non-monotone GDRCs (Figure F.2C).

## G Simulations

### G.1 Monotonicity Simulations

We used the UKB LoF and duplication burden genotypes for simulation. These simulations are not from the models we used to derive the various estimators. Thus, these tests are fair evaluations of our estimators and also demonstrate the validity of using the RSS likelihood to model burden summary statistics. Some of our estimators assume fixed effect sizes, while others assume random effect sizes. We performed two separate suites of simulations for these estimators.

The fixed-effect simulations assumed that each gene had some fixed effect 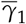 via LoF variants and 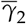 via duplication variants. We also performed fixed-effect simulations where each gene had a unique effect, with a genome-wide average effect of 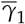 and 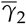 respectively. The random-effect simulations assumed that the effect size for each gene was generated from

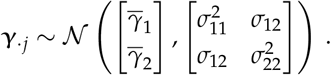

We simulated effect sizes for genes that had both LoF and duplication burden genotypes, corresponding to passing our genotyping filters described in our methods. Once effect sizes were assigned to each gene, we used PLINK [21] to score each sample based on the effect sizes. This provided us with genetic values for each individual under the simulation. Next, we added noise using draws from a normal distribution such that the expected variance of the phenotype would be one.

We used PLINK to run a single-variant association scan with the burden genotypes. We included 15 genotyping PCs, 20 rare variant PCs, and genotyping batch as covariates. Since we simulated from the actual genotypes, we had to include these covariates to control for confounding from population structure. Each parameter value was tested with 100 simulations. When perturbing one parameter, the other parameters were held constant at 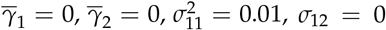, and 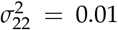. The default variance values are based on the order of magnitude of burden heritability estimates from prior work [22].

#### G.1.1 Method-of-Moments Average Burden Effect Estimators

Fixed-effect simulations were used to test 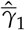 and 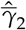. These estimators were convincingly unbiased for the various realistic values that we simulated (Figure G.1). We also developed estimators for the variance of these fixed-effect average burden effect estimators. Confidence intervals built using these variance estimates are well-calibrated (Figures G.2 and G.3). We also tested the estimators under a regime where each gene had a unique effect size, with relatively unbiased behavior and good calibration (Figures G.4, G.5, and G.6).

**Figure G.1.**
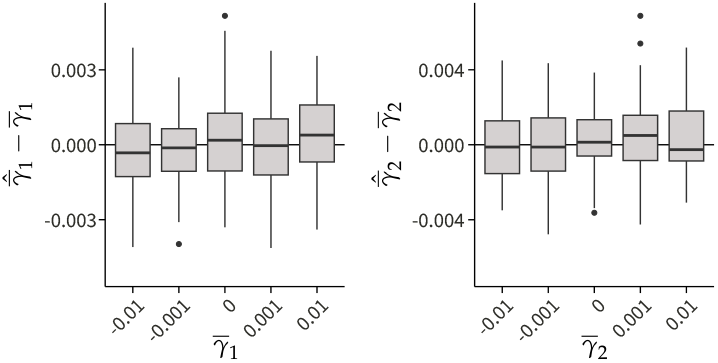
(Left) The difference in the estimated value 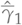 and the true value 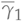 for various tested values are shown for 100 simulations. (Right) The difference in the estimated value 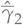 and the true value 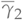 for various tested values are shown for 100 simulations.

**Figure G.2.**
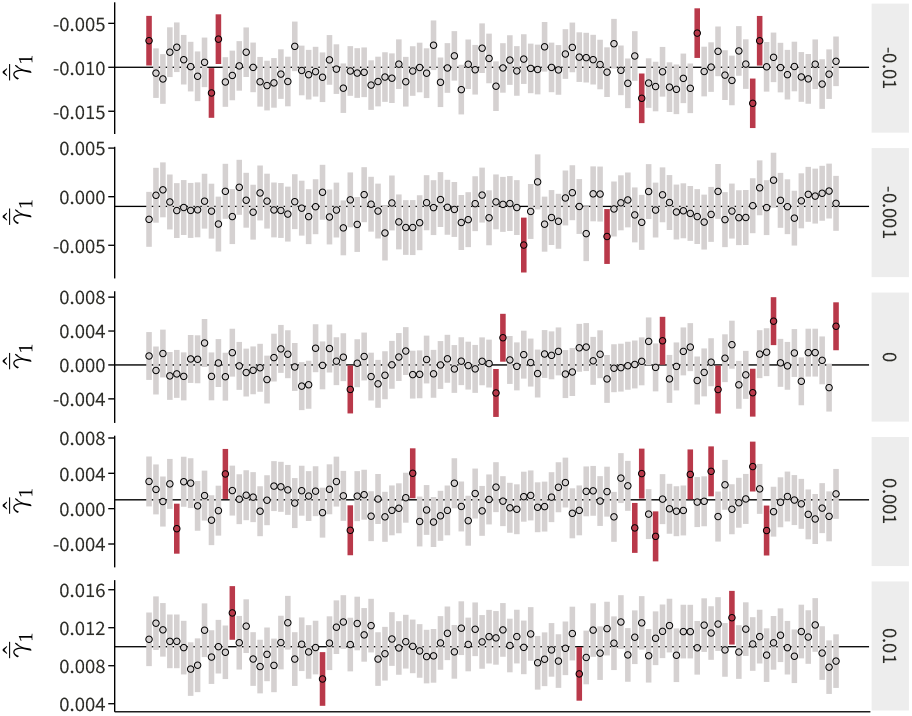
The plot shows 95% confidence intervals from 100 simulations for various values of 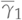. Confidence intervals in red represent realizations that do not cover the true value.

**Figure G.3.**
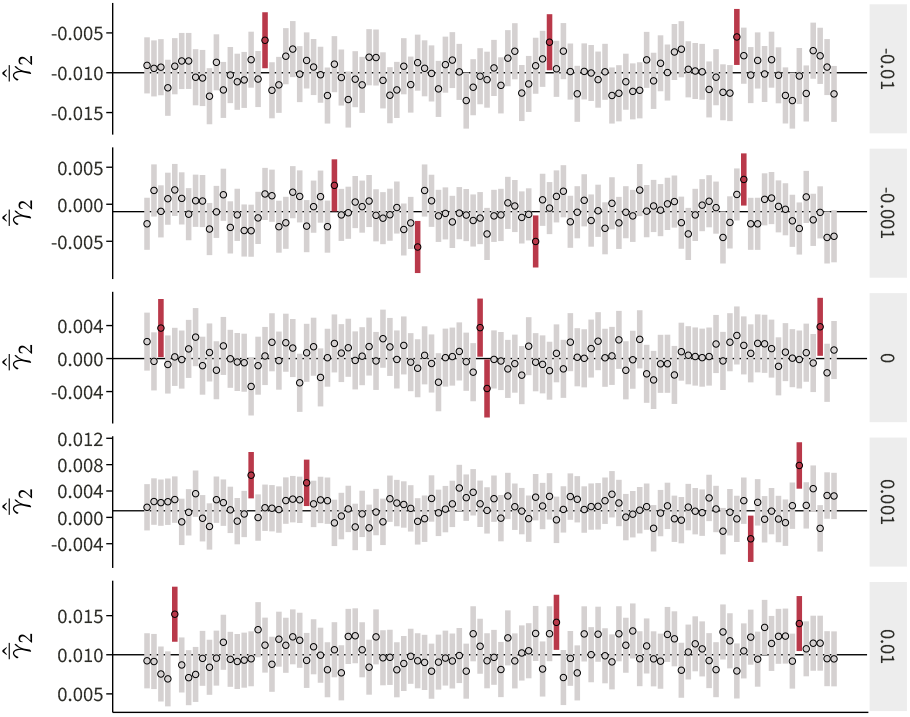
The plot shows 95% confidence intervals from 100 simulations for various values of 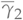. Confidence intervals in red represent realizations that do not cover the true value.

**Figure G.4.**
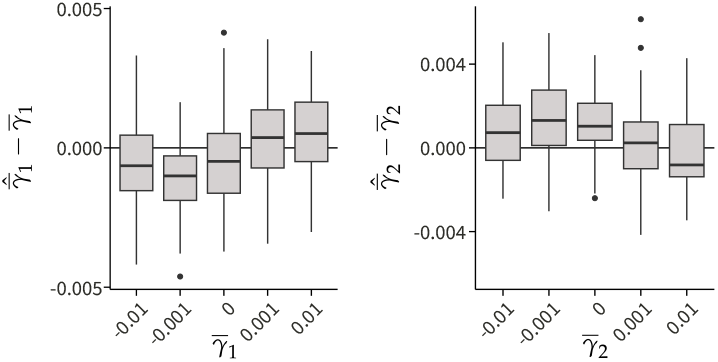
(Left) The difference in the estimated value 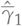 and the true value 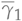 for various tested values are shown for 100 simulations. (Right) The difference in the estimated value 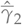 and the true value 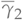 for various tested values are shown for 100 simulations.

**Figure G.5.**
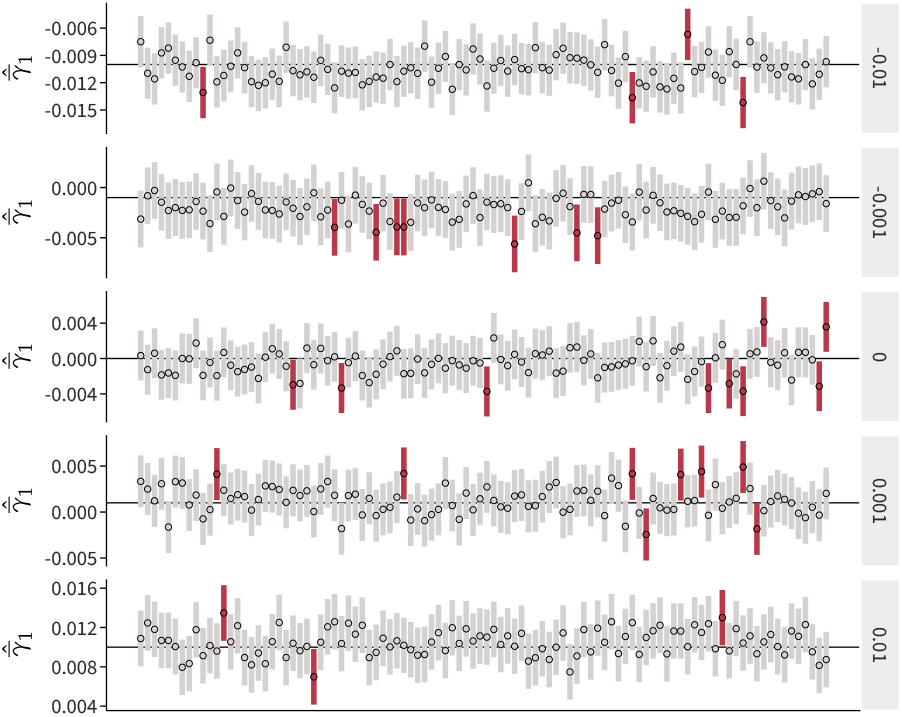
The plot shows 95% confidence intervals from 100 simulations for various values of 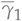. Confidence intervals in red represent realizations that do not cover the true value.

**Figure G.6.**
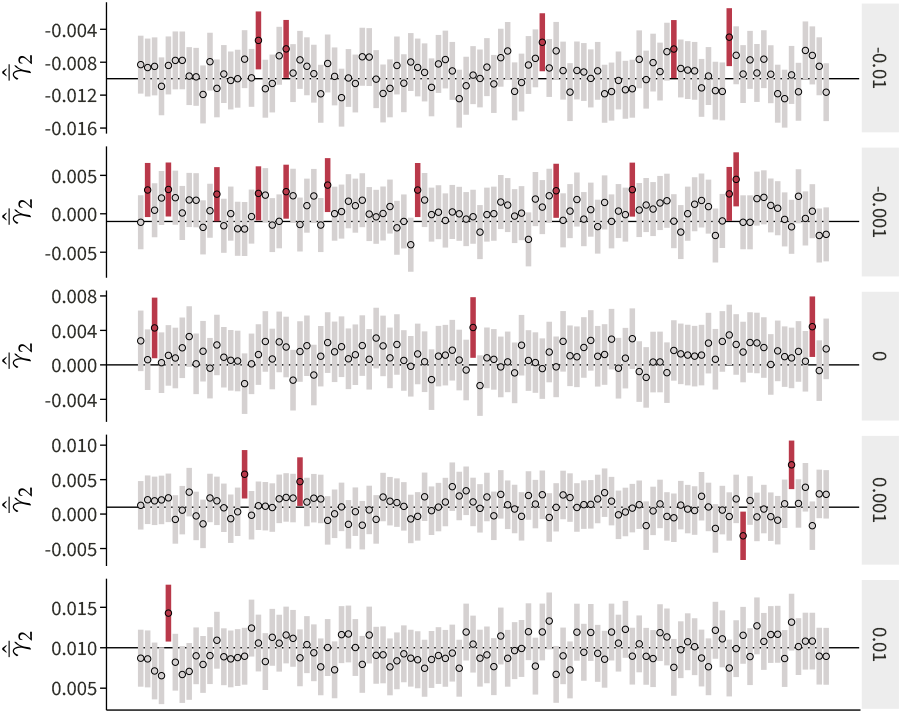
The plot shows 95% confidence intervals from 100 simulations for various values of 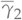. Confidence intervals in red represent realizations that do not cover the true value.

#### G.1.2 Method-of-Moments Average Squared Burden Effect Estimator

We used the fixed-effect simulations with different gene-level effects to test 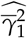 and 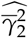. The estimator for 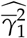 was relatively unbiased for various realistic values that we simulated (Figure G.7). However, the estimator for 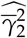 was biased downwards significantly (Figure G.7). As we discuss in our derivation of 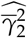, this is the effect of our assumption about the unobserved ***γ***, which is predicted to make our estimator conservative for the true value of the squared effect. The approximate confidence intervals using the Central Limit Theorem are well-calibrated for 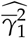 (Figure G.8) but uniformly underestimate 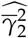 (Figure G.9).

**Figure G.7.**
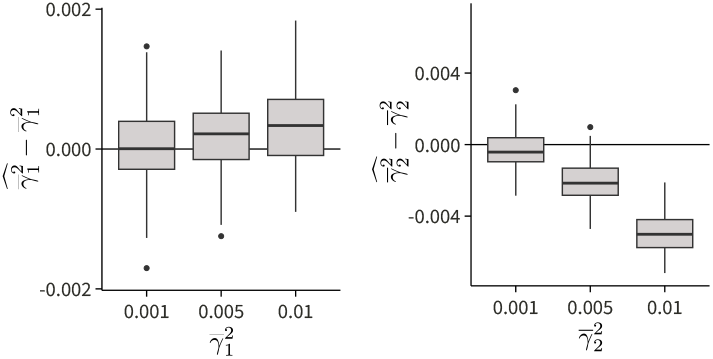
(Left) The difference in the estimated value 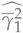 and the true value 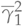 for various tested values are shown for 100 simulations. (Right) The difference in the estimated value 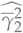 and the true value 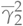 for various tested values are shown for 100 simulations.

**Figure G.8.**
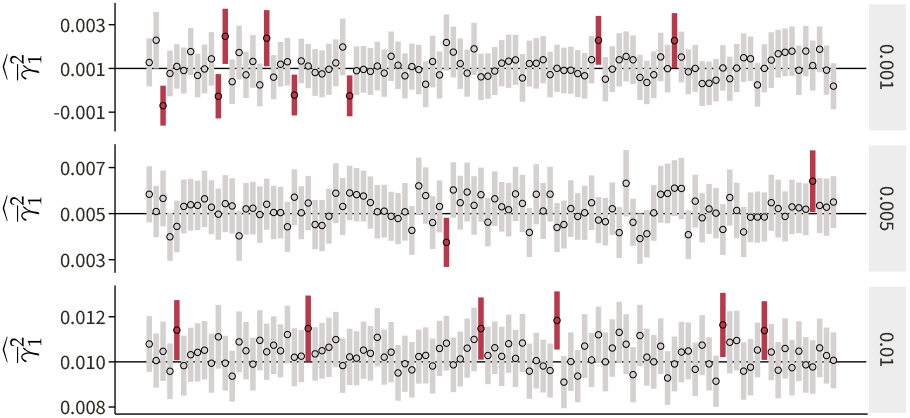
The plot shows 95% confidence intervals from 100 simulations for various values of 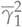. Confidence intervals in red represent realizations that do not cover the true value.

**Figure G.9.**
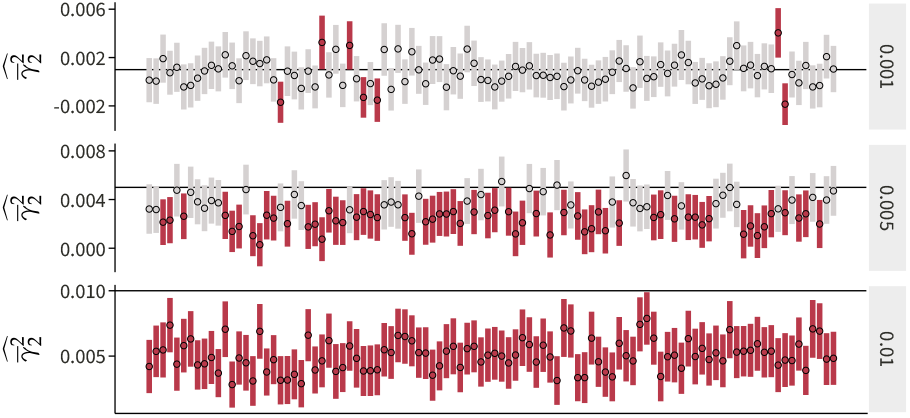
The plot shows 95% confidence intervals from 100 simulations for various values of 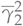. Confidence intervals in red represent realizations that do not cover the true value.

#### G.1.3 Method-of-Moments Monotonicity Estimator

Our covariance component estimators were close to unbiased (Figure G.10). We noted a slight upward bias in the LoF burden effect variance estimator 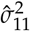. However, the magnitude of this bias was only around 5% of the true parameter value. We believe that this represents residual confounding that is not corrected for using the genotyping and rare variant PCs used in our simulations.

**Figure G.10.**
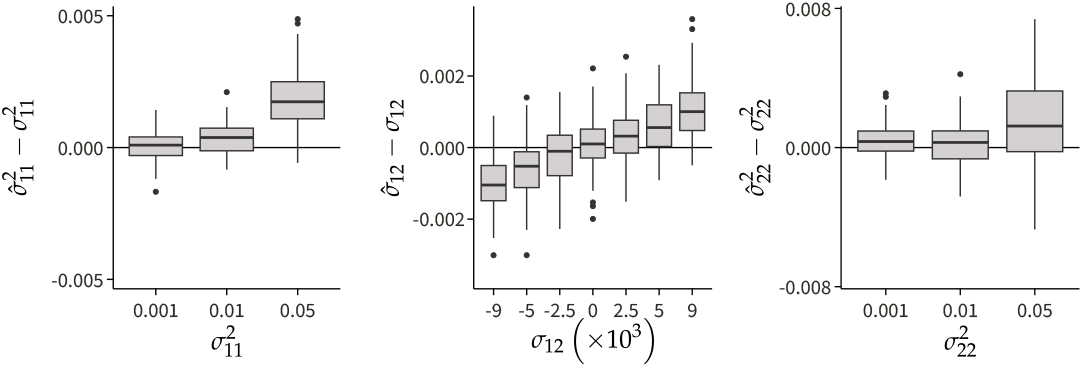
(Left) The difference in the estimated value 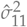 and the true value 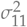 for various tested values are shown for 100 simulations. (Center) The difference in the estimated value 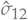 and the true value 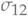 for various tested values are shown for 100 simulations. (Right) The difference in the estimated value 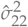 and the true value 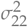 for various tested values are shown for 100 simulations.

Our MoM estimator for monotonicity is biased away from zero for extreme values of *ϕ*. (Figure G.11). The confidence intervals estimated using the bootstrap were relatively well-calibrated (Figure G.12).

**Figure G.11.**
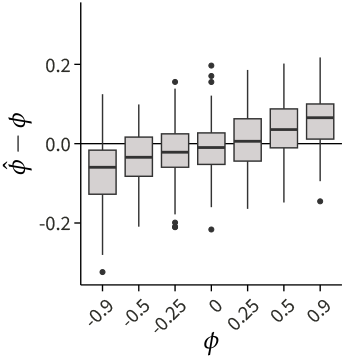
The difference in the estimated value 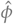 and the true value *ϕ* for various tested values are shown for 100 simulations.

**Figure G.12.**
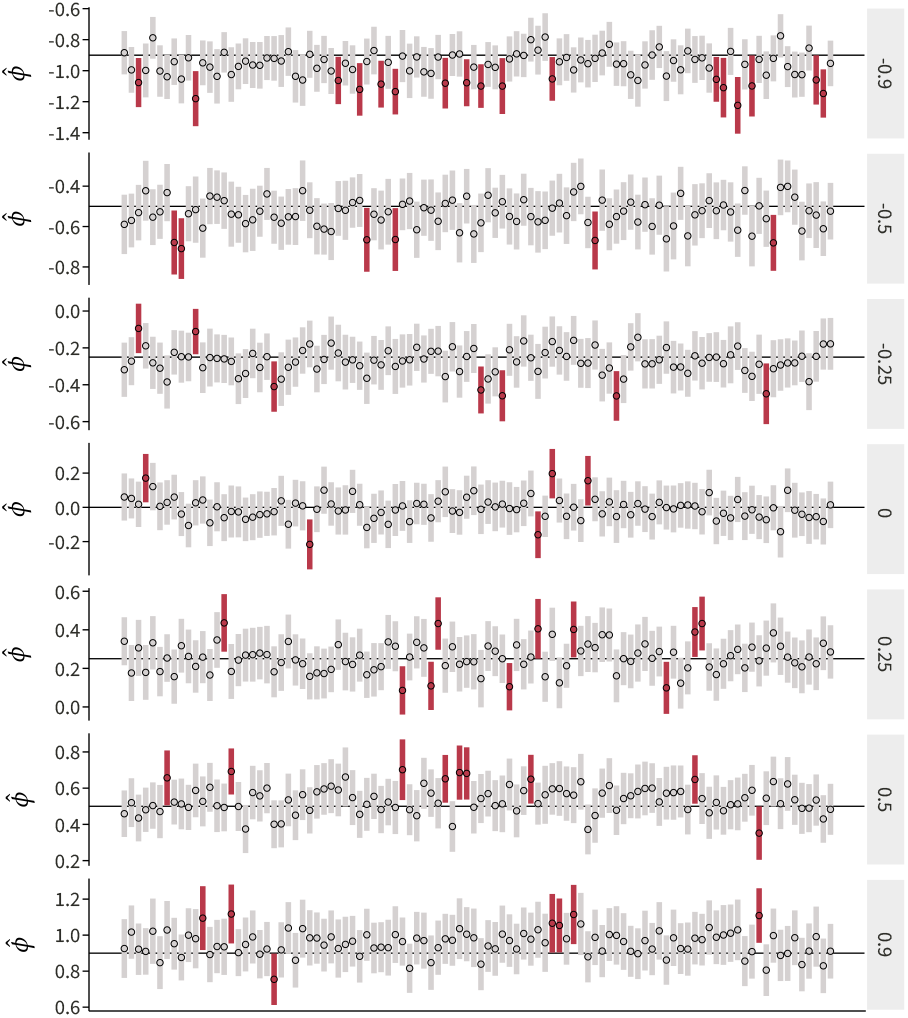
The plot shows 95% confidence intervals from 100 simulations for various values of *ϕ*. Confidence intervals in red represent realizations that do not cover the true value.

#### G.1.4 Maximum Likelihood Average Burden Effect Estimators

For the maximum likelihood estimation approach, we used random-effect simulations to test 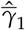 and 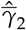. These estimators were also convincingly unbiased, similar to our unbiased estimators (Figure G.13). The approximate confidence intervals are well-calibrated (Figures G.14 and G.15).

**Figure G.13.**
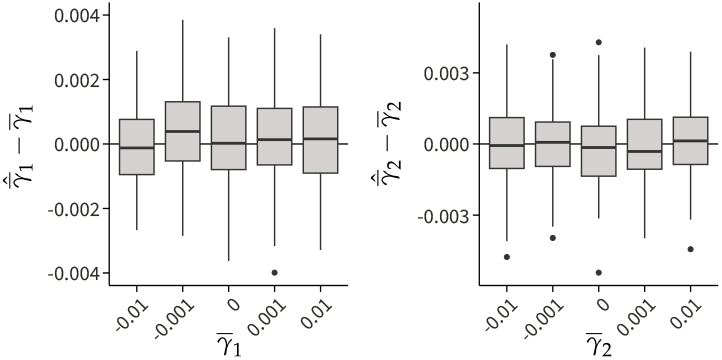
(Left) The difference in the estimated value 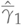 and the true value 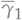 for various tested values are shown for 100 simulations. (Right) The difference in the estimated value 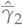 and the true value 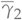 for various tested values are shown for 100 simulations.

**Figure G.14.**
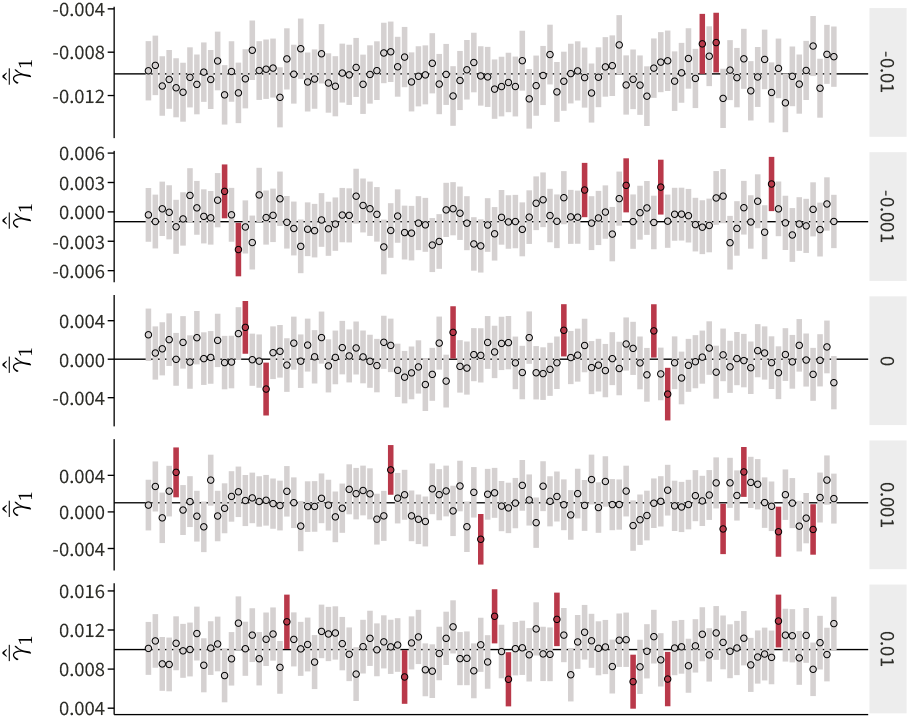
The plot shows 95% confidence intervals from 100 simulations for various values of 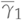. Confidence intervals in red represent realizations that do not cover the true value.

**Figure G.15.**
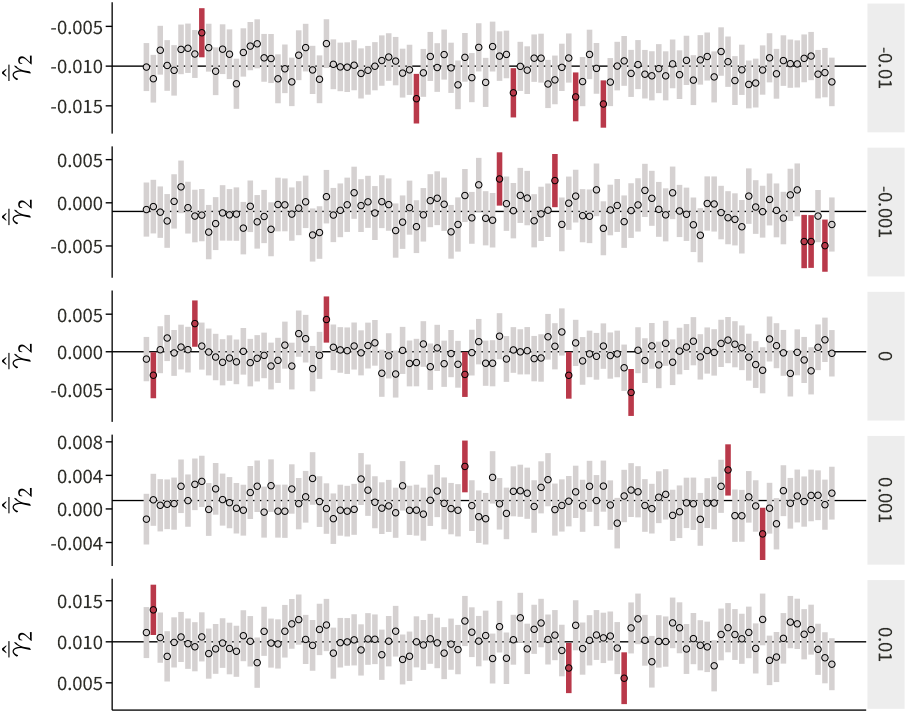
The plot shows 95% confidence intervals from 100 simulations for various values of 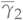. Confidence intervals in red represent realizations that do not cover the true value.

#### C.1.5 Maximum Likelihood Monotonicity Estimator

Our covariance component estimators from the maximum likelihood approach had a similar performance to the unbiased estimators (Figure G.16). We noted a slight upward bias in the LoF burden effect variance estimator, similar to the unbiased estimator. The bias was again only around 5% of the true parameter value.

**Figure G.16.**
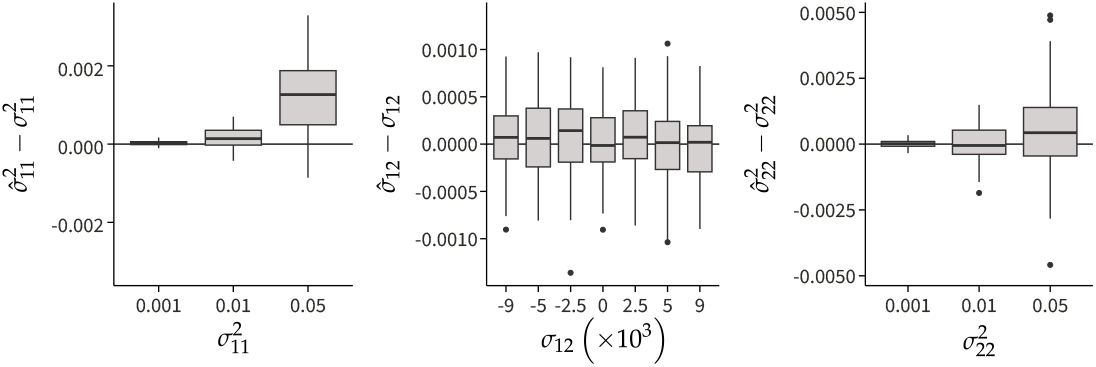
(Left) The difference in the estimated value 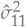 and the true value 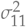 for various tested values are shown for 100 simulations. (Center) The difference in the estimated value 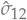 and the true value 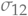 for various tested values are shown for 100 simulations. (Right) The difference in the estimated value 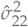 and the true value 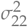 for various tested values are shown for 100 simulations.

The maximum likelihood estimate for monotonicity was slightly biased towards zero for extreme values of *ϕ* (Figure G.17). The magnitude of bias was small compared to the true value, and the approximate confidence intervals using the delta method remained well-calibrated (Figure G.18). The variance of the estimator was approximately half that of the MoM estimator (Figure G.11).

**Figure G.17.**
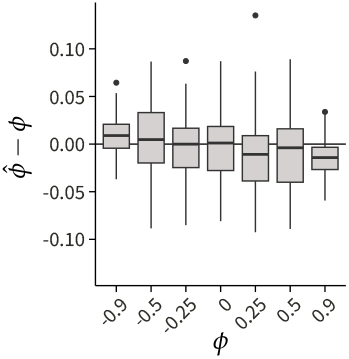
The difference in the estimated value 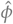 and the true value *ϕ* for various tested values are shown for 100 simulations.

**Figure G.18.**
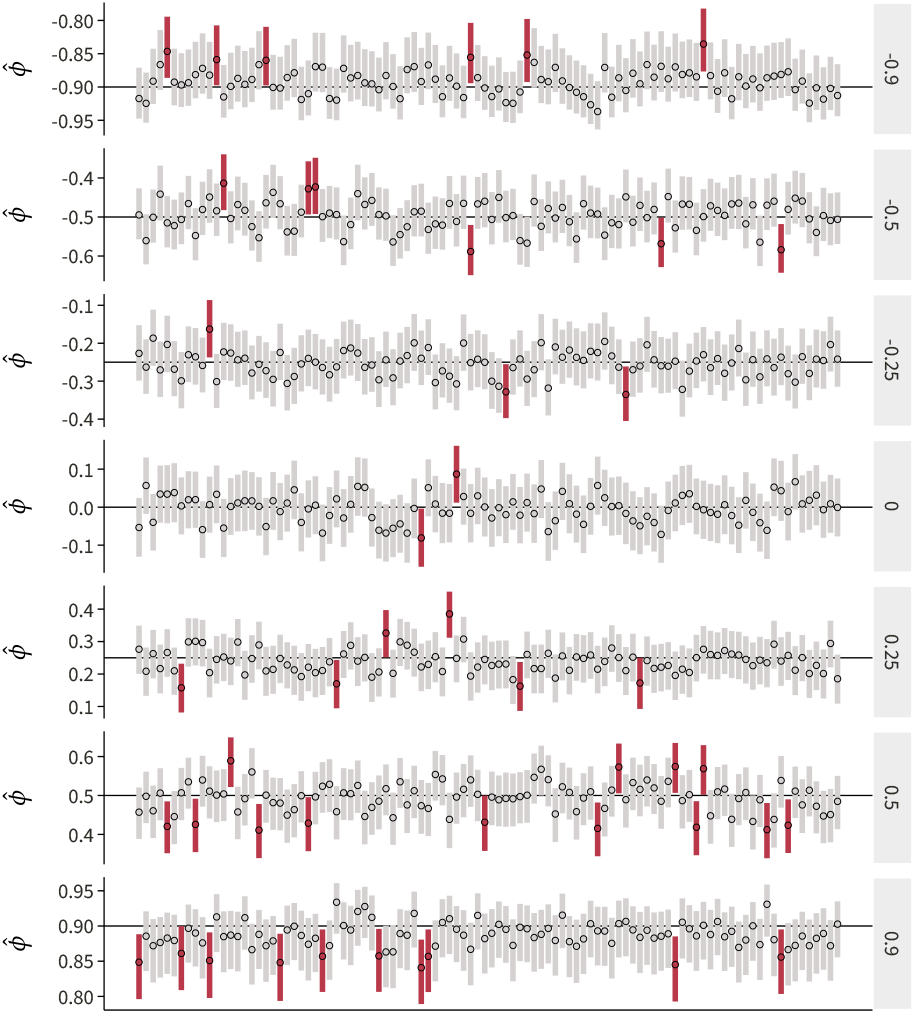
The plot shows 95% confidence intervals from 100 simulations for various values of *ϕ*. Confidence intervals in red represent realizations that do not cover the true value.

### G.2 Trait Buffering Simulations

To assess the trait buffering model, we performed three types of simulations. The goal of the first simulation was to use a realistic distribution of effect sizes and perform inference using our trait buffering model. The goal of the second and third simulations was to use an extreme distribution of effect sizes to assess the behavior of *ξ*_m_ and *ξ*_nm_.

For the first simulation, we simulated 100 traits by drawing true effect sizes from

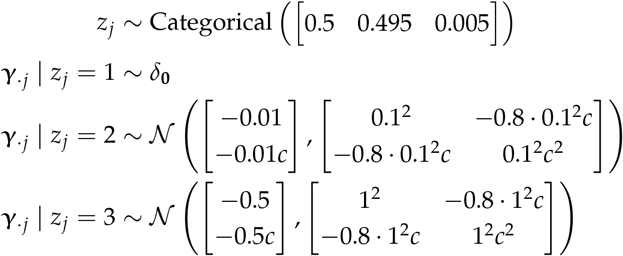

where 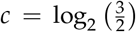 and *j* indexes over genes. Here, *δ*_**0**_ represents the Dirac measure at **0**. The first component represents true null effects, the second components represents genes with small effects, and the last component represents genes with large effects. This simulation contains both monotone and non-monotone GDRCs (Figure G.19). Negative trait buffering is simulated by using a non-zero, negative mean effect for the second and third components. We simulated monotone traits by using a correlation of -0.8 for the effect sizes.

The second and third simulations were used to evaluate the robustness of *ξ*_m_ and *ξ*_nm_, which measure the contributions of monotone and non-monotone GDRCs respectively. For the second simulation, we generated only monotone GDRCs by modifying the first simulation and using truncated distributions for the components (Figure G.19). The truncations restricted samples to the monotone quadrants. For the third simulation, we generated only non-monotone GDRCs using the same strategy as the second simulation (Figure G.19).

**Figure G.19.**
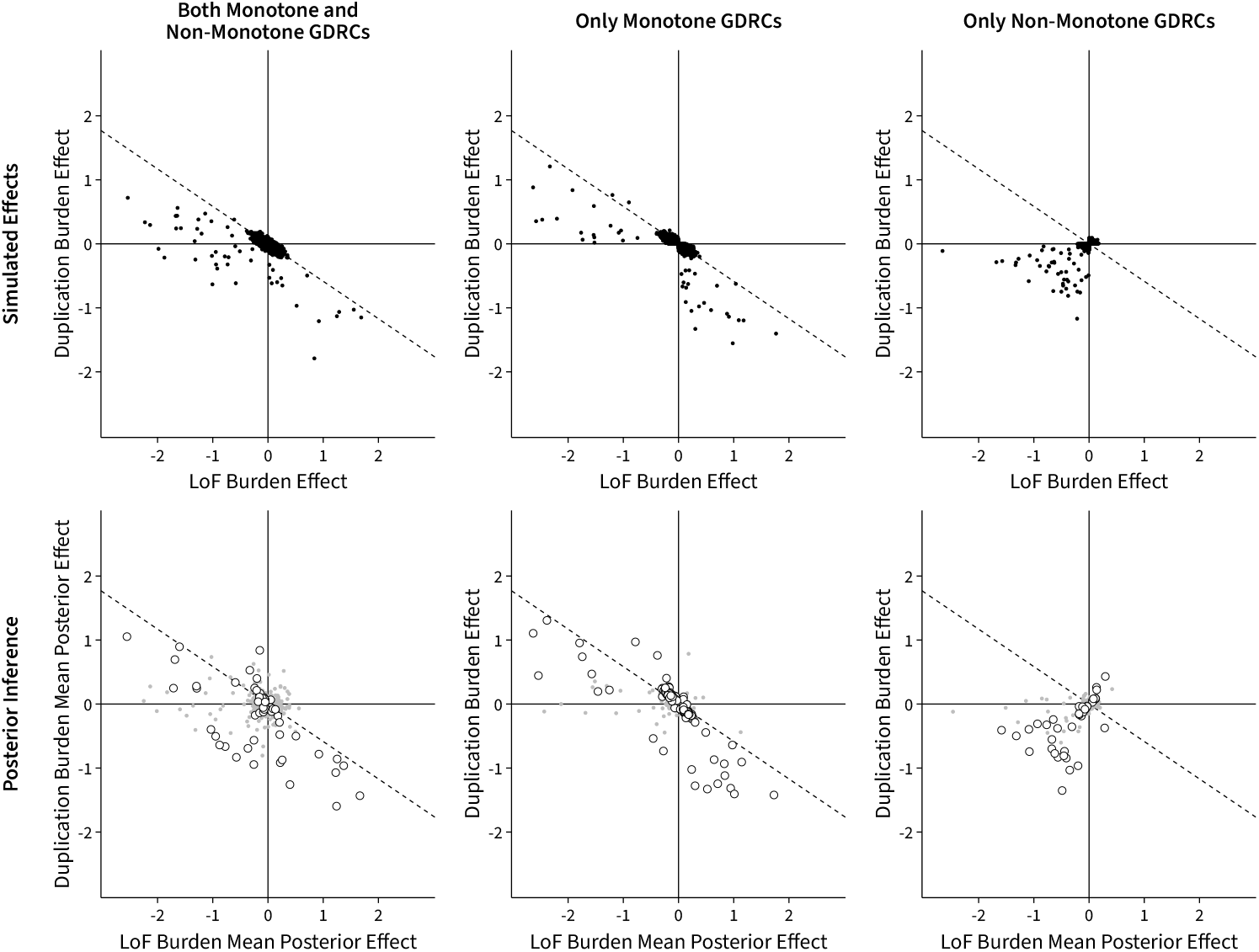
(Top) An example of simulated data for the trait buffering model, where each point represents the burden effects for a gene. The first example is simulated data with both types of GDRCs. The second example contains only monotone GDRCs. The third example contains only non-monotone GDRCs. (Bottom) Posterior mean burden effect estimates from the trait buffering model for the simulated data. The large white points represent genes where the maximum LFSR is less than 0.1.

In general, we observed that the mean posterior effect sizes from our inference framework recovered the trait-level distribution of effect sizes, even under the extreme examples of the truncated distributions (Figure G.19). At the gene level, we were faithfully able to recover the simulated burden effect sizes (Figure G.20).

**Figure G.20.**
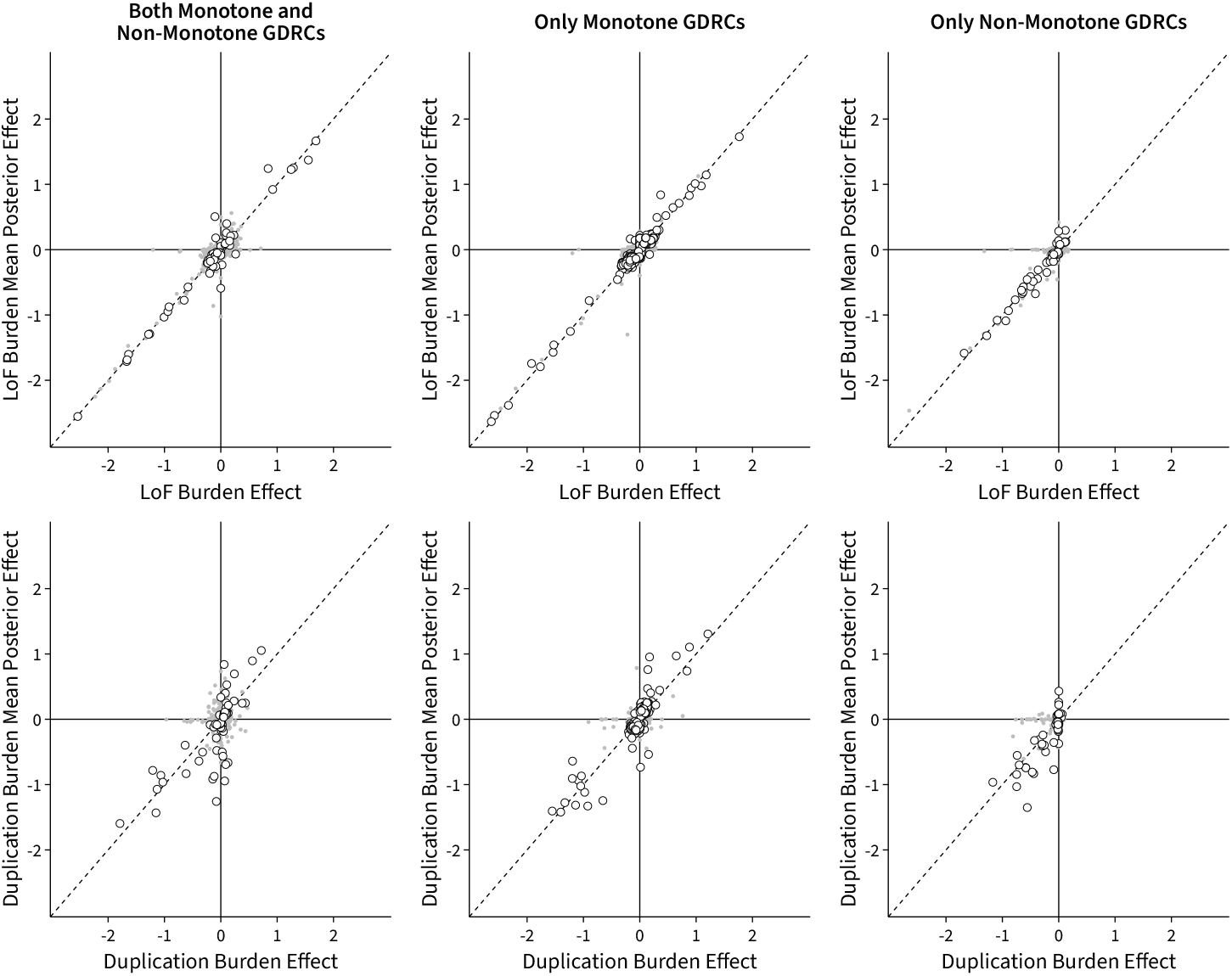
Each column represents one simulated trait. The horizontal axes represent the true burden effect from simulation, and the vertical axes represent the inferred mean posterior effect under the trait buffering model. The large white points represent genes where the maximum LFSR is less than 0.1.

#### G.2.1 Estimated Trait Buffering

We used the first simulation to assess the model’s performance at estimating *ξ*. As expected, the estimates were all negative, in line with the negative trait buffering that was assumed for the simulation. The estimates are shrunk towards zero, which is expected since the expected value of *ξ* is zero under the prior.

**Figure G.21.**
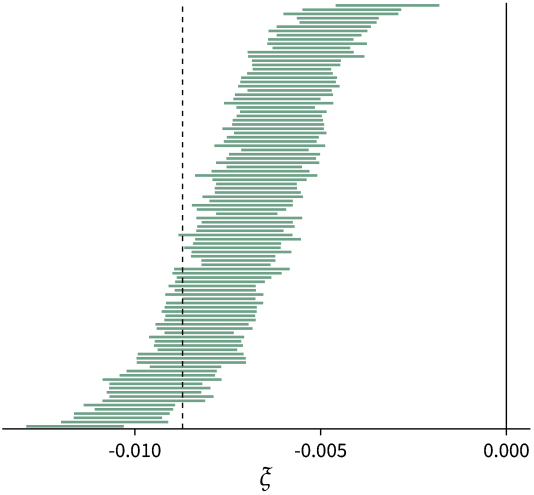
The 95% credible intervals for *ξ* from the trait buffering model. The 100 traits from the first simulation were used for this assessment. The dotted line represents the expected value of *ξ* for the simulated latent effect sizes.

#### G.2.2 Component Measures are Biased

From the trait buffering model, we derived two statistical measures of the contribution to *ξ* that comes from monotone and non-monotone GDRCs. Recall that this decomposition is

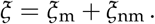

Consider the method to estimate one of these components, *ξ*_nm_, which is the non-monotone component. The statistic is calculated by aggregating all posterior samples for ***γ*** that fall into the non-monotone quadrants. However, if trait buffering is occurring only due to monotone GDRCs, any uncertainty in their sign results in sampling from the non-monotone quadrants in a biased manner, introducing bias in the estimated component. This effect can be seen in Figure G.22, where we estimated the monotone and non-monotone component from our simulation with only monotone GDRCs. The non-monotone component is substantially biased due to the uncertainty in the monotone component.

**Figure G.22.**
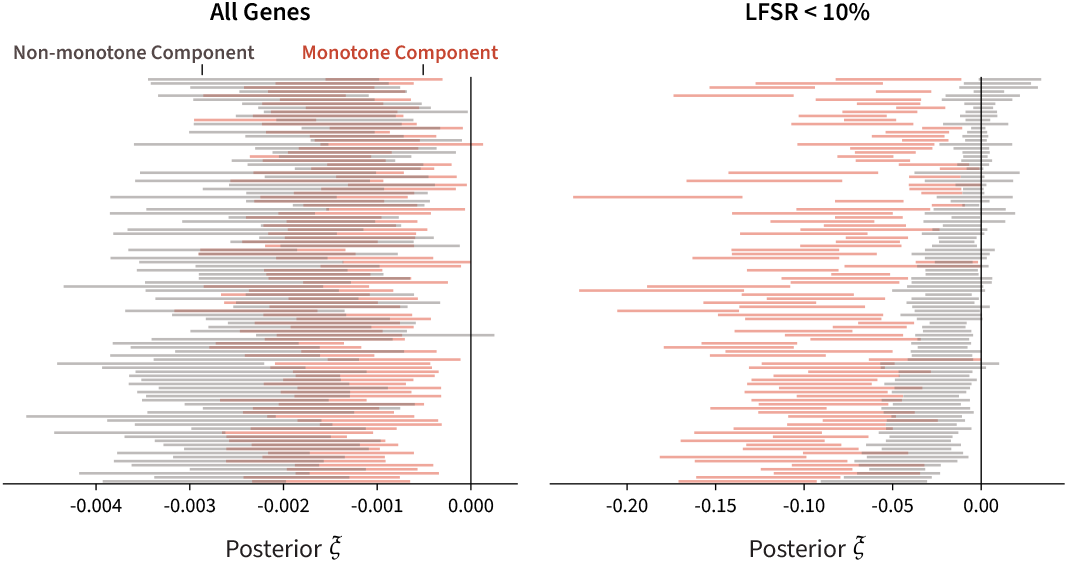
Estimates of the monotone and non-monotone component of *ξ* for the 100 simulated traits with only monotone GDRCs. (Left) These components were estimated across all genes. (Right) These components were estimated for all genes with a local false sign rate of less than 10% for both LoF and duplication burden effects.

We expect this effect to be ameliorated when we are more certain in the sign of effect, since fewer samples will fall in the incorrect quadrant. Indeed, the signal from the monotone GDRCs reveals itself when we subset to samples from genes with local false sign rate (LFSR) less than 10% (Figure G.22). A similar bias is expected and observed in the monotone component if a trait is driven only by non-monotone GDRCs (Figure G.23), although it is much less severe.

**Figure G.23.**
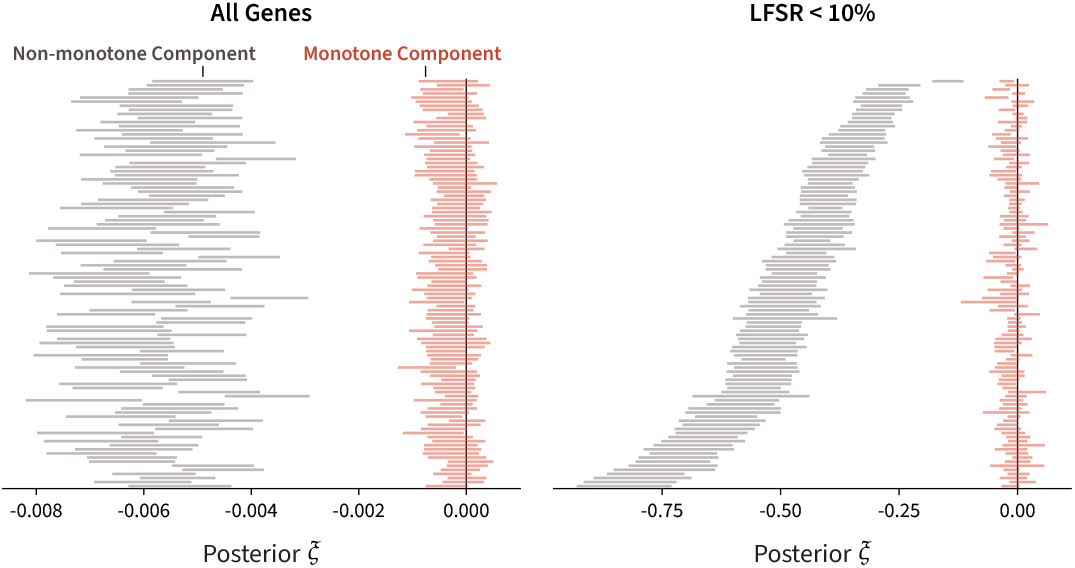
Estimates of the monotone and non-monotone component of *ξ* for the 100 simulated traits with only non-monotone GDRCs. (Left) These components were estimated across all genes. (Right) These components were estimated for all genes with a local false sign rate of less than 10% for both LoF and duplication burden effects.

These experiments suggest that it is challenging to estimate the absolute proportion of contribution from monotone or non-monotone GDRCs to *ξ* due to the geometry of the problem. However, in the simulations with only monotone GDRCs, the monotone component is larger than the non-monotone component when we restrict to genes with a LFSR less than 10% (Figure G.22). When we perform the same analysis with the observed data, we see that the non-monotone component remains larger than the monotone component after restricting to genes with LFSR less than 10% for many of the traits. Given the simulation results above, this suggests that non-monotone GDRCs are contributing to the trait buffering in the observed data.

**Figure G.24.**
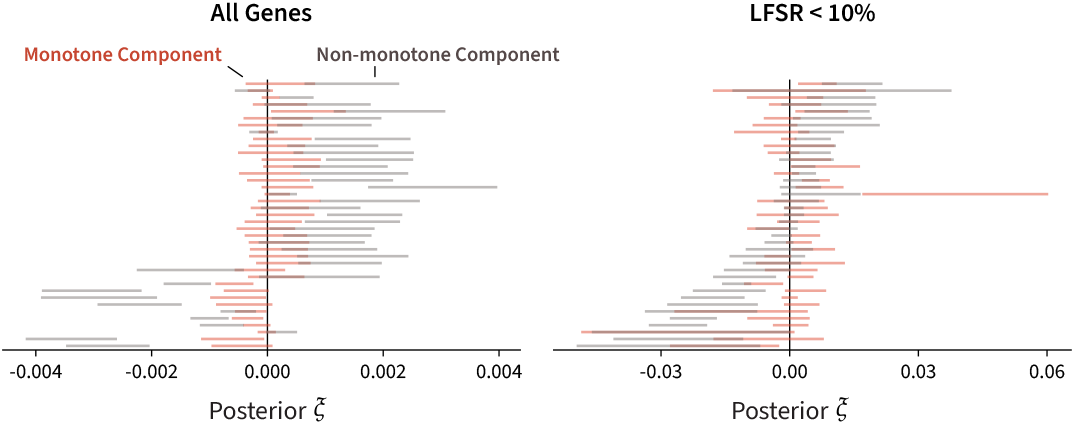
Estimates of the monotone and non-monotone component of *ξ* for the traits with a nominally significant non-monotone aGDRC. These are the same traits pictured in Figure 5. (Left) These components were estimated across all genes. (Right) These components were estimated for all genes with a local false sign rate of less than 10% for both LoF and duplication burden effects.

## H Gene Burden Tests

### H.1 Synonymous Variants

Stratification or other confounding can inflate the estimated effect sizes from association analyses. We can ensure that we are adequately controlling for confounding by estimating the effect of synonymous variants, which are expected to not affect protein function and downstream traits.

We used synonymous variants to test the effect of using different subsets of individuals. The original analysis of LoF variants in the UKB [23] used a subset of around 430K individuals with genetic similarity to the EUR superpopulation from the 1000 Genomes Project. In addition to a population with genetic similarity to EUR (called EUR for brevity), we tested the subset of 390K unrelated individuals in the EUR subset (called unrelated) and the subset of all 460K individuals with WES data (called WES). This allowed us to determine if relatedness or population stratification inflated our estimates of effect size. The EUR population was defined using self-reported information and boundaries in genotyping PC space from prior genetic analysis in the UKB [24]: −20 ≤ PC1 ≤ 40 and −25 ≤ PC2 ≤ 10 (Array items 1 and 2 from field 22009 in the UKB) for either self-identified “White British” or self-identified “non-British White” (Field 21000 in the UKB).

Additionally, we also used synonymous variants to test the effect of using different numbers of genotyping PCs. The original analysis used 10 genotyping PCs when performing association analyses within populations with high genetic similarity [23]. In addition to 10 genotyping PCs, we tested 15 and 20 genotyping PCs since we planned to use as many individuals in the UKB as possible, which might introduce additional confounding due to population stratification.

To test for stratification, we ran synonymous variant burden tests on a subset of nine continuous traits: height, body mass index (BMI), low-density lipoprotein cholesterol (LDL-C), mean corpuscular hemoglobin (MCH), red blood cell distribution width (RDW), forced vital capacity (FVC), creatinine, cystatin C, and the north coordinate of the place of birth in the United Kingdom (NC). Strong effects for NC should be a good measure of uncontrolled stratification.

#### H.1.1 Number of Genotyping Principal Components

Synonymous variants are expected to not have any effect on traits. Therefore, the mean squared effect of synonymous variant burden associations should provide an estimate of inflation in effect sizes due to other sources of confounding. The gold standard for genetic association analysis is using the cohort of unrelated individuals with high genetic similarity. Compared to this cohort, the amount of inflation was indistinguishable in the EUR and WES cohorts (Figure H.1). Thus, we decided to use the WES cohort to maximize our sample size.

**Figure H.1.**
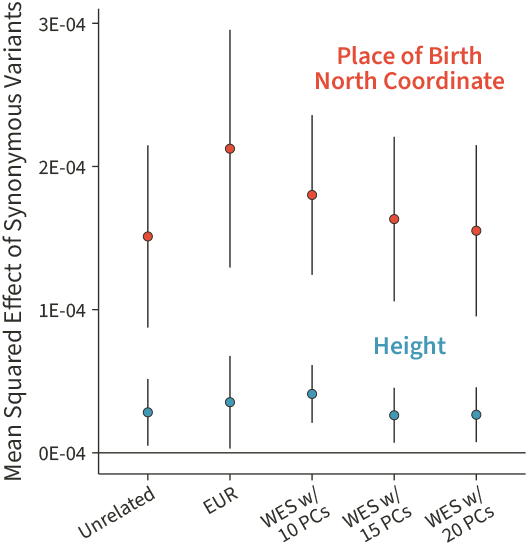
The mean squared effect of synonymous variants for various cohorts of individuals and different numbers of genotyping PCs. The unrelated and EUR cohort burden tests used 15 genotyping PCs. We tested 10, 15, and 20 genotyping PCs in the WES cohort burden tests.

The effect of including 10 genotyping PCs was also indistinguishable from 15 or 20 genotyping PCs (Figure H.1). The original analysis included 10 genotyping PCs [23], but performed analyses in cohorts of high genetic similarity. Since we were including all individuals in the UKB, we decided to conservatively use 15 genotyping PCs.

#### H.1.2 Inflation from Confounding

Since we detected a significant mean squared effect for NC, we were concerned about inflation of effect sizes due to confounding. To test the effect of this inflation, we compared the mean squared effect of LoF variant burden associations with the mean squared effect of synonymous variant burden associations across various traits. We noted that the mean squared effect of LoF burden associations was an order of magnitude larger than the effect of synonymous burden associations (Figure H.2). Thus, we concluded that although confounding is likely present in our association tests, the signal is at least 10 times greater than the bias.

**Figure H.2.**
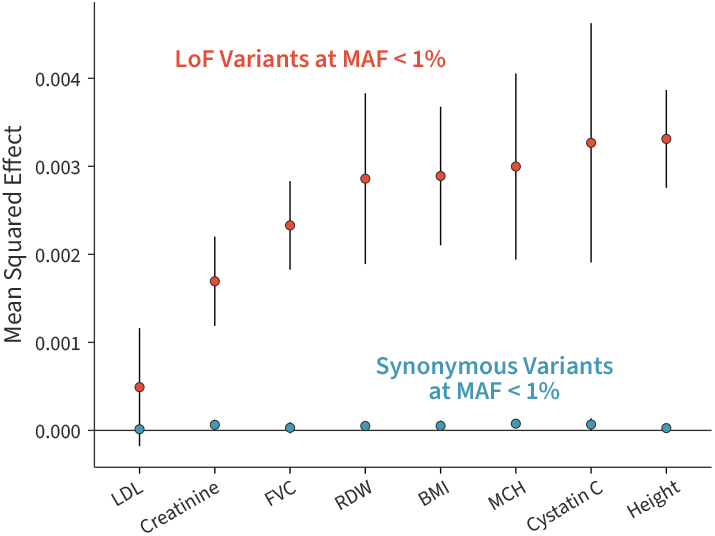
LoF variants have a magnitude larger effect on traits than synonymous variants across various continuous traits.

#### H.1.3 Utility of Misannotation Probability

**Figure H.3.**
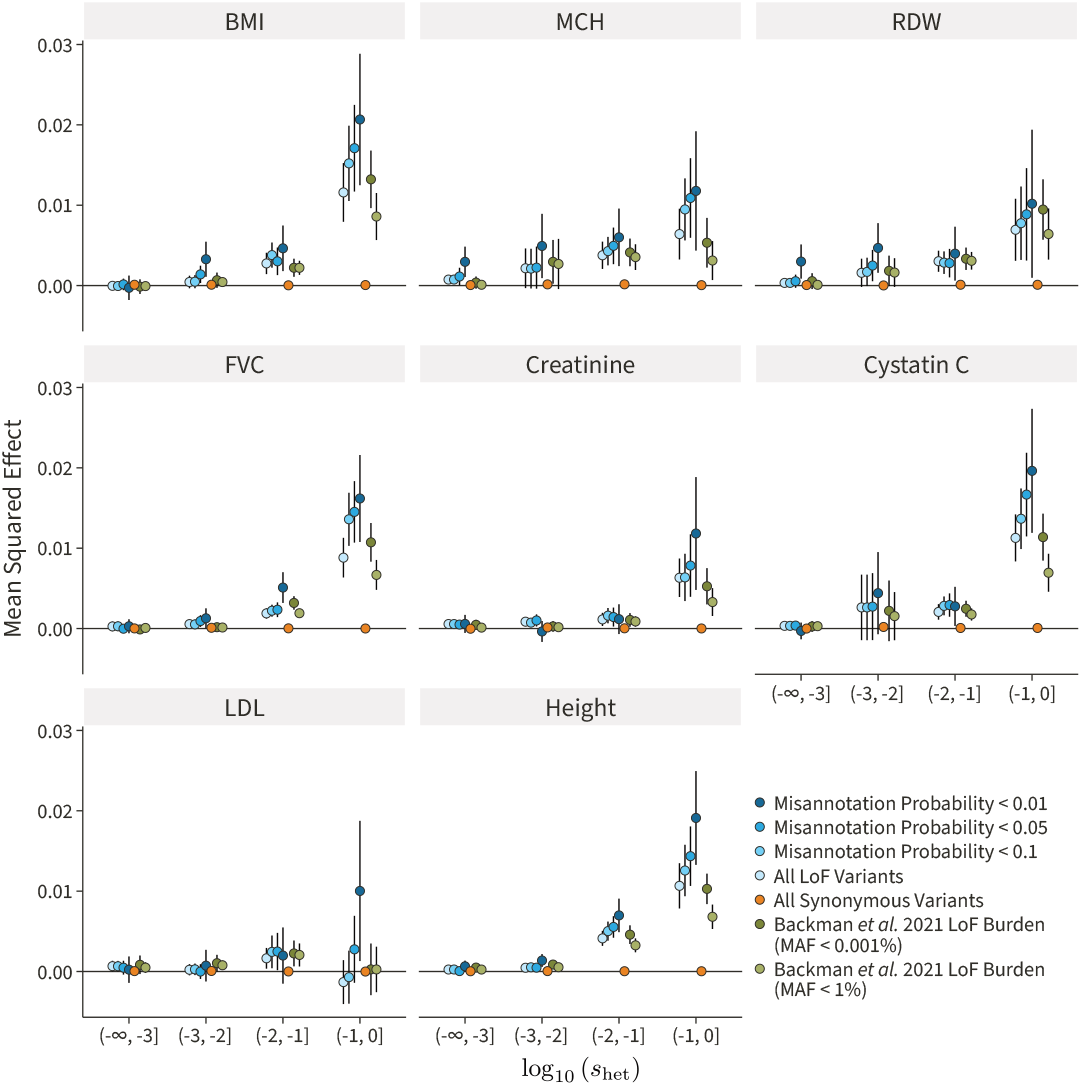
The mean squared effect of genes in various buckets of log_10_ (*s*_het_) for various traits. Genes with larger effects tend to be more constrained. All data here uses a MAF < 1% filter other than the MAF < 0.001% associations from [23]. The use of the misannotation probability increases the signal detected. The signal in the original analysis of the LoF data is also shown for reference. 95% confidence intervals are displayed for each estimate.

Burden tests often use various maximum minor allele frequency (MAF) filters. For instance, Backman et al. used MAF filters of 1%, 0.1%, 0.01%, and 0.001% [23]. Presumably, such filters assume that increasingly stringent filters will reduce the number of false positive LoF variants that are aggregated into the burden genotype, as LoF variants that are at high frequency in the population might represent misannotated LoF variants. However, such filters result in asymmetric behavior across genes depending on their constraint. For example, the highly stringent 0.001% filter will reduce the false-positive rate in highly constrained genes, but will remove true LoF variants in unconstrained genes. Ideally, a gene under high constraint should use a stringent MAF filter, while a gene under low constraint should use a liberal MAF filter.

**Figure H.4.**
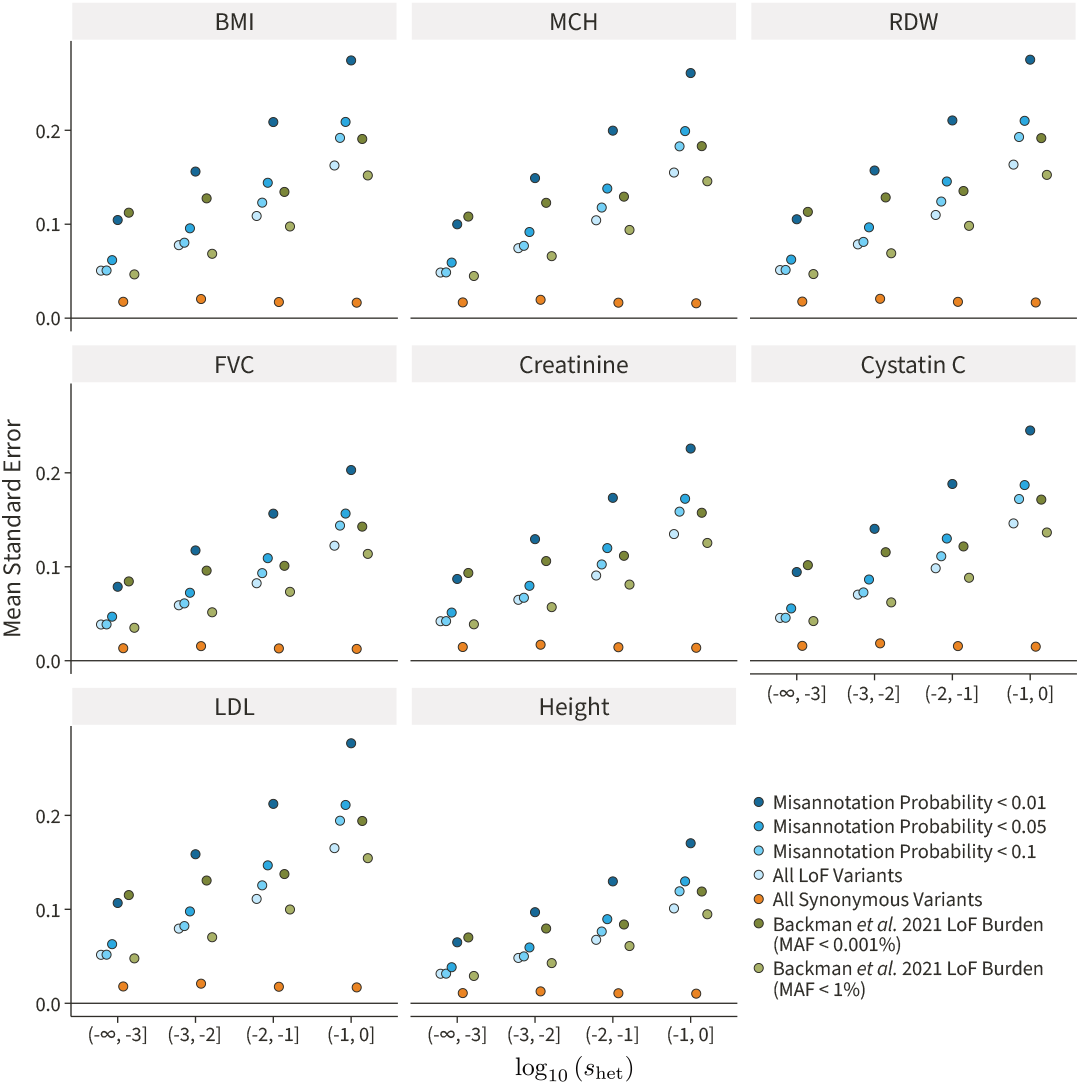
The mean standard error of burden estimates for genes in various buckets of log_10_ (*s*_het_) for various traits. Since larger-effect variants are under increased selective constraint, their frequencies are lower and the estimation noise is larger. All data here uses a MAF < 1% filter other than the MAF < 0.001% associations from [23]. The use of the misannotation probability increases the noise of the estimates. The noise in the original analysis of the LoF data is also shown for reference. 95% confidence intervals are present on the plots but not visible due to their small length.

To account for this, Zeng et al. have calculated misannotation probabilities for all potential LoF-introducing SNPs in genes for which they had estimated *s*_het_ values [25]. We used various misannotation probability filters instead of MAF filters. We tested filters of 10%, 5%, and 1% mis-annotation probability for all variants with MAF < 1%. We estimated the total signal, as measured by the mean squared effect size, in various buckets of gene constraint (Figure H.3). Increasingly stringent misannotation probability filters increased the signal across various selection buckets. In addition, the various filters were as or more effective than both the 1% and 0.001% MAF filters from [23].

Increasing stringency with the misannotation probability filters does increase the amount of estimation noise in 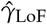 as more LoF variants are removed from the burden genotypes (Figure H.4). We found that a misannotation probability filter of 10% provided signal comparable to or better than a MAF filter of 0.001% with a minimal increase in noise. This filter was used in all subsequent analyses.

### H.2 CNV Genotyping Error

When we first estimated the correlation matrices for the duplication burden genotypes, we noticed large amounts of long-range correlations (Figure H.5).

**Figure H.5.**
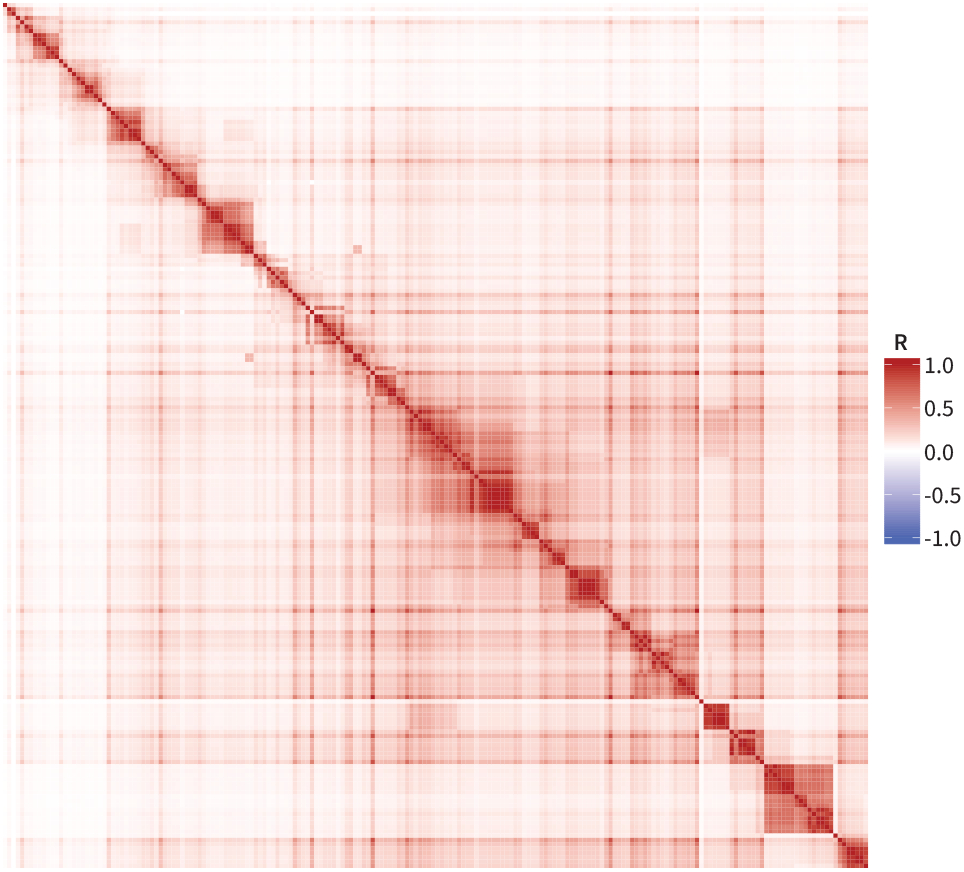
The correlation matrix for the first 200 genes on chromosome 3 using all the samples. Although segregating duplications can be large, they are not expected to capture so many genes.

We reasoned that this is driven by a few poor-quality samples with systematically higher copy number. In their initial report on the UKB WGS data, The UK Biobank Whole-Genome Sequencing Consortium et al. reported an average structural variant (SV) burden of 3.6 Mbp per haploid genome [26]. Assuming that the total CNV burden is on a similar order of magnitude, we decided to exclude any samples with more than 10 Mbp of either deletion or duplication burden. Most samples had less than 10 Mbp of duplications or deletions. However, some samples had upwards of 100 Mbp of affected sequence (Figure H.6).

**Figure H.6.**
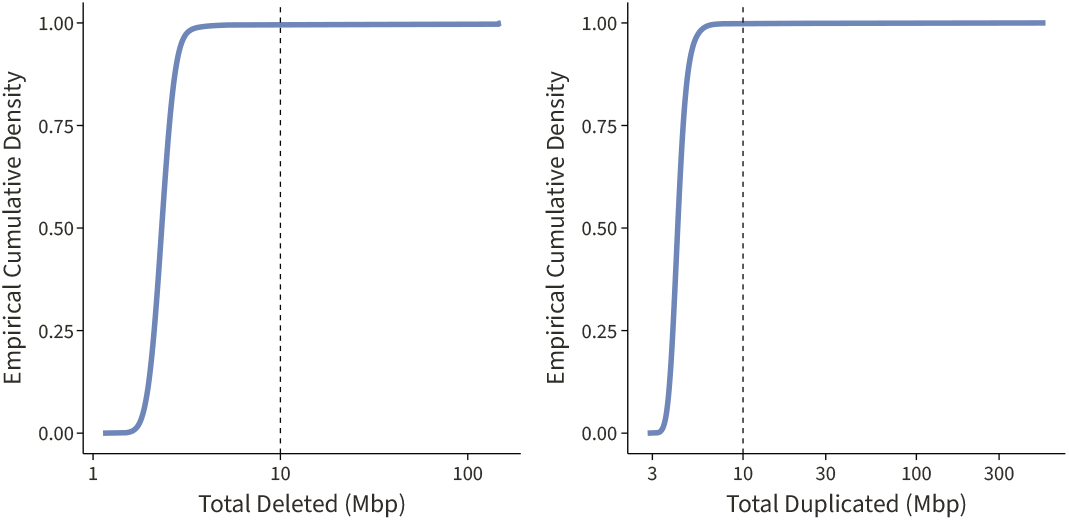
The empirical cumulative density function (eCDF) of the total amount of genome sequence affected by either deletion or duplication. Our 10 Mbp filtering step impacted only a few samples.

Filtering out 2,674 samples with deletion or duplication burden greater than 10 Mbp resulted in more reasonable duplication correlation estimates (Figure H.7).

**Figure H.7.**
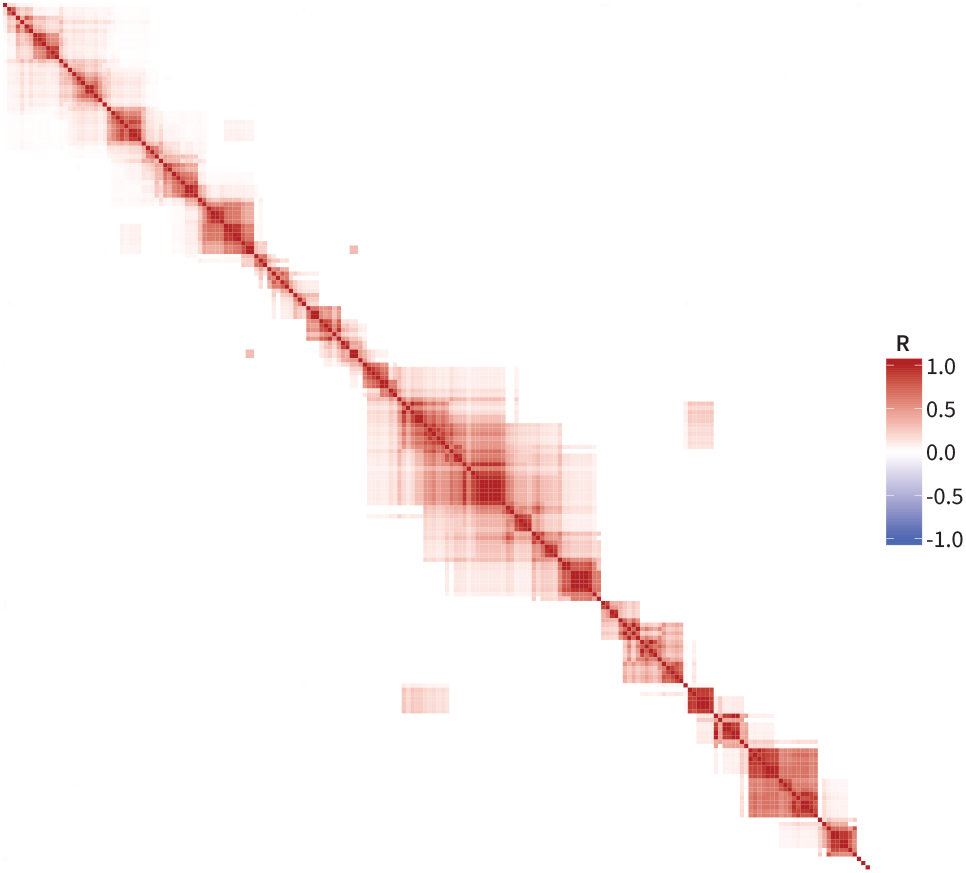
The correlation matrix for the first 200 genes on chromosome 3 using the filtered set of samples.

### H.3 Estimating CNV Correlation Matrices

Multiple covariates need to be accounted for when performing burden tests. Suppose that **Z**_*c*_ *∈* R^*N×C*^ represents the covariate values for the duplication burden tests for *C* covariates. The burden test model used to derive summary statistics in REGENIE is

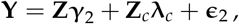

where ***λ***_*c*_ *∈* ℝ^*C*^ are the effects of the covariates on the phenotype. To remove these effects before estimating the correlation matrix, we partial out the covariates by projecting the burden genotypes into the subspace orthogonal to the column space of the covariates. Specifically, we define

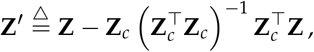

which removes the effect of the covariates by removing their contribution to the burden genotypes using the projection matrix 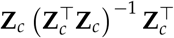. Then, we estimate the correlation matrix using these augmented burden genotypes **Z**^*′*^. The same procedure is used for the deletion burden genotype correlation matrix.

## I Data Analysis

### I.1 Significant Average Effects

**Table I.1.**
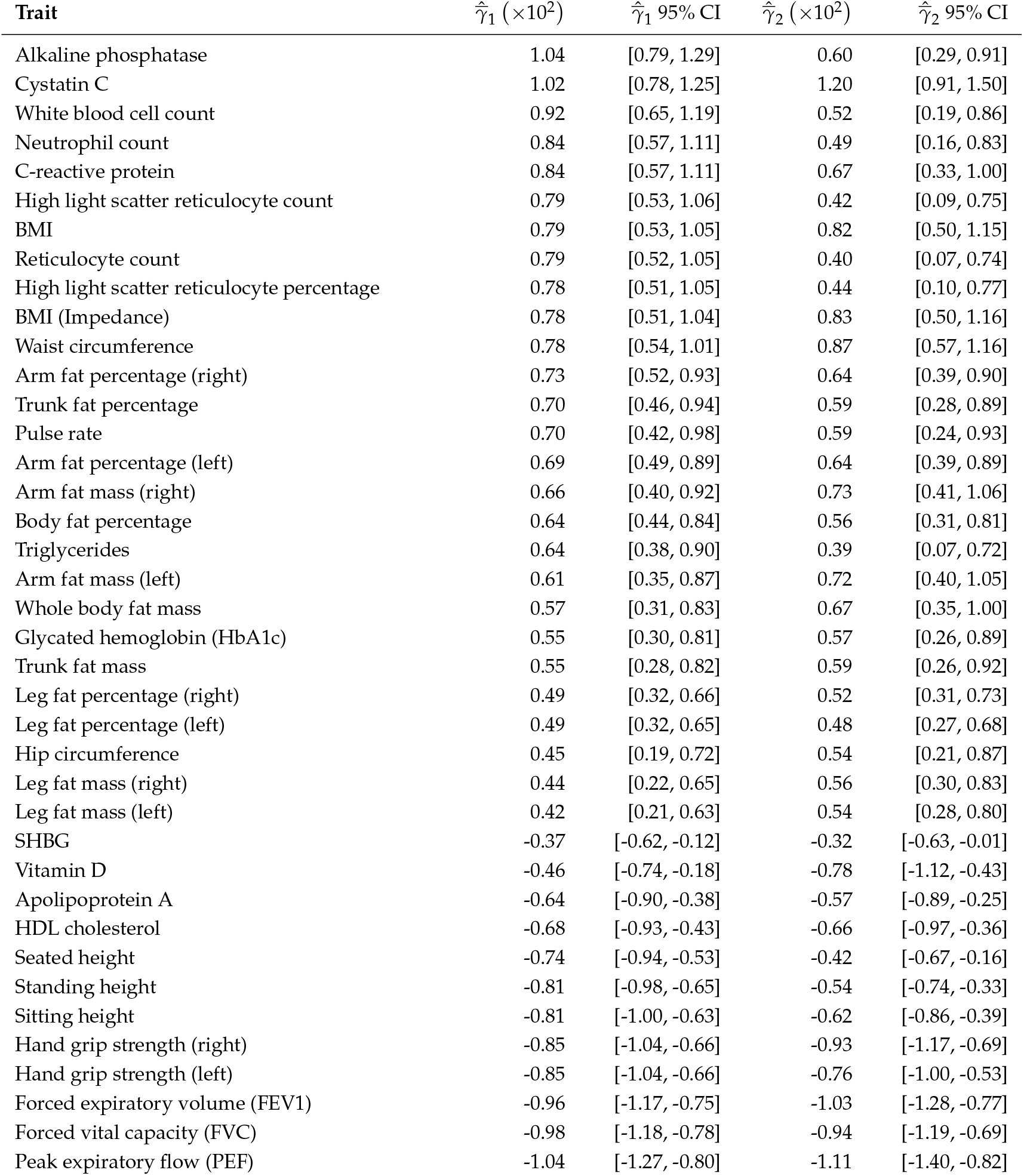
Traits with significant average burden effects were ascertained at an *α* = 0.05 level. All such traits have non-monotone average effects. These estimates are from unbiased estimators with exact confidence intervals.

| Trait | $\hat{\gamma}_1 (\times 10^2)$ | $\hat{\gamma}_1$ 95% CI | $\hat{\gamma}_2 (\times 10^2)$ | $\hat{\gamma}_2$ 95% CI |
| --- | --- | --- | --- | --- |
| Alkaline phosphatase | 1.04 | [0.79, 1.29] | 0.60 | [0.29, 0.91] |
| Cystatin C | 1.02 | [0.78, 1.25] | 1.20 | [0.91, 1.50] |
| White blood cell count | 0.92 | [0.65, 1.19] | 0.52 | [0.19, 0.86] |
| Neutrophil count | 0.84 | [0.57, 1.11] | 0.49 | [0.16, 0.83] |
| C-reactive protein | 0.84 | [0.57, 1.11] | 0.67 | [0.33, 1.00] |
| High light scatter reticulocyte count | 0.79 | [0.53, 1.06] | 0.42 | [0.09, 0.75] |
| BMI | 0.79 | [0.53, 1.05] | 0.82 | [0.50, 1.15] |
| Reticulocyte count | 0.79 | [0.52, 1.05] | 0.40 | [0.07, 0.74] |
| High light scatter reticulocyte percentage | 0.78 | [0.51, 1.05] | 0.44 | [0.10, 0.77] |
| BMI (Impedance) | 0.78 | [0.51, 1.04] | 0.83 | [0.50, 1.16] |
| Waist circumference | 0.78 | [0.54, 1.01] | 0.87 | [0.57, 1.16] |
| Arm fat percentage (right) | 0.73 | [0.52, 0.93] | 0.64 | [0.39, 0.90] |
| Trunk fat percentage | 0.70 | [0.46, 0.94] | 0.59 | [0.28, 0.89] |
| Pulse rate | 0.70 | [0.42, 0.98] | 0.59 | [0.24, 0.93] |
| Arm fat percentage (left) | 0.69 | [0.49, 0.89] | 0.64 | [0.39, 0.89] |
| Arm fat mass (right) | 0.66 | [0.40, 0.92] | 0.73 | [0.41, 1.06] |
| Body fat percentage | 0.64 | [0.44, 0.84] | 0.56 | [0.31, 0.81] |
| Triglycerides | 0.64 | [0.38, 0.90] | 0.39 | [0.07, 0.72] |
| Arm fat mass (left) | 0.61 | [0.35, 0.87] | 0.72 | [0.40, 1.05] |
| Whole body fat mass | 0.57 | [0.31, 0.83] | 0.67 | [0.35, 1.00] |
| Glycated hemoglobin (HbA1c) | 0.55 | [0.30, 0.81] | 0.57 | [0.26, 0.89] |
| Trunk fat mass | 0.55 | [0.28, 0.82] | 0.59 | [0.26, 0.92] |
| Leg fat percentage (right) | 0.49 | [0.32, 0.66] | 0.52 | [0.31, 0.73] |
| Leg fat percentage (left) | 0.49 | [0.32, 0.65] | 0.48 | [0.27, 0.68] |
| Hip circumference | 0.45 | [0.19, 0.72] | 0.54 | [0.21, 0.87] |
| Leg fat mass (right) | 0.44 | [0.22, 0.65] | 0.56 | [0.30, 0.83] |
| Leg fat mass (left) | 0.42 | [0.21, 0.63] | 0.54 | [0.28, 0.80] |
| SHBG | -0.37 | [-0.62, -0.12] | -0.32 | [-0.63, -0.01] |
| Vitamin D | -0.46 | [-0.74, -0.18] | -0.78 | [-1.12, -0.43] |
| Apolipoprotein A | -0.64 | [-0.90, -0.38] | -0.57 | [-0.89, -0.25] |
| HDL cholesterol | -0.68 | [-0.93, -0.43] | -0.66 | [-0.97, -0.36] |
| Seated height | -0.74 | [-0.94, -0.53] | -0.42 | [-0.67, -0.16] |
| Standing height | -0.81 | [-0.98, -0.65] | -0.54 | [-0.74, -0.33] |
| Sitting height | -0.81 | [-1.00, -0.63] | -0.62 | [-0.86, -0.39] |
| Hand grip strength (right) | -0.85 | [-1.04, -0.66] | -0.93 | [-1.17, -0.69] |
| Hand grip strength (left) | -0.85 | [-1.04, -0.66] | -0.76 | [-1.00, -0.53] |
| Forced expiratory volume (FEV1) | -0.96 | [-1.17, -0.75] | -1.03 | [-1.28, -0.77] |
| Forced vital capacity (FVC) | -0.98 | [-1.18, -0.78] | -0.94 | [-1.19, -0.69] |
| Peak expiratory flow (PEF) | -1.04 | [-1.27, -0.80] | -1.11 | [-1.40, -0.82] |

We estimated average burden effects using unbiased estimators. Traits with significant LoF and duplication burden effects were ascertained at an *α* = 0.05 level (Table I.1).

### I.2 Lack of Average Effect in Genome-Wide Association Studies

For traits with significant average burden effects (Table I.1), we downloaded genome-wide association study (GWAS) summary statistics from the Neale Lab (https://www.nealelab.is/uk-biobank, version 3, both sexes) based on 337,199 White British individuals in the UKB. In their processing pipeline, phenotypic values were inverse-rank normal transformed. Age, age squared, inferred sex, age-by-sex, age-squared-by-sex, and 20 genotyping PCs were included as covariates.

We estimated conditionally independent hits and their effect sizes using GCTA-COJO [27] with parameters `--cojo-p 5e-8 --cojo-slct`. We used the genotypes of 10,000 unrelated White British individuals in the UKB to compute the LD reference panel. We polarized significant trait-associated variants into ancestral and derived states with variation features obtained from the Ensembl Variation database [28].

**Figure I.1.**
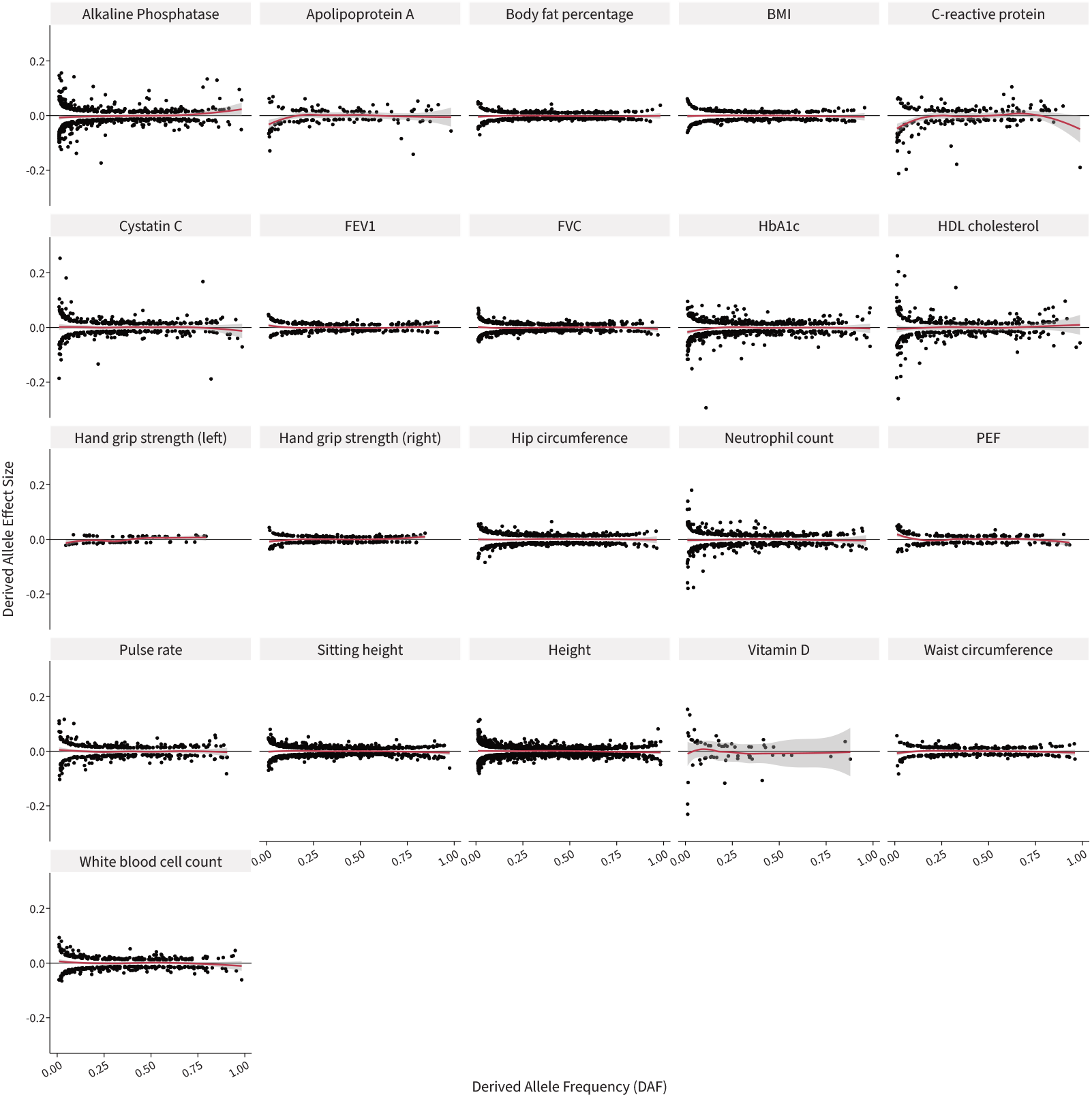
Trumpet plots of the derived allele effect size versus the derived allele frequency for top genome-wide association study (GWAS) associations for traits with significant loss-of-function (LoF) and duplication average burden effects.

In the trumpet plots of derived allele effect versus derived allele frequency (Figures 1B and I.1), we use top significant conditionally independent hits per locus detected in a GWAS. One concern in such a plot is that some top hits may be tagging SNPs, and the causal SNPs may be in negative or positive LD with the tagging SNPs. Tagging SNPs in negative LD with the causal SNP can mask an underlying true non-zero average effect.

To address these concerns, we point to recent work done to understand selection using common variation [29]. Under neutral coalescent simulations, the causal and tagging SNPs are expected to be in positive LD with high probability for derived allele frequencies in the interval [0, 0.5] as shown in Figure S14 of [29]. The trumpet plot continues to display a nearly zero average effect in this interval, suggesting that the underlying causal SNPs also do not show an average effect on the trait. We estimated the proportion of trait-increasing variants in the interval [0, 0.5] using a binomial test, and found no evidence for a bias towards one direction for any trait (Figure I.2).

**Figure I.2.**
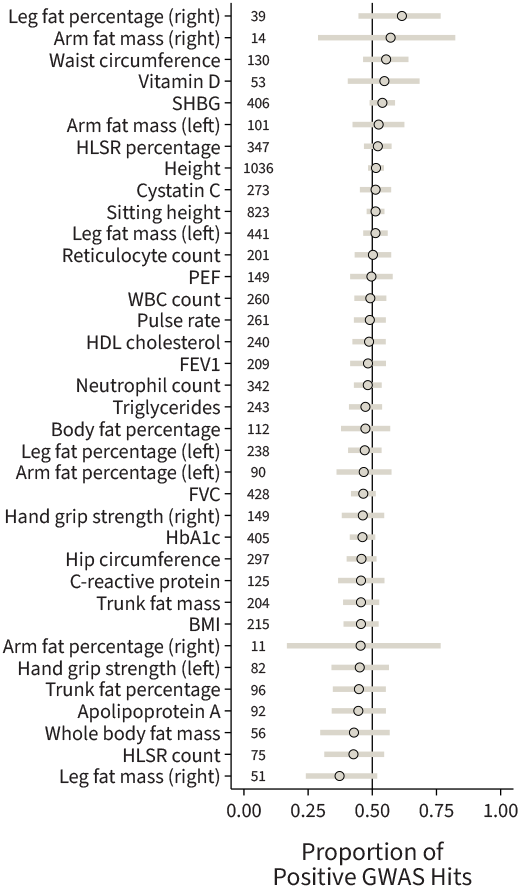
Proportion of trait-increasing derived alleles for each trait. This proportion is estimated from conditionally-independent GWAS hits, which are plotted in Figure I.1, with a derived allele frequency of less than 0.5. 95% confidence intervals were estimated using the Clopper-Pearson method. The number of variants in each test is included as a column on the left.

### I.3 Analysis of Monotonicity Effect Estimates

We estimated 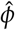 using both the MLE and the MoM estimates. The main results for the MLEs are presented in Figure 3B. The point estimates and confidence intervals for the MLEs are displayed in Figure I.3. The point estimates and confidence intervals for the MoM estimates are displayed in Figure I.4.

**Figure I.3.**
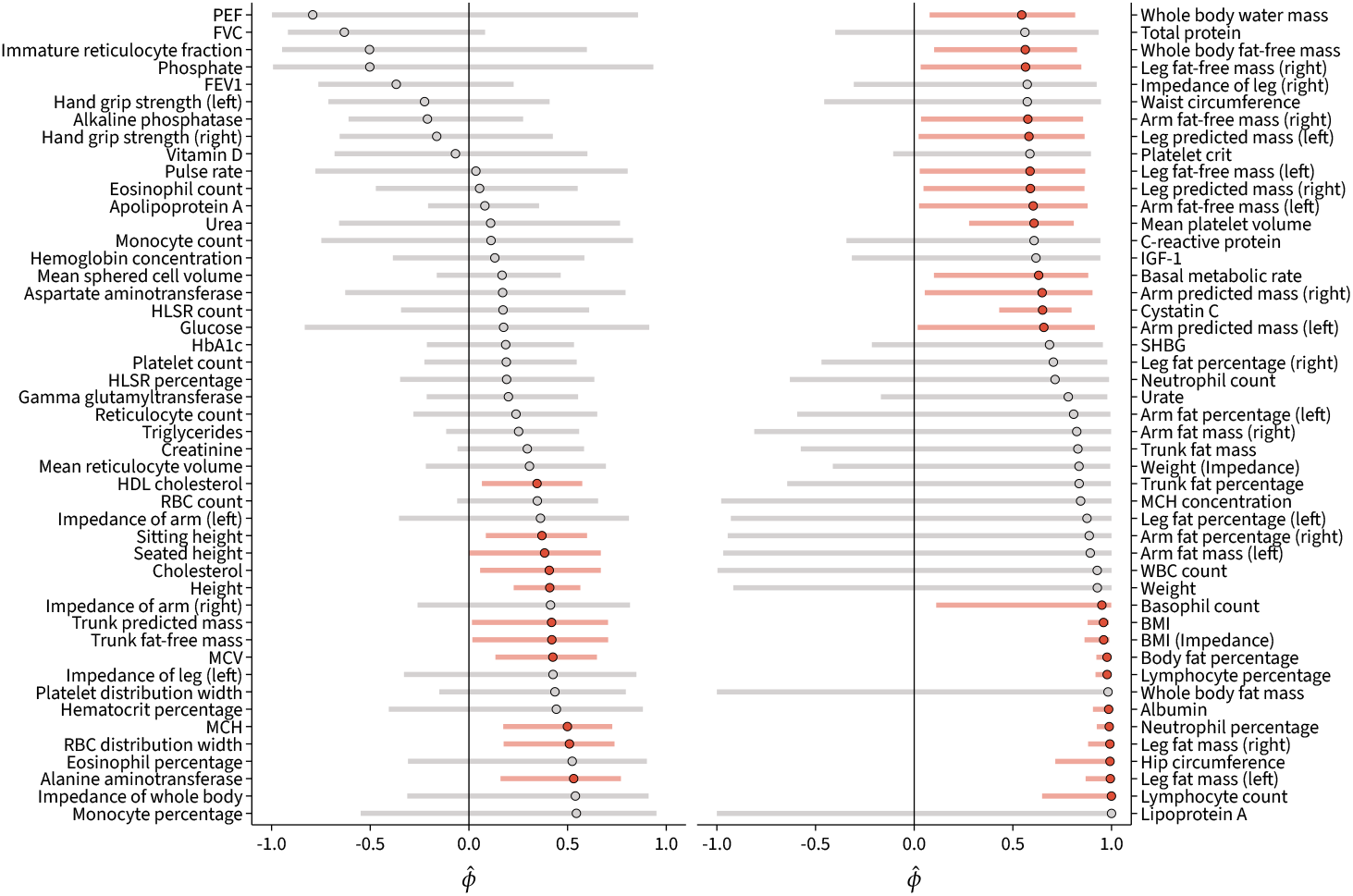
Point estimates with 95% confidence intervals for monotonicity from the maximum likelihood estimation approach. Orange points represent significantly positive values of 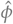 at the *α* = 0.05 level. Gray points represent non-significant values.

**Figure I.4.**
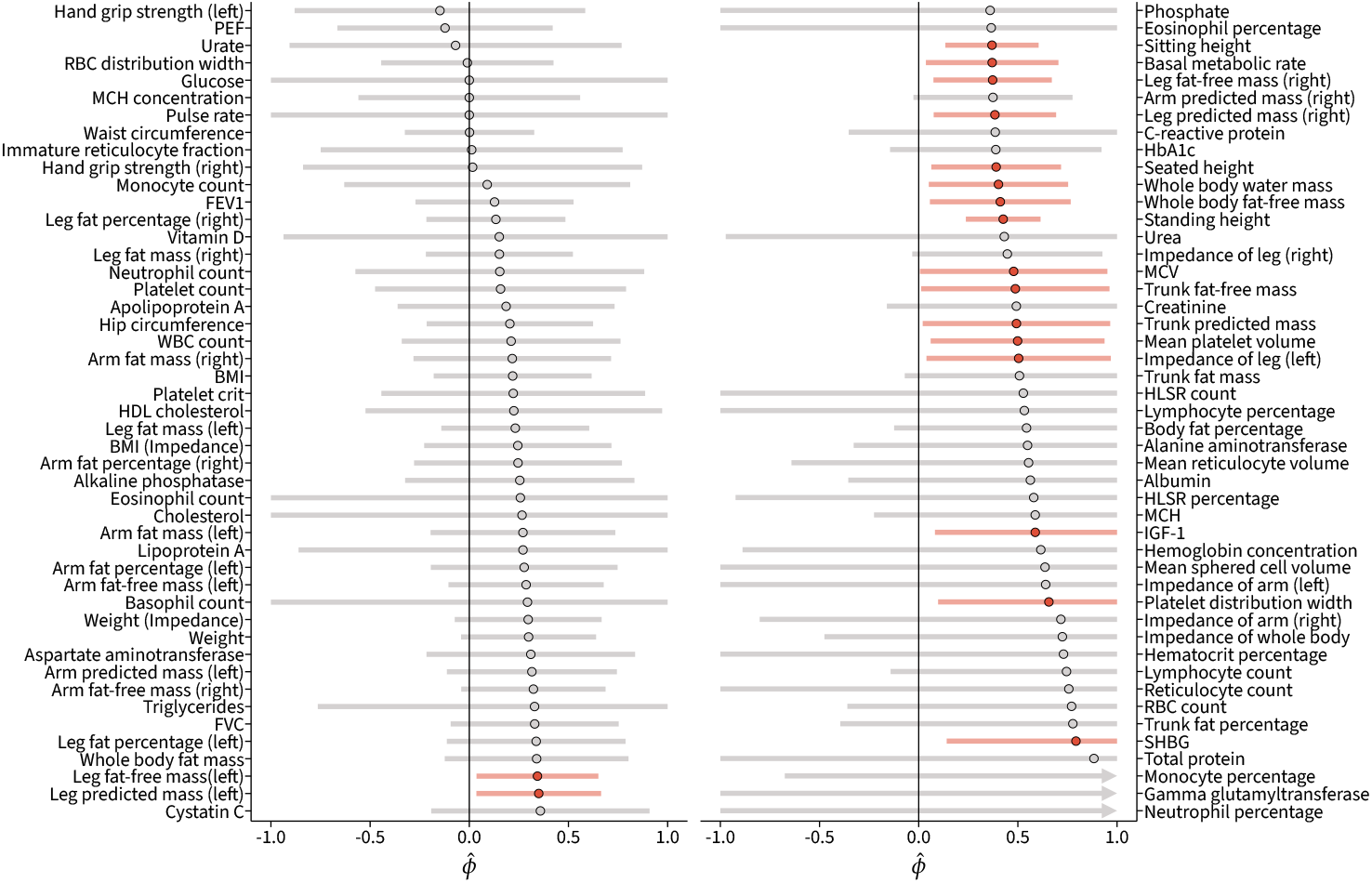
Point estimates with 95% confidence intervals for monotonicity from the method-of-moments (MoM) approach. Orange points represent significantly positive values of 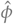 at the *α* = 0.05 level. Gray points represent non-significant values.

### I.4 Concordance of Monotonicity Estimates

Our two estimators for *ϕ* are based on different approaches (maximum likelihood estimation and MoM), and are each guaranteed to be consistent but not be unbiased in a finite sample. For this section, suppose that 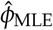 represents the MLE and 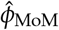 represents the MoM estimate. For the MLE, recall that we estimate the standard error for 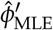 (Section C.1.2), which is a transformation of *ϕ* into an unconstrained space. For the MoM estimate, we use the bootstrap to estimate the standard error.

We use an errors-in-variables approach to estimate the concordance between the two estimators for *ϕ*. Suppose that

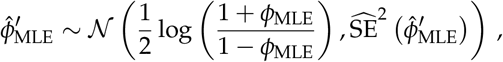

which is reasonable since the sampling distribution of the MLE asymptotically approaches this distribution. We make the stronger modeling assumption that

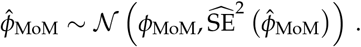

Finally, to estimate the concordance, we assume a linear relationship,

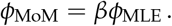

We fit this non-linear, errors-in-variables model using the orthogonal distance regression routines implemented in scipy [30]. The estimates from the two methods were broadly concordant, with a confidently positive estimate for the regression coefficient (Figure I.5).

**Figure I.5.**
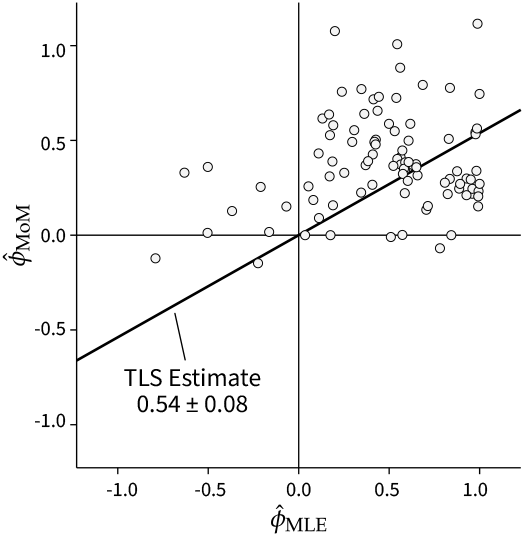
Comparing maximum likelihood estimation versus method-of-moments estimation for *ϕ*. The regression line is estimated using total least squares (TLS). A 95% confidence interval for the regression estimate is included. Each point represents one of the traits selected for analysis.

### I.5 Concordance of LoF and Deletion Effects

In addition to the monotone and non-monotone models that we present in Figure 1E and Figure 1F, the prior observation that genome-wide burden effects are non-zero for various traits may be due to a mechanism that is independent of dosage effects in *cis*. The effect of the number of CNVs in an individual may be due to dysregulation that is not caused by the genes present within the variant. If we assume that CNVs have such an independent effect, we would expect deletion burden effect estimates to be uncorrelated with LoF burden effect estimates.

To estimate the extent to which the burden tests using LoF variants are concordant with burden tests performed using deletions, we estimated the genetic correlation between the effects from the two variant classes. We did so by performing maximum likelihood estimation using the monotonicity model with an added constraint. Specifically, when we assume that the average effects are zero 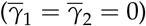, the correlation

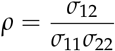

is similar to the genetic correlation derived under the LDSC model [31]. While we assume a normal prior distribution over the latent effect sizes, the original LDSC model assumes no functional prior form and only specifies finite first and second moments.

We derived the MLE 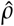 using the same inference machinery used for monotonicity, assuming fixed zero average effect sizes. We mapped *ρ ∈* [−1, 1] to an unconstrained space *ρ*^*′*^ *∈* ℝ by using

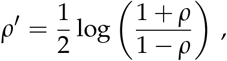

as we did for *ϕ* (Appendix C.1.2). The standard error for 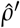 was estimated using the asymptotic approximation to the sampling distribution for maximum likelihood.

The estimated confidence intervals for *ρ* are displayed in Figure I.6. Since there are very few genes with deletions in the UKB (Appendix A.5), these estimates are much noisier than the estimates of monotonicity we present in Figure 3B.

**Figure I.6.**
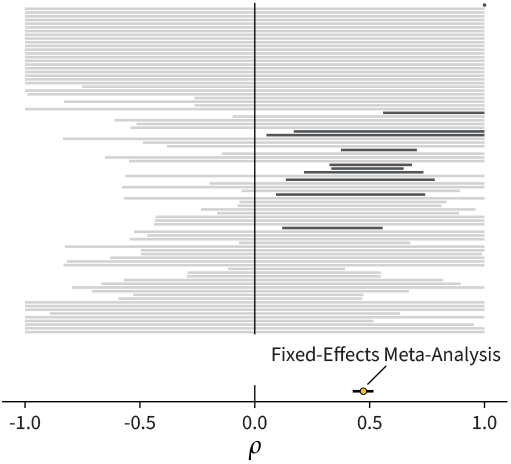
(Top) The bars represent 95% confidence intervals for *ρ* for the traits in our analysis. The black bars are significantly non-zero (p < 0.05). The bars are ordered by 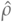. The point at the top represents a very confident estimate with a small confidence interval. (Bottom) The estimated *ρ* across traits using a fixed-effects meta-analysis.

We estimated a common *ρ* across all traits using a fixed-effects meta-analysis. Let 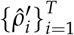 represent the observed, transformed genetic correlation estimates for *T* traits with standard errors 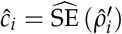. Suppose that

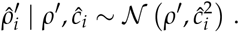

This is a reasonable choice because it is the asymptotic likelihood for maximum likelihood estimates. We performed a fixed-effects meta-analysis using the `metafor` package [32], and mapped back the estimate for *ρ*^*′*^ and the associated 95% confidence interval to *ρ* using the inverse map. The result is displayed in Figure I.6, and indicates a positive correlation between LoF and deletion burden effect estimates.

Next, we estimated the latent distribution of genetic correlations by fitting a transformed beta distribution. Let 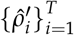 represent the observed, transformed genetic correlation estimates for *T* traits with standard errors 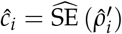. Suppose that

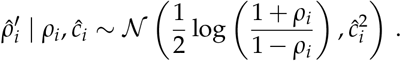

We assume that *ρ*_*i*_ is drawn from a transformed beta distribution over the interval [−1, 1],

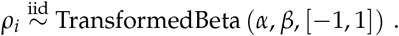

The likelihood for the transformed beta distribution can be derived using the transformation of random variables approach. Suppose that *U ∼* Beta (*α, β*). We define *V* as

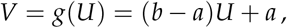

for *a, b ∈* ℝ and *a < b*. Note that the support of *V* is [*a, b*]. We then define

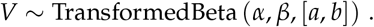

The inverse transform is

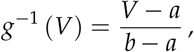

and the derivative is

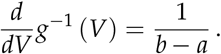

The likelihood is therefore

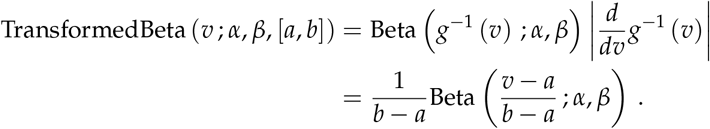

We use an empirical Bayes approach by fitting the prior distribution to the data. We do so by maximizing the evidence,

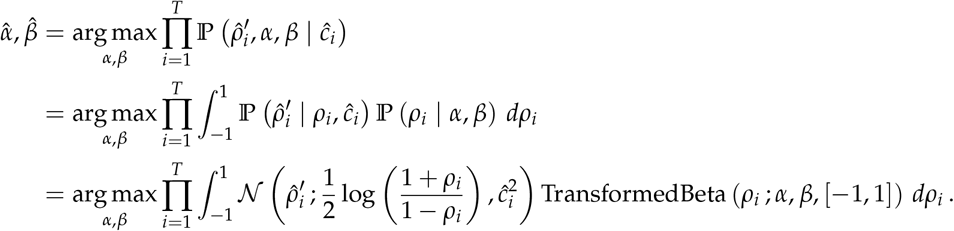

We used Riemann integration to numerically evaluate the evidence and used gradient ascent with a fixed number of steps to estimate 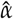 and 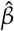. Gradients were estimated using automatic differentiation and optimization was performed using the `adamw` implementation in `optax` [10].

**Figure I.7.**
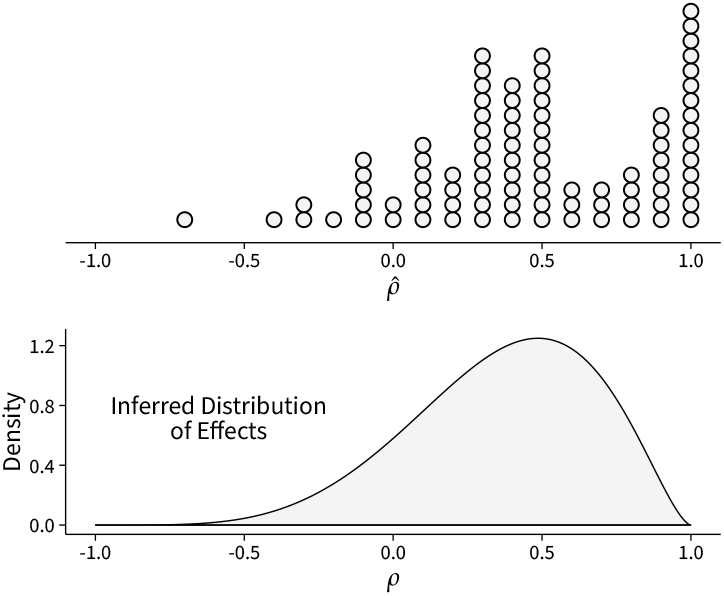
(Top) The maximum likelihood estimates (MLEs) for genetic correlation for the traits that were analyzed. The genetic correlation is estimated between loss-of-function (LoF) burden effects and deletion burden effects. (Bottom) The inferred distribution of effects from a random-effects meta-analysis.

This analysis demonstrates that LoF variant and deletion burden tests are markedly similar (Figure I.7). 87.0% of the density of the inferred distribution of genetic correlations is over the positive part of the domain of *ρ*. For this analysis, 5 of the 94 traits did not converge to estimates of *ρ*^*′*^, likely due to the small number of genes that have both LoF and deletion burden estimates, and were excluded from the analysis. These traits were: hand grip strength (left), arm fat mass (right), arm fat mass (left), urate, and whole body fat mass.

### I.6 Estimates of Trait Buffering Align with Average GDRC Direction

*ξ* is the average signed deviation of GDRCs from linearity, and should be concordant with the average GDRCs (aGDRCs) of the trait. For example, a trait experiencing negative trait buffering will have GDRCs that decrease a trait more than they increase the trait (Figure 5A). The aGDRC for these curves should be in the negative direction, which implies that the average LoF burden effect and the average duplication burden effect are negative. Indeed, we observe that our estimates of *ξ* align well with the aGDRCs of the traits (Figure I.8).

**Figure I.8.**
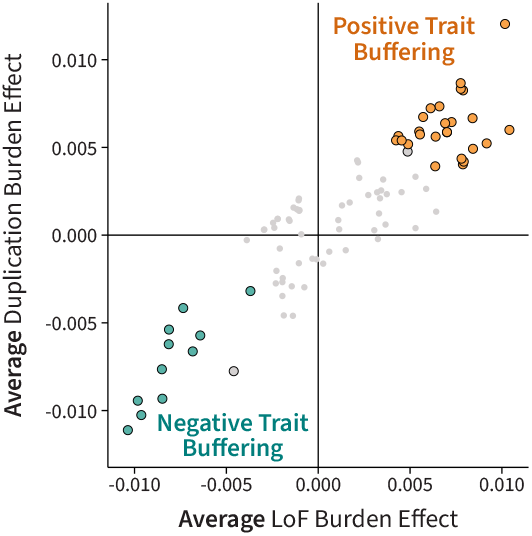
The average LoF and duplication burden effects for the traits in our analysis. The data presented here is the same as Figure 1B. Large points represents traits with a nominally significant average LoF burden effect and average duplication burden effect at the *α* = 0.05 level. The color of the points represents the sign of the estimate of *ξ* for all genes with a local false sign rate of less than 5%.

### I.7 Posterior Mean Burden Effect Estimates for Traits

We can use the posterior estimates from our trait buffering model to visualize these latent spaces for traits that we study. In our main figure (Figure 5C), we looked specifically at BMI, height, and peak expiratory flow (PEF). We can explore these using the posterior samples from our trait buffering model. The posterior mean effect sizes from our model for BMI, height, and PEF are displayed in Figures I.9, I.10, and I.11 respectively. In each case, we can see the effects of trait buffering by visualizing the distance genes deviate from the diagonal line.

**Figure I.9.**
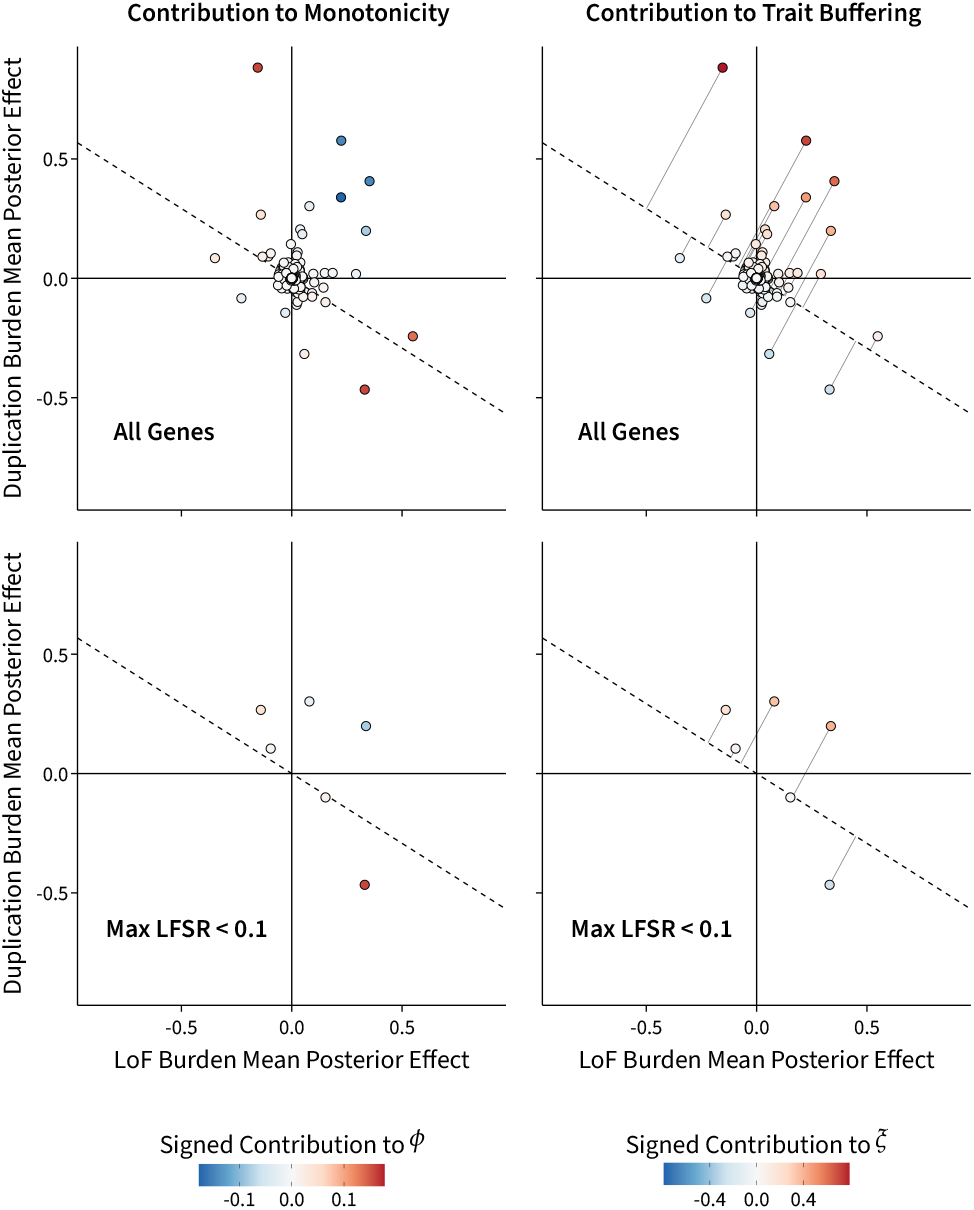
Posterior mean effect estimates for LoF and duplication burden tests from our trait buffering model for body mass index (BMI). The top row displays all genes, while the bottom row displays genes where both LoF and duplication burden tests have a local false sign rate (LFSR) of less than 10%. The left column displays the contribution of each gene to *ϕ*, while the right column displays the contribution of each gene to *ξ*.

**Figure I.10.**
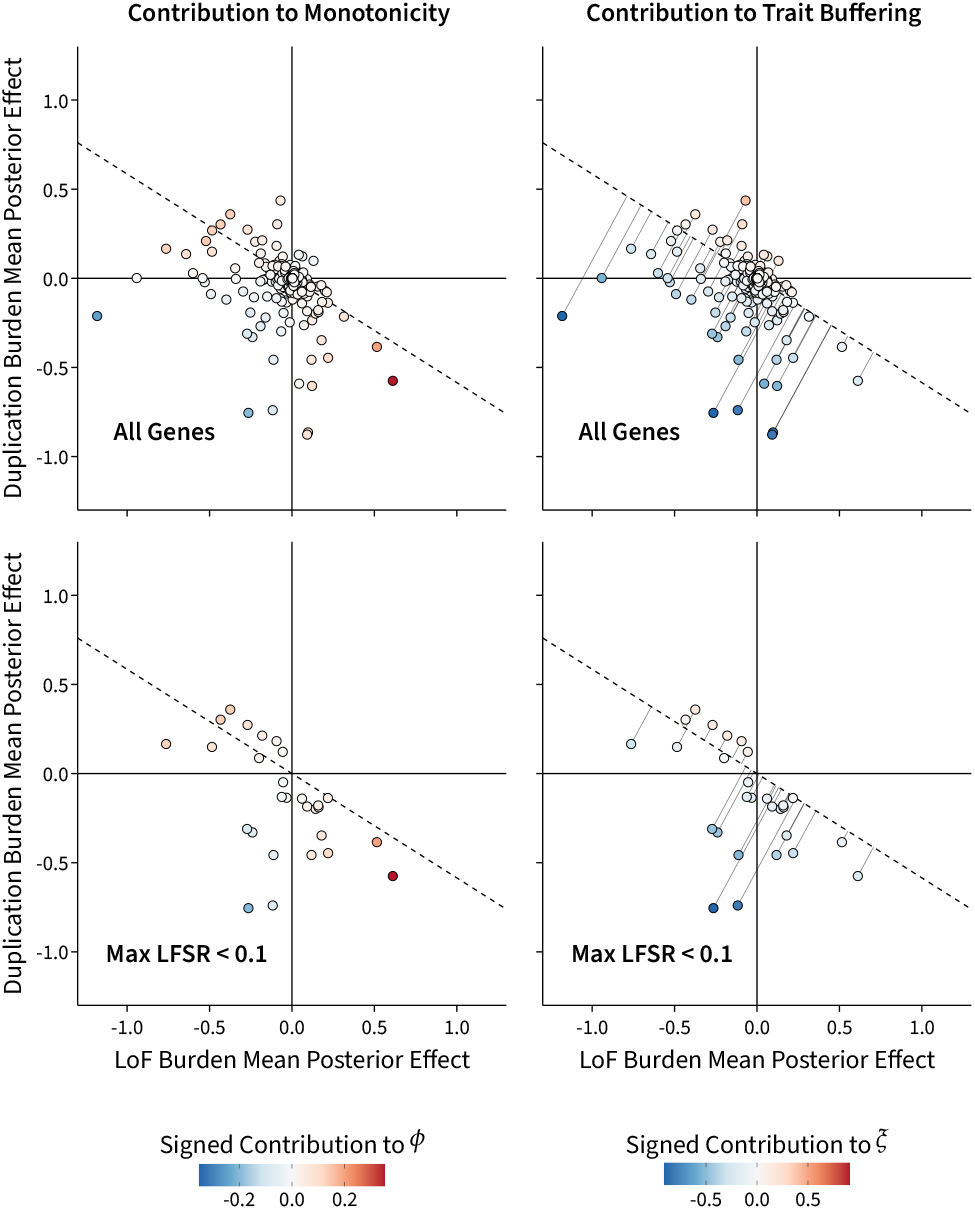
Posterior mean effect estimates for LoF and duplication burden tests from our trait buffering model for height. The top row displays all genes, while the bottom row displays genes where both LoF and duplication burden tests have a local false sign rate (LFSR) of less than 10%. The left column displays the contribution of each gene to *ϕ*, while the right column displays the contribution of each gene to *ξ*.

**Figure I.11.**
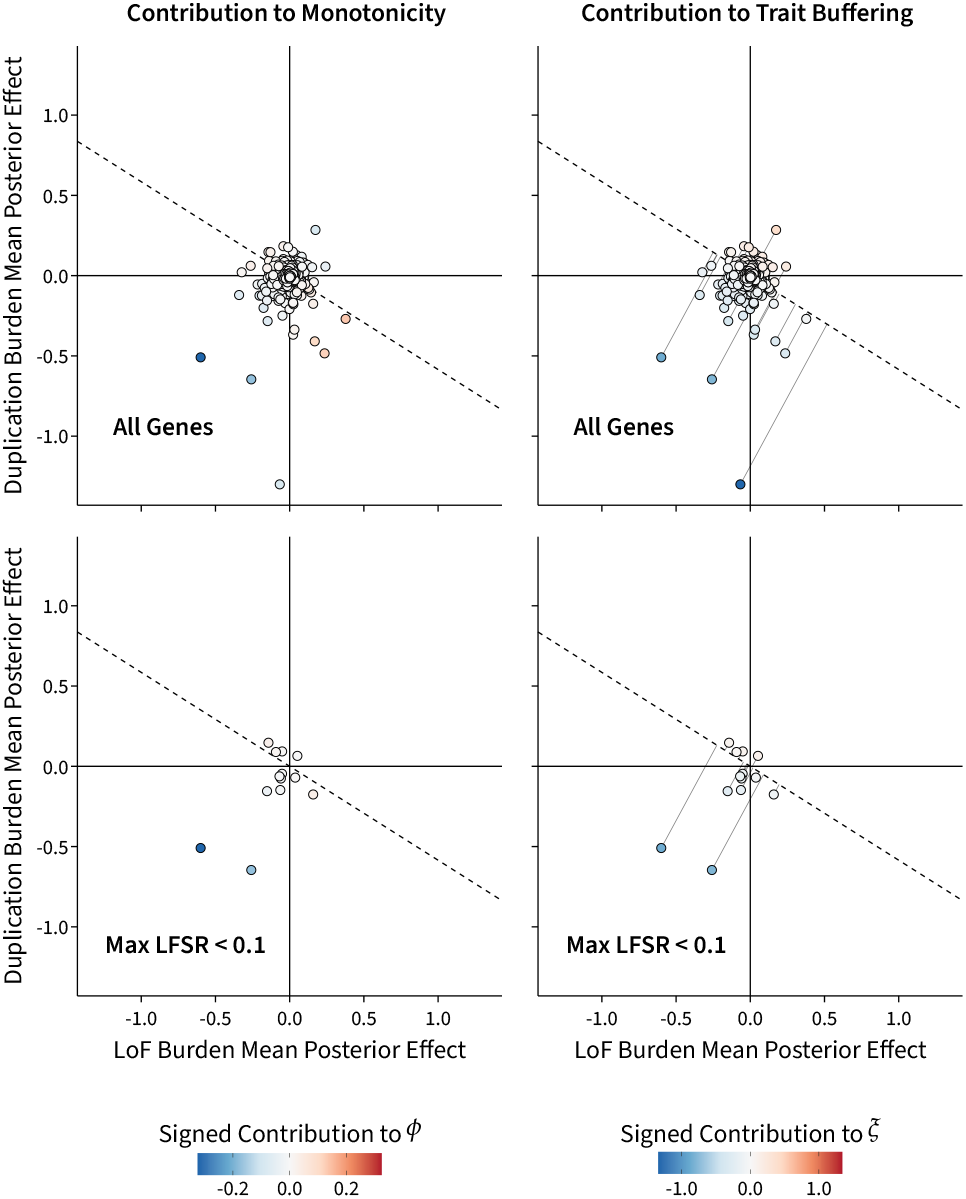
Posterior mean effect estimates for LoF and duplication burden tests from our trait buffering model for peak expiratory flow (PEF). The top row displays all genes, while the bottom row displays genes where both LoF and duplication burden tests have a local false sign rate (LFSR) of less than 10%. The left column displays the contribution of each gene to *ϕ*, while the right column displays the contribution of each gene to *ξ*.

The contribution to *ϕ* that is used in the visualization of Figures I.9, I.10, and I.11 is derived from the posterior samples for each gene. It represents the average value of −*γ*_LoF_ *× γ*_Dup_ for each gene across the posterior samples, and is not strictly equivalent to the contribution to *ϕ* under the monotonicity model. In contrast, the contribution to *ξ* used in the visualization can be interpreted as the gene-level contribution to the statistic reported in our main analysis since it is based on the posterior samples.

BMI is a trait with large and positive 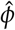, height is a trait with moderate and positive 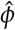, and PEF is a trait with non-significant 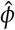. These visualized latent spaces should thus span the various types of traits present in our data set. BMI has a positive average GDRC, while both height and PEF have negative average GDRCs. As expected, the direction of trait buffering for these traits is consistent with their average GDRCs and is visually discernible in the visualized latent spaces.

## References

[1] Melina Claussnitzer et al. “A brief history of human disease genetics”. Nature 577.7789 (Jan. 2020), pp. 179–189. DOI: 10.1038/s41586-019-1879-7.

[2] Elliot Sollis et al. “The NHGRI-EBI GWAS Catalog: knowledgebase and deposition resource”. Nucleic Acids Research 51.D1 (Jan. 2023), pp. D977–D985. DOI: 10.1093/nar/gkac1010.

[3] Matthew T. Maurano et al. “Systematic Localization of Common Disease-Associated Variation in Regulatory DNA”. Science 337.6099 (Sept. 2012), pp. 1190–1195. DOI: 10.1126/science.1222794.

[4] Anshul Kundaje et al. “Integrative analysis of 111 reference human epigenomes”. Nature 518.7539 (Feb. 2015), pp. 317–330. DOI: 10.1038/nature14248.

[5] Eric R. Gamazon et al. “A gene-based association method for mapping traits using reference transcriptome data”. Nature Genetics 47.9 (Aug. 2015), pp. 1091–1098. DOI: 10.1038/ng.3367.

[6] Alexander Gusev et al. “Integrative approaches for large-scale transcriptome-wide association studies”. Nature Genetics 48.3 (Feb. 2016), pp. 245–252. DOI: 10.1038/ng.3506.

[7] Farhad Hormozdiari et al. “Colocalization of GWAS and eQTL Signals Detects Target Genes”. The American Journal of Human Genetics 99.6 (Dec. 2016), pp. 1245–1260. DOI: 10.1016/j.ajhg.2016.10.003.

[8] Wouter Meuleman et al. “Index and biological spectrum of human DNase I hypersensitive sites”. Nature 584.7820 (July 2020), pp. 244–251. DOI: 10.1038/s41586-020-2559-3.

[9] Silvia Stingele et al. “Global analysis of genome, transcriptome and proteome reveals the response to aneuploidy in human cells”. Molecular Systems Biology 8.1 (Sept. 2012), p. 608. DOI: 10.1038/msb.2012.40.

[10] Terry Hassold and Patricia Hunt. “To err (meiotically) is human: the genesis of human aneuploidy”. Nature Reviews Genetics 2.4 (Apr. 2001), pp. 280–291. DOI: 10.1038/35066065.

[11] CD Lehman et al. “Trisomy 13 syndrome: prenatal US findings in a review of 33 cases.” Radiology 194.1 (Jan. 1995), pp. 217–222. DOI: 10.1148/radiology.194.1.7997556.

[12] Anna Cereda and John C. Carey. “The trisomy 18 syndrome”. Orphanet Journal of Rare Diseases 7.1 (Oct. 2012), p. 81. DOI: 10.1186/1750-1172-7-81.

[13] Marilyn J. Bull. “Down Syndrome”. New England Journal of Medicine 382.24 (June 2020), pp. 2344– 2352. DOI: 10.1056/NEJMra1706537.

[14] Xin Shao et al. “Copy number variation is highly correlated with differential gene expression: a pan-cancer study”. BMC medical genetics 20.1 (Nov. 2019), p. 175. DOI: 10.1186/s12881-019-0909-5.

[15] Peter H. Sudmant et al. “Global diversity, population stratification, and selection of human copynumber variation”. Science 349.6253 (Sept. 2015), aab3761. DOI: 10.1126/science.aab3761.

[16] Haley J. Abel et al. “Mapping and characterization of structural variation in 17,795 human genomes”. Nature 583.7814 (May 2020), pp. 83–89. DOI: 10.1038/s41586-020-2371-0.

[17] Ryan L. Collins et al. “A structural variation reference for medical and population genetics”. Nature 581.7809 (May 2020), pp. 444–451. DOI: 10.1038/s41586-020-2287-8.

[18] Mohamed A. Almarri et al. “Population Structure, Stratification, and Introgression of Human Structural Variation”. Cell 182.1 (July 2020), 189–199.e15. DOI: 10.1016/j.cell.2020.05.024.

[19] Andrew Dauber et al. “Genome-wide Association of Copy-Number Variation Reveals an Association between Short Stature and the Presence of Low-Frequency Genomic Deletions”. The American Journal of Human Genetics 89.6 (Dec. 2011), pp. 751–759. DOI: 10.1016/j.ajhg.2011.10.014.

[20] Eleanor Wheeler et al. “Genome-wide SNP and CNV analysis identifies common and low-frequency variants associated with severe early-onset obesity”. Nature Genetics 45.5 (Apr. 2013), pp. 513–517. DOI: 10.1038/ng.2607.

[21] Aurélien Macé et al. “CNV-association meta-analysis in 191,161 European adults reveals new loci associated with anthropometric traits”. Nature Communications 8.1 (Sept. 2017), p. 744. DOI: 10.1038/s41467-017-00556-x.

[22] Christian R. Marshall et al. “Contribution of copy number variants to schizophrenia from a genomewide study of 41,321 subjects”. Nature Genetics 49.1 (Nov. 2016), pp. 27–35. DOI: 10.1038/ng.3725.

[23] Matthew Aguirre, Manuel A. Rivas, and James Priest. “Phenome-wide Burden of Copy-Number Variation in the UK Biobank”. The American Journal of Human Genetics 105.2 (Aug. 2019), pp. 373– 383. DOI: 10.1016/j.ajhg.2019.07.001.

[24] Chiara Auwerx et al. “The individual and global impact of copy-number variants on complex human traits”. The American Journal of Human Genetics 109.4 (Apr. 2022), pp. 647–668. DOI: 10.1016/j.ajhg.2022.02.010.

[25] Margaux L. A. Hujoel et al. “Influences of rare copy-number variation on human complex traits”. Cell 185.22 (Oct. 2022), 4233–4248.e27. DOI: 10.1016/j.cell.2022.09.028.

[26] Ryan L. Collins et al. “A cross-disorder dosage sensitivity map of the human genome”. Cell 185.16 (Aug. 2022), 3041–3055.e25. DOI: 10.1016/j.cell.2022.06.036.

[27] Chiara Auwerx et al. Copy-number variants as modulators of common disease susceptibility. Aug. 2023. DOI: 10.1101/2023.07.31.23293408.

[28] Andrea Ganna et al. “Quantifying the Impact of Rare and Ultra-rare Coding Variation across the Phenotypic Spectrum”. The American Journal of Human Genetics 102.6 (June 2018), pp. 1204–1211. DOI: 10.1016/j.ajhg.2018.05.002.

[29] Chia-Yen Chen et al. “The impact of rare protein coding genetic variation on adult cognitive function”. Nature Genetics 55.6 (June 2023), pp. 927–938. DOI: 10.1038/s41588-023-01398-8.

[30] Yunfeng Huang et al. “Rare genetic variants impact muscle strength”. Nature Communications 14.1 (June 2023), p. 3449. DOI: 10.1038/s41467-023-39247-1.

[31] Joshua D. Backman et al. “Exome sequencing and analysis of 454,787 UK Biobank participants”. Nature 599.7886 (Oct. 2021), pp. 628–634. DOI: 10.1038/s41586-021-04103-z.

[32] Shuwei Li et al. Whole-genome sequencing of half-a-million UK Biobank participants. Dec. 2023. DOI: 10.1101/2023.12.06.23299426.

[33] Joelle Mbatchou et al. “Computationally efficient whole-genome regression for quantitative and binary traits”. Nature Genetics 53.7 (May 2021), pp. 1097–1103. DOI: 10.1038/s41588-021-00870-7.

[34] Michael S. Brown and Joseph L. Goldstein. “Lowering LDL–Not Only How Low, But How Long?” Science 311.5768 (Mar. 2006), pp. 1721–1723. DOI: 10.1126/science.1125884.

[35] Joseph L. Goldstein and Michael S. Brown. “The LDL Receptor”. Arteriosclerosis, Thrombosis, and Vascular Biology 29.4 (Apr. 2009), pp. 431–438. DOI: 10.1161/ATVBAHA.108.179564.

[36] Tsutomu Ogata, Nobutake Matsuo, and Gen Nishimura. “SHOX haploinsufficiency and overdosage: impact of gonadal function status”. Journal of Medical Genetics 38.1 (Jan. 2001), pp. 1–6. DOI: 10.1136/jmg.38.1.1.

[37] Jeremy J. Berg et al. Mutation-selection-drift balance models of complex diseases. May 2025. DOI: 10.1101/2025.05.18.654722.

[38] Robert M. Plenge, Edward M. Scolnick, and David Altshuler. “Validating therapeutic targets through human genetics”. Nature Reviews Drug Discovery 12.8 (July 2013), pp. 581–594. DOI: 10.1038/nrd4051.

[39] Leeat Keren et al. “Massively Parallel Interrogation of the Effects of Gene Expression Levels on Fitness”. Cell 166.5 (Aug. 2016), 1282–1294.e18. DOI: 10.1016/j.cell.2016.07.024.

[40] Sahin Naqvi et al. “Precise modulation of transcription factor levels identifies features underlying dosage sensitivity”. Nature Genetics 55.5 (May 2023), pp. 841–851. DOI: 10.1038/s41588-023-01366-2.

[41] Júlia Domingo et al. “Non-linear transcriptional responses to gradual modulation of transcription factor dosage”. eLife 13 (Dec. 2024). DOI: 10.7554/eLife.100555.1.

[42] Sébastien Jacquemont et al. “Mirror extreme BMI phenotypes associated with gene dosage at the chromosome 16p11.2 locus”. Nature 478.7367 (Aug. 2011), pp. 97–102. DOI: 10.1038/nature10406.

[43] Christelle Golzio et al. “KCTD13 is a major driver of mirrored neuroanatomical phenotypes of the 16p11.2 copy number variant”. Nature 485.7398 (May 2012), pp. 363–367. DOI: 10.1038/nature11091.

[44] John D. Storey. “A Direct Approach to False Discovery Rates”. Journal of the Royal Statistical Society Series B: Statistical Methodology 64.3 (Aug. 2002), pp. 479–498. DOI: 10.1111/1467-9868.00346.

[45] Lauren A. Weiss et al. “Association between Microdeletion and Microduplication at 16p11.2 and Autism”. New England Journal of Medicine 358.7 (Feb. 2008), pp. 667–675. DOI: 10.1056/NEJMoa075974.

[46] Christelle Golzio and Nicholas Katsanis. “Genetic architecture of reciprocal CNVs”. Current Opinion in Genetics & Development. Molecular and genetic bases of disease 23.3 (June 2013), pp. 240–248. DOI: 10.1016/j.gde.2013.04.013.

[47] Chiara Auwerx et al. “Rare copy-number variants as modulators of common disease susceptibility”. Genome Medicine 16.1 (Jan. 2024), p. 5. DOI: 10.1186/s13073-023-01265-5.

[48] Chiara Auwerx et al. “Disentangling mechanisms behind the pleiotropic effects of proximal 16p11.2 BP4-5 CNVs”. The American Journal of Human Genetics 0.0 (Sept. 2024). DOI: 10.1016/j.ajhg.2024.08.014.

[49] Veronique Vitart et al. “SLC2A9 is a newly identified urate transporter influencing serum urate concentration, urate excretion and gout”. Nature Genetics 40.4 (Apr. 2008), pp. 437–442. DOI: 10.1038/ng.106.

[50] Naohiko Anzai et al. “Plasma urate level is directly regulated by a voltage-driven urate efflux transporter URATv1 (SLC2A9) in humans”. The Journal of Biological Chemistry 283.40 (Oct. 2008), pp. 26834–26838. DOI: 10.1074/jbc.C800156200.

[51] David B. Mount, Charles Y. Kwon, and Kambiz Zandi-Nejad. “Renal Urate Transport”. Rheumatic Disease Clinics of North America. Gout 32.2 (May 2006), pp. 313–331. DOI: 10.1016/j.rdc.2006.02.006.

[52] A. Kugelman et al. “gamma-Glutamyl transpeptidase is increased by oxidative stress in rat alveolar L2 epithelial cells”. American Journal of Respiratory Cell and Molecular Biology 11.5 (Nov. 1994), pp. 586– 592. DOI: 10.1165/ajrcmb.11.5.7946387.

[53] Duk-Hee Lee, Rune Blomhoff, and David R. Jacobs. “Is serum gamma glutamyltransferase a marker of oxidative stress?” Free Radical Research 38.6 (June 2004), pp. 535–539. DOI: 10.1080/10715760410001694026.

[54] Cui Bai et al. “Gamma-Glutamyltransferase Activity (GGT) Is a Long-Sought Biomarker of Redox Status in Blood Circulation: A Retrospective Clinical Study of 44 Types of Human Diseases”. Oxidative Medicine and Cellular Longevity 2022 (June 2022), p. 8494076. DOI: 10.1155/2022/8494076.

[55] André B. P. van Kuilenburg et al. “beta-Ureidopropionase deficiency: an inborn error of pyrimidine degradation associated with neurological abnormalities”. Human Molecular Genetics 13.22 (Nov. 2004), pp. 2793–2801. DOI: 10.1093/hmg/ddh303.

[56] Aza Shetewy et al. “Mitochondrial defects associated with -alanine toxicity: relevance to hyper-betaalaninemia”. Molecular and cellular biochemistry 416.1-2 (May 2016), pp. 11–22. DOI: 10.1007/s11010-016-2688-z.

[57] Doreen Dobritzsch et al. “-Ureidopropionase deficiency due to novel and rare UPB1 mutations affecting pre-mRNA splicing and protein structural integrity and catalytic activity”. Molecular Genetics and Metabolism 136.3 (July 2022), pp. 177–185. DOI: 10.1016/j.ymgme.2022.01.102.

[58] Sarah M. Urbut et al. “Flexible statistical methods for estimating and testing effects in genomic studies with multiple conditions”. Nature Genetics 51.1 (Nov. 2018), pp. 187–195. DOI: 10.1038/s41588-018-0268-8.

[59] Yuval B. Simons et al. “A population genetic interpretation of GWAS findings for human quantitative traits”. PLOS Biology 16.3 (Mar. 2018), e2002985. DOI: 10.1371/journal.pbio.2002985.

[60] Guy Sella and Nicholas H. Barton. “Thinking About the Evolution of Complex Traits in the Era of Genome-Wide Association Studies”. Annual Review of Genomics and Human Genetics 20.1 (Aug. 2019), pp. 461–493. DOI: 10.1146/annurev-genom-083115-022316.

[61] Yuval B. Simons et al. Simple scaling laws control the genetic architectures of human complex traits. Oct. 2022. DOI: 10.1101/2022.10.04.509926.

[62] E. Koch et al. Genetic association data are broadly consistent with stabilizing selection shaping human common diseases and traits. July 2024. DOI: 10.1101/2024.06.19.599789.

[63] Roshni A. Patel et al. Conditional frequency spectra as a tool for studying selection on complex traits in biobanks. June 2024. DOI: 10.1101/2024.06.15.599126.

[64] Mei Sum Chan et al. “A Biomarker-based Biological Age in UK Biobank: Composition and Prediction of Mortality and Hospital Admissions”. The Journals of Gerontology: Series A 76.7 (July 2021), pp. 1295–1302. DOI: 10.1093/gerona/glab069.

[65] Sergiy Libert, Alex Chekholko, and Cynthia Kenyon. A mathematical model that predicts human biological age from physiological traits identifies environmental and genetic factors that influence aging. Oct. 2023. DOI: 10.1101/2022.04.14.488358.

[66] Carlos López-Otín et al. “The Hallmarks of Aging”. Cell 153.6 (June 2013), pp. 1194–1217. DOI: 10.1016/j.cell.2013.05.039.

[67] Ralf Schmidt et al. “CRISPR activation and interference screens decode stimulation responses in primary human T cells”. Science 375.6580 (Feb. 2022), eabj4008. DOI: 10.1126/science.abj4008.

[68] Anna Hutchinson, Jennifer Asimit, and Chris Wallace. “Fine-mapping genetic associations”. Human Molecular Genetics 29.R1 (Sept. 2020), R81–R88. DOI: 10.1093/hmg/ddaa148.

[69] Tuuli Lappalainen and Daniel G. MacArthur. “From variant to function in human disease genetics”. Science 373.6562 (Sept. 2021), pp. 1464–1468. DOI: 10.1126/science.abi8207.

[70] Joseph Nasser et al. “Genome-wide enhancer maps link risk variants to disease genes”. Nature 593.7858 (Apr. 2021), pp. 238–243. DOI: 10.1038/s41586-021-03446-x.

[71] Noah J Connally et al. “The missing link between genetic association and regulatory function”. eLife 11 (Dec. 2022). Ed. by Jonathan Flint and Molly Przeworski, e74970. DOI: 10.7554/eLife.74970.

[72] Steven Gazal et al. “Combining SNP-to-gene linking strategies to identify disease genes and assess disease omnigenicity”. Nature Genetics 54.6 (June 2022), pp. 827–836. DOI: 10.1038/s41588-022-01087-y.

[73] Andreas R. Gschwind et al. An encyclopedia of enhancer-gene regulatory interactions in the human genome. Nov. 2023. DOI: 10.1101/2023.11.09.563812.

[74] Nicholas Mancuso et al. “Probabilistic fine-mapping of transcriptome-wide association studies”. Nature Genetics 51.4 (Mar. 2019), pp. 675–682. DOI: 10.1038/s41588-019-0367-1.

[75] Zhihong Zhu et al. “Integration of summary data from GWAS and eQTL studies predicts complex trait gene targets”. Nature Genetics 48.5 (Mar. 2016), pp. 481–487. DOI: 10.1038/ng.3538.

[76] Hakhamanesh Mostafavi et al. “Systematic differences in discovery of genetic effects on gene expression and complex traits”. Nature Genetics (Oct. 2023), pp. 1–10. DOI: 10.1038/s41588-023-01529-1.

[77] Tony Zeng et al. “Bayesian estimation of gene constraint from an evolutionary model with gene features”. Nature Genetics (July 2024), pp. 1–12. DOI: 10.1038/s41588-024-01820-9.

[78] David R. Kelley et al. “Sequential regulatory activity prediction across chromosomes with convolutional neural networks”. Genome Research 28.5 (Mar. 2018), pp. 739–750. DOI: 10.1101/gr.227819.117.

[79] Vikram Agarwal and Jay Shendure. “Predicting mRNA Abundance Directly from Genomic Sequence Using Deep Convolutional Neural Networks”. Cell Reports 31.7 (May 2020). DOI: 10.1016/j.celrep.2020.107663.

[80] iga Avsec et al. “Effective gene expression prediction from sequence by integrating long-range interactions”. Nature Methods 18.10 (Oct. 2021), pp. 1196–1203. DOI: 10.1038/s41592-021-01252-x.

[81] Shiron Drusinsky, Sean Whalen, and Katherine S. Pollard. Deep-learning prediction of gene expression from personal genomes. July 2024. DOI: 10.1101/2024.07.27.605449.

[82] Atray Dixit et al. “Perturb-Seq: Dissecting Molecular Circuits with Scalable Single-Cell RNA Profiling of Pooled Genetic Screens”. Cell 167.7 (Dec. 2016), 1853–1866.e17. DOI: 10.1016/j.cell.2016.11.038.

[83] Ryan Tewhey et al. “Direct Identification of Hundreds of Expression-Modulating Variants using a Multiplexed Reporter Assay”. Cell 165.6 (June 2016), pp. 1519–1529. DOI: 10.1016/j.cell.2016.04.027.

[84] Martin Kircher et al. “Saturation mutagenesis of twenty disease-associated regulatory elements at single base-pair resolution”. Nature Communications 10.1 (Aug. 2019), p. 3583. DOI: 10.1038/s41467-019-11526-w.

[85] Jun Cheng et al. “Accurate proteome-wide missense variant effect prediction with AlphaMissense”. Science 381.6664 (Sept. 2023), eadg7492. DOI: 10.1126/science.adg7492.

[86] Douglas M. Fowler and Stanley Fields. “Deep mutational scanning: a new style of protein science”. Nature Methods 11.8 (July 2014), pp. 801–807. DOI: 10.1038/nmeth.3027.

[87] GTEx Consortium. “Genetic effects on gene expression across human tissues”. Nature 550.7675 (Oct. 2017), pp. 204–213. DOI: 10.1038/nature24277.

[88] Marco Jost et al. “Titrating gene expression using libraries of systematically attenuated CRISPR guide RNAs”. Nature Biotechnology 38.3 (Mar. 2020), pp. 355–364. DOI: 10.1038/s41587-019-0387-5.

[89] Julia Joung et al. “A transcription factor atlas of directed differentiation”. Cell 186.1 (Jan. 2023), 209– 229.e26. DOI: 10.1016/j.cell.2022.11.026.

[90] Srinivas Niranj Chandrasekaran et al. “Three million images and morphological profiles of cells treated with matched chemical and genetic perturbations”. Nature Methods 21.6 (June 2024), pp. 1114– 1121. DOI: 10.1038/s41592-024-02241-6.

[91] Bicna Song et al. “Decoding heterogeneous single-cell perturbation responses”. Nature Cell Biology 27.3 (Mar. 2025), pp. 493–504. DOI: 10.1038/s41556-025-01626-9.

[92] Min Zhao et al. “Computational tools for copy number variation (CNV) detection using next-generation sequencing data: features and perspectives”. BMC Bioinformatics 14.11 (Sept. 2013), S1. DOI: 10.1186/1471-2105-14-S11-S1.

[93] Xiang Zhu and Matthew Stephens. “Bayesian large-scale multiple regression with summary statistics from genome-wide association studies”. The Annals of Applied Statistics 11.3 (Sept. 2017), pp. 1561– 1592. DOI: 10.1214/17-AOAS1046.

[94] Heli Julkunen et al. “Atlas of plasma NMR biomarkers for health and disease in 118,461 individuals from the UK Biobank”. Nature Communications 14.1 (Feb. 2023), p. 604. DOI: 10.1038/s41467-023-36231-7.

[95] Clare Bycroft et al. “The UK Biobank resource with deep phenotyping and genomic data”. Nature 562.7726 (Oct. 2018), pp. 203–209. DOI: 10.1038/s41586-018-0579-z.

[96] Gad Abraham, Yixuan Qiu, and Michael Inouye. “FlashPCA2: principal component analysis of Biobank-scale genotype datasets”. Bioinformatics 33.17 (Sept. 2017), pp. 2776–2778. DOI: 10.1093/bioinformatics/btx299.

[97] William McLaren et al. “The Ensembl Variant Effect Predictor”. Genome Biology 17.1 (June 2016), p. 122. DOI: 10.1186/s13059-016-0974-4.

[98] Konrad J. Karczewski et al. “The mutational constraint spectrum quantified from variation in 141,456 humans”. Nature 581.7809 (May 2020), pp. 434–443. DOI: 10.1038/s41586-020-2308-7.

[99] Sairam Behera et al. “Comprehensive genome analysis and variant detection at scale using DRA-GEN”. Nature Biotechnology (Oct. 2024), pp. 1–15. DOI: 10.1038/s41587-024-02382-1.

[100] The UK Biobank Whole-Genome Sequencing Consortium et al. “Whole-genome sequencing of 490,640 UK Biobank participants”. Nature (Aug. 2025), pp. 1–10. DOI: 10.1038/s41586-025-09272-9.

[101] Adam Frankish et al. “GENCODE: reference annotation for the human and mouse genomes in 2023”. Nucleic Acids Research 51.D1 (Jan. 2023), pp. D942–D949. DOI: 10.1093/nar/gkac1071.

[102] Karen N. Conneely and Michael Boehnke. “So Many Correlated Tests, So Little Time! Rapid Adjustment of P Values for Multiple Correlated Tests”. The American Journal of Human Genetics 81.6 (Dec. 2007), pp. 1158–1168. DOI: 10.1086/522036.

[103] Brendan K. Bulik-Sullivan et al. “LD Score regression distinguishes confounding from polygenicity in genome-wide association studies”. Nature Genetics 47.3 (Feb. 2015), pp. 291–295. DOI: 10.1038/ng.3211.

[104] Bernard Delyon, Marc Lavielle, and Eric Moulines. “Convergence of a Stochastic Approximation Version of the EM Algorithm”. The Annals of Statistics 27.1 (Feb. 1999), pp. 94–128.

[105] Estelle Kuhn and Marc Lavielle. “Coupling a stochastic approximation version of EM with an MCMC procedure”. ESAIM: Probability and Statistics 8 (Aug. 2004), pp. 115–131. DOI: 10.1051/ps:2004007.

[106] Jeffrey P. Spence et al. A flexible modeling and inference framework for estimating variant effect sizes from GWAS summary statistics. Apr. 2022. DOI: 10.1101/2022.04.18.488696.

[107] Fabio Morgante et al. “A flexible empirical Bayes approach to multivariate multiple regression, and its improved accuracy in predicting multi-tissue gene expression from genotypes”. PLOS Genetics 19.7 (July 2023), e1010539. DOI: 10.1371/journal.pgen.1010539.

[108] Deborah Kunkel et al. Improving polygenic prediction from summary data by learning patterns of effect sharing across multiple phenotypes. May 2024. DOI: 10.1101/2024.05.06.592745.

[109] Matthew Stephens. “False discovery rates: a new deal”. Biostatistics (Oct. 2016), kxw041. DOI: 10.1093/biostatistics/kxw041.

## Appendix References

[1] Chiara Auwerx et al. “The individual and global impact of copy-number variants on complex human traits”. The American Journal of Human Genetics 109.4 (Apr. 2022), pp. 647–668. DOI: 10.1016/j.ajhg.2022.02.010.

[2] Benjamin B. Sun et al. “Plasma proteomic associations with genetics and health in the UK Biobank”. Nature 622.7982 (Oct. 2023), pp. 329–338. DOI: 10.1038/s41586-023-06592-6.

[3] Matthew Stephens. “False discovery rates: a new deal”. Biostatistics (Oct. 2016), kxw041. DOI: 10.1093/biostatistics/kxw041.

[4] John D. Storey. “A Direct Approach to False Discovery Rates”. Journal of the Royal Statistical Society Series B: Statistical Methodology 64.3 (Aug. 2002), pp. 479–498. DOI: 10.1111/1467-9868.00346.

[5] Benjamin H Good. “Linkage disequilibrium between rare mutations”. Genetics 220.4 (Apr. 2022), iyac004. DOI: 10.1093/genetics/iyac004.

[6] Xiang Zhu and Matthew Stephens. “Bayesian large-scale multiple regression with summary statistics from genome-wide association studies”. The Annals of Applied Statistics 11.3 (Sept. 2017), pp. 1561– 1592. DOI: 10.1214/17-AOAS1046.

[7] Shun-ichi Amari. “Natural Gradient Works Efficiently in Learning”. Neural Computation 10.2 (Feb. 1998), pp. 251–276. DOI: 10.1162/089976698300017746.

[8] Luigi Malagò and Giovanni Pistone. “Information Geometry of the Gaussian Distribution in View of Stochastic Optimization”. Proceedings of the 2015 ACM Conference on Foundations of Genetic Algorithms XIII. FOGA ‘15. New York, NY, USA: Association for Computing Machinery, Jan. 2015, pp. 150–162. DOI: 10.1145/2725494.2725510.

[9] Larry Armijo. “Minimization of functions having Lipschitz continuous first partial derivatives.” Pacific Journal of Mathematics 16.1 (Jan. 1966), pp. 1–3.

[10] Roy Frostig, Matthew James Johnson, and Chris Leary. “Compiling machine learning programs via high-level tracing”. SysML. Stanford, CA, Feb. 2018.

[11] Charles C. Margossian. “A Review of automatic differentiation and its efficient implementation”. WIREs Data Mining and Knowledge Discovery 9.4 (Nov. 2018), e1305. DOI: 10.1002/WIDM.1305.

[12] Brendan K. Bulik-Sullivan et al. “LD Score regression distinguishes confounding from polygenicity in genome-wide association studies”. Nature Genetics 47.3 (Feb. 2015), pp. 291–295. DOI: 10.1038/ng.3211.

[13] Sarah M. Urbut et al. “Flexible statistical methods for estimating and testing effects in genomic studies with multiple conditions”. Nature Genetics 51.1 (Nov. 2018), pp. 187–195. DOI: 10.1038/s41588-018-0268-8.

[14] Bernard Delyon, Marc Lavielle, and Eric Moulines. “Convergence of a Stochastic Approximation Version of the EM Algorithm”. The Annals of Statistics 27.1 (Feb. 1999), pp. 94–128.

[15] Estelle Kuhn and Marc Lavielle. “Coupling a stochastic approximation version of EM with an MCMC procedure”. ESAIM: Probability and Statistics 8 (Aug. 2004), pp. 115–131. DOI: 10.1051/ps:2004007.

[16] A. P. Dempster, N. M. Laird, and D. B. Rubin. “Maximum Likelihood from Incomplete Data Via the EM Algorithm”. Journal of the Royal Statistical Society: Series B (Methodological) 39.1 (Sept. 1977), pp. 1–22. DOI: 10.1111/j.2517-6161.1977.tb01600.x.

[17] Eli Bingham et al. “Pyro: Deep Universal Probabilistic Programming”. Journal of Machine Learning Research 20.28 (Feb. 2019), pp. 1–6.

[18] Du Phan, Neeraj Pradhan, and Martin Jankowiak. Composable Effects for Flexible and Accelerated Probabilistic Programming in NumPyro. Dec. 2019. DOI: 10.48550/arXiv.1912.11554.

[19] Estelle Kuhn, Catherine Matias, and Tabea Rebafka. “Properties of the stochastic approximation EM algorithm with mini-batch sampling”. Statistics and Computing 30.6 (Nov. 2020), pp. 1725–1739. DOI: 10.1007/s11222-020-09968-0.

[20] Deep Mind et al. The DeepMind JAX Ecosystem. 2020.

[21] Christopher C Chang et al. “Second-generation PLINK: rising to the challenge of larger and richer datasets”. GigaScience 4.1 (Dec. 2015), s13742.–015–0047–8. DOI: 10.1186/s13742-015-0047-8.

[22] Daniel J. Weiner et al. “Polygenic architecture of rare coding variation across 394,783 exomes”. Nature 614.7948 (Feb. 2023), pp. 492–499. DOI: 10.1038/s41586-022-05684-z.

[23] Joshua D. Backman et al. “Exome sequencing and analysis of 454,787 UK Biobank participants”. Nature 599.7886 (Oct. 2021), pp. 628–634. DOI: 10.1038/s41586-021-04103-z.

[24] Nasa Sinnott-Armstrong et al. “Genetics of 35 blood and urine biomarkers in the UK Biobank”. Nature Genetics 53.2 (Jan. 2021), pp. 185–194. DOI: 10.1038/s41588-020-00757-z.

[25] Tony Zeng et al. “Bayesian estimation of gene constraint from an evolutionary model with gene features”. Nature Genetics (July 2024), pp. 1–12. DOI: 10.1038/s41588-024-01820-9.

[26] The UK Biobank Whole-Genome Sequencing Consortium et al. “Whole-genome sequencing of 490,640 UK Biobank participants”. Nature (Aug. 2025), pp. 1–10. DOI: 10.1038/s41586-025-09272-9.

[27] Jian Yang et al. “Conditional and joint multiple-SNP analysis of GWAS summary statistics identifies additional variants influencing complex traits”. Nature Genetics 44.4 (Apr. 2012), pp. 369–375. DOI: 10.1038/ng.2213.

[28] Fergal J Martin et al. “Ensembl 2023”. Nucleic Acids Research 51.D1 (Jan. 2023), pp. D933–D941. DOI: 10.1093/nar/gkac958.

[29] Roshni A. Patel et al. Conditional frequency spectra as a tool for studying selection on complex traits in biobanks. June 2024. DOI: 10.1101/2024.06.15.599126.

[30] Pauli Virtanen et al. “SciPy 1.0: fundamental algorithms for scientific computing in Python”. Nature Methods 17.3 (Mar. 2020), pp. 261–272. DOI: 10.1038/s41592-019-0686-2.

[31] Brendan Bulik-Sullivan et al. “An atlas of genetic correlations across human diseases and traits”. Nature Genetics 47.11 (Sept. 2015), pp. 1236–1241. DOI: 10.1038/ng.3406.

[32] Wolfgang Viechtbauer. “Conducting Meta-Analyses in R with the metafor Package”. Journal of Statistical Software 36 (Aug. 2010), pp. 1–48. DOI: 10.18637/jss.v036.i03.

